# Loss of DOT1L disrupts neuronal transcription, behavior, and leads to a neurodevelopmental disorder

**DOI:** 10.1101/2024.10.31.24314716

**Authors:** Marissa J. Maroni, Melissa Barton, Katherine Lynch, Ashish R. Deshwar, Philip D. Campbell, Josephine Millard, Rachel Lee, Annastelle Cohen, Rili Ahmad, Alekh Paranjapye, Víctor Faundes, Gabriela M. Repetto, Caoimhe McKenna, Amelle L. Shillington, Chanika Phornphutkul, Hanne B. Hove, Grazia M. S. Mancini, Rachel Schot, Tahsin Stefan Barakat, Christopher M. Richmond, Julie Lauzon, Ahmed Ibrahim Elsayed Ibrahim, Caroline Nava, Delphine Héron, Minke M. A. van Aalst, Slavena Atemin, Mila Sleptsova, Iliyana Aleksandrova, Albena Todorova, Debra L. Watkins, Mariya A. Kozenko, Daniel Natera-de Benito, Carlos Ortez, Berta Estevez-Arias, François Lecoquierre, Kévin Cassinari, Anne-Marie Guerrot, Jonathan Levy, Xenia Latypova, Alain Verloes, A. Micheil Innes, Xiao-Ru Yang, Siddharth Banka, Katharina Vill, Maureen Jacob, Michael Kruer, Peter Skidmore, Carolina I. Galaz-Montoya, Somayeh Bakhtiari, Jessica L. Mester, Michael Granato, Karim-Jean Armache, Gregory Costain, Erica Korb

## Abstract

Individuals with monoallelic gain-of-function variants in the histone lysine methyltransferase DOT1L display global developmental delay and varying congenital anomalies. However, the impact of monoallelic loss of *DOT1L* remains unclear.

Here, we sought to define the effects of partial DOT1L loss by applying bulk and single-nucleus RNA-sequencing, ChIP-sequencing, imaging, multielectrode array recordings, and behavioral analysis of zebrafish and multiple mouse models.

We present a cohort of 16 individuals (12 females, 4 males) with neurodevelopmental disorders and monoallelic *DOT1L* variants, including a frameshift deletion, an in-frame deletion, a nonsense, and missense variants clustered in the catalytic domain. We demonstrate that specific variants cause loss of methyltransferase activity. In primary cortical neurons, *Dot1l* knockdown disrupts transcription of synaptic genes, neuron branching, expression of a synaptic protein, and neuronal activity. Further in the cortex of heterozygous *Dot1l* mice, *Dot1l* loss causes sex-specific transcriptional responses and H3K79me2 depletion, including within down-regulated genes. Lastly using both zebrafish and mouse models, we found behavioral disruptions that include developmental deficits and sex-specific social behavioral changes.

Overall, we define how DOT1L loss leads to neurological dysfunction by demonstrating that partial *Dot1l* loss impacts neuronal transcription, neuron morphology, and behavior across multiple models and systems.

## Introduction

Neurodevelopmental disorders (NDDs) are a diverse group of highly prevalent (0.3 – 18.5%)^1^ conditions that manifest during development and impact central nervous system functions.^2,3^ The spectrum of NDDs include intellectual disability, autism spectrum disorder (ASD), attention deficit/hyperactivity disorder (ADHD), communication disorders, specific learning disabilities, and motor disorders.^4^ The cause of NDDs is multifactorial, and includes both inherited and *de novo*, genetic variants with a notable overrepresentation of epigenetic regulators.^5–9^ One subset of epigenetic regulators, histone methyltransferases, are linked to numerous NDDs^10–12^ and function by methylating histones to regulate transcription. Histone methyltransferases are critical for neurogenesis, neuronal migration, neuronal differentiation, synaptic plasticity, and cognition^13^ yet several disease-linked methyltransferases have not yet been studied in the context of neuronal function or animal behavior.

Prior exome sequencing studies identified variants in the histone methyltransferase *DOT1L* as a potential causative driver of NDDs.^6,12^ More recent work identified two variants in *DOT1L* in individuals displaying ADHD^14^ and nine monoallelic (presumed) *de novo* variants of *DOT1L* were identified in individuals with global developmental delay.^15^ Complete loss of DOT1L in mouse models is embryonic lethal^16^ while in *Drosophila*, loss of *grappa*, the *Drosophila DOT1L* ortholog, leads to developmental delay and lethality.^15^ However, *grappa* is highly divergent from DOT1L and thus does not provide an ideal model to study an emerging human disorder. Further, while two previously identified variants were proposed to be gain-of-function based on human cell based-assays^15^ most identified variants have unclear functional consequences. Lastly, while most prior work used full and transmitted knockout models, variants are typically monoallelic and *de novo* making it difficult to define the effect of partial DOT1L disruption from existing data. Thus, the underlying mechanisms linking DOT1L to NDDs remain unclear.

DOT1L is a methyltransferase responsible for depositing mono-, di-, and trimethyl methyl marks on the histone-fold domain on residue 79 of histone H3 (H3K79me)^17,18^, although other enzymes may also deposit the mark.^19^ All three H3K79me states are enriched in gene bodies. However, H3K79me1 is more broadly distributed^20^ whereas H3K79me2/3 is peaks after the transcription start site,^17^ with higher methyl states linked to greater transcriptional output.^20^ DOT1L interacts with RNA polymerase II^21^ and TFIID^22^ and recruits effector proteins such as Menin^23^ to regulate transcription. DOT1L functions in numerous processes, including development^24^ such as in neural progenitor proliferation and differentiation in the cortex, cerebellum, and spinal cord^25–28^ and in maintaining the transcriptional state in differentiating neural progenitors.^29–32^ Further work demonstrated that stress modulates DOT1L expression and H3K79me in the nucleus accumbens and that monoallelic loss of *Dot1l* in the midbrain disrupts synaptic and mitochondrial genes.^33^ Cumulatively, this suggests that DOT1L is critical in neuronal development and neuronal function. Despite these advances, the majority of DOT1L research has focused on biallelic loss of *Dot1l,* which does not reflect the monoallelic nature of individual variants or has not examined effects on development and behavior. Thus, the consequences of monoallelic disruptions of *DOT1L* remain poorly understood.

Here, we identified 16 individuals with monoallelic variants in *DOT1L* displaying a spectrum of neurodevelopmental phenotypes and dysmorphic facial features. Using structural protein modeling, biochemical studies, and patient-derived cells, we found that several variants cause loss of DOT1L methyltransferase activity. Utilizing a *dot1l* knockdown system in zebrafish, we identified disruptions in motor responses to sensory stimuli. Harnessing both bulk RNA-sequencing of primary cultured cortical neurons and single-nucleus RNA-sequencing of mouse cortical tissue, we show that partial loss of *Dot1l* affects transcription of critical neuronal genes linked to synaptic function and causes sex-specific transcriptional responses. Further, cortical neurons display disruptions in neuronal morphology and activity upon *Dot1l* depletion. We demonstrate that H3K79me2 is depleted upon monoallelic *Dot1l* loss in the cortex, especially in down-regulated genes identified in single nucleus RNA-sequencing. Finally, we identified behavioral alterations upon both ubiquitous and neuron-specific monoallelic loss of *Dot1l* in mice. Together, our work demonstrates that partial loss of *Dot1l* causes transcriptional disruptions impacting cognitive function and provides insight into the neurodevelopmental disruptions found in individuals with *DOT1L* variants.

## Materials and methods

The ‘Materials and methods’ are detailed in the Supplementary material.

## Results

### Identification of individuals with neurodevelopmental disorders and monoallelic DOT1L variants

Given the recent discovery of DOT1L’s association with an emerging neurodevelopmental disorder, we searched specifically for individuals harboring variants in *DOT1L*. We identified a cohort of individuals through collaborating clinicians and GeneMatcher^34^ with *DOT1L* variants identified through genome sequencing or exome sequencing. We included individuals with unexplained neurodevelopmental phenotypes and with *DOT1L* rare variants (i.e., not observed in multiple individuals from the general population), where phenotype descriptions were available and there was patient/family consent to be included in a case series.

Using these criteria, we compiled a cohort of 16 probands with variants in *DOT1L*. All individuals have only a single monoallelic variant in *DOT1L*. Inheritance of these variants was *de novo* (10/16), maternal (1/16) or unknown due to one or both parents being unavailable to be sequenced (5/16). Unspecified distinctive craniofacial features and skeletal phenotype were reported in the mother of the individual with the maternally inherited variant. *DOT1L* variant types include missense (13/16), in-frame deletion (1/16), frameshift deletion (1/16), and nonsense (1/16). Notably, all but two of the missense variants (11/13) are within the catalytic domain of DOT1L and affect amino acids that are rarely altered in humans (i.e. dn/ds score of <0.2, except in p.P332R dn/ds = 0.44), indicating intolerance to variation at these sites (Fig. 1A). According to gnomAD (v4.1.0),^35^ DOT1L has a high probability of loss-of-function intolerance (pLI = 1, LOEUF = 0.32) and a high probability of deletion intolerance (pHaplo = 0.98). All variants were absent in gnomAD (v4.1.0) apart from one counted allele of p.L1067Dfs*66. The cohort displays a non-specific constellation of congenital anomalies, including craniofacial anomalies (14/16) such as midface hypoplasia (Fig. 1B and C). Based on the information available at this time, there is no recognizable pattern of morphological differences that would suggest the diagnosis in the absence of molecular genetic testing. Additional individual phenotypes include intellectual disability (4/16), language delay (13/16), motor delay (9/16), a diagnosis of ASD (4/16), and seizures/epilepsy (3/16) (Fig. 1C and Supplementary Table 2). Three additional individuals with high quality, rare, predicted damaging, inheritance unknown variants in *DOT1L* were identified through the MSSNG^36^ database with a diagnosis of ASD but are not included in the main cohort due to lack of additional available information (Supplementary Fig. 1A and B, Supplementary Table 3). Further, four individuals with variants in *DOT1L* also contained additional potential pathogenic variants or had a *DOT1L* variant found in the general population and thus did not meet criteria for inclusion in the cohort. We include them here (Supplementary Fig. 1A and B, Supplementary Table 3) given that they shared some features with the main cohort and that we cannot rule out the possibility of incomplete penetrance of this disorder. Interestingly, while the cohort is not sufficiently powered to confidently determine sex enrichment and the prior smaller cohort^15^ was split roughly equally by sex, 12 out of 16 the individuals in this cohort are female suggestive of possible sex bias.

**Figure 1.**
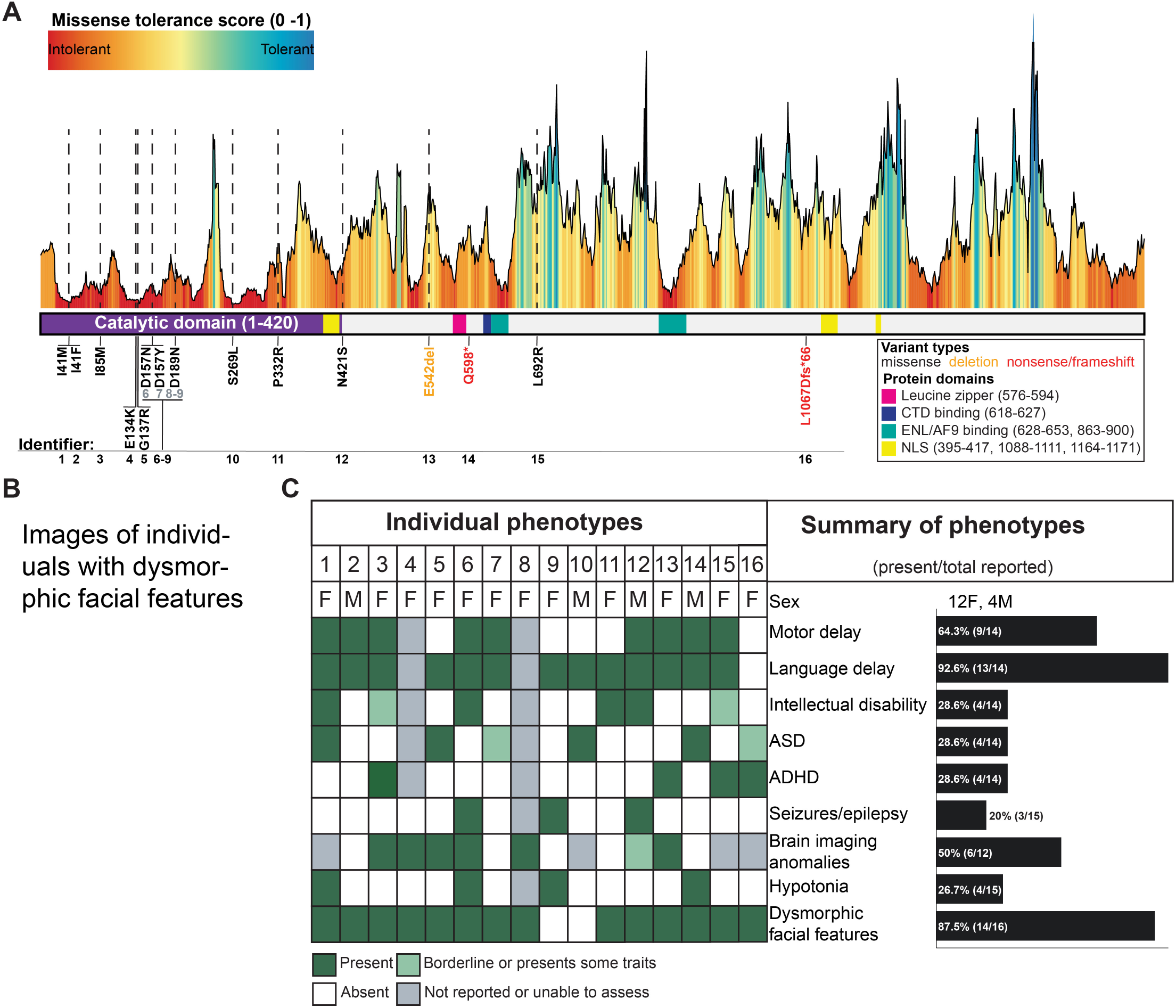
Monoallelic variants in DOT1L are associated with a human neurodevelopmental disorder. (**A**) The landscape of missense tolerance of DOT1L from Metadome. Schematic of DOT1L protein domains (purple = catalytic, pink = leucine zipper, blue = CTD binding, teal = ENL/AF9 binding, and yellow = nuclear localization signal) and locations of *DOT1L* variants (black = missense, orange = deletion, red = nonsense/frameshift). (**B**) Images of dysmorphic facial features in individuals with DOT1L variants. (**C**) *DOT1L* variant individuals’ phenotypic spectrum and prevalences. ASD = autism spectrum disorder. ADHD = attention deficit hyperactivity disorder.

We next determined the location of missense variants in the catalytic domain of DOT1L based on a published structure of DOT1L (PDBID: 6NJ9)^37^ (Fig. 2A). Amino acids with variants are highly conserved across species and are spread throughout the catalytic domain, including regions in close proximity to the binding pocket likely to affect DOT1L function (Fig. 2B and C). To assess the impact of catalytic domain variants on protein stability, we used mutation Cutoff Scanning Matrix (mCSM)^38^. We found that 9/10 missense variants in our cohort were predicted to be destabilizing with an average ΔΔG of -0.411 kcal/mol (Fig. 2D, Supplementary Table 7). Utilizing RING^39^, we examined inter- and intra-protein contacts of residues with missense variants. No variants had interprotein contacts with the nucleosome structure or ubiquitin (Fig. 2D). Variant p.G137R has a contact with S-adenosylmethionine (SAM) suggesting potential effects on DOT1L catalytic activity. Further, variant p.E134K, p.D189N, and p.S269L lie in close proximity to SAM and variant p.P332R is close to where ubiquitin interacts with DOT1L.

**Figure 2.**
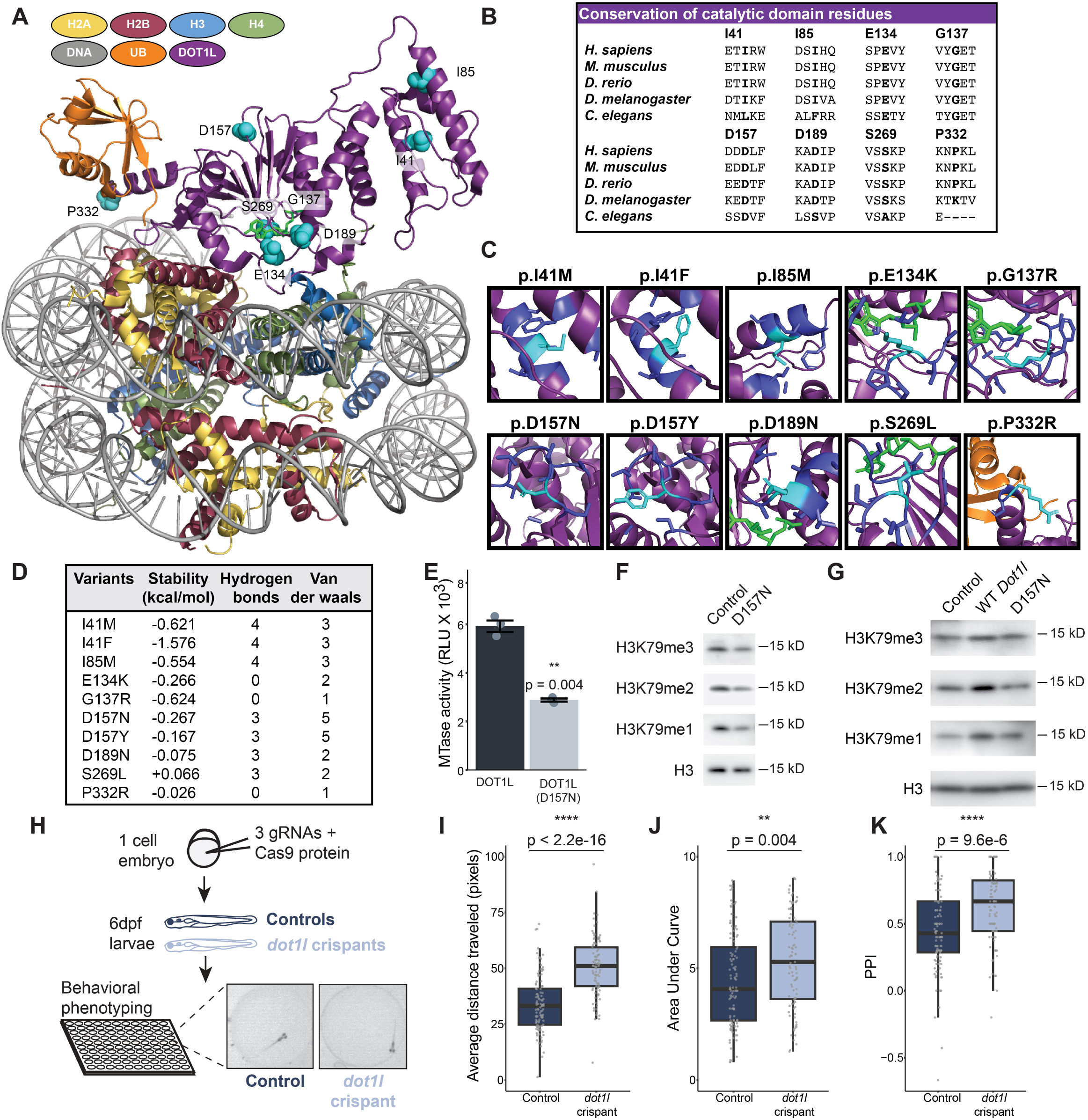
*DOT1L* variants structural and functional analyses and dot1l controls behavioral responses to sensory stimuli in zebrafish. (**A**) Structure of the DOT1L nucleosome complex (PDBID: 6NJ9) highlighting the position of variants (variants = cyan spheres, DOT1L = purple, DNA = grey, H2A = yellow, H2B = red, H3 = blue, H4 = dark green, ubiquitin [UB] = orange, and S-adenosylmethionine [SAM] = green sticks). (**B**) Conservation of amino acids with variants in the catalytic domain across species. (**C**) Local environment of each catalytic domain missense variant (nearby residues = dark blue, defined as residues <=5 angstroms away from the variant residue). (**D**) Predicted stability change from missense variants and the number of nearby residue contacts. (**E**) Methyltransferase (MTase) activity of human DOT1L or variant DOT1L (D157N) on unmodified nucleosomes. Graph shows mean ± SE (n = 3/condition, unpaired two-tailed t-test). RLU = relative light units. (**F**) H3K79 methyl levels in patient-derived fibroblasts from individual 6 (p.D157N variant) beside an age- and sex-matched control. (**G**) H3K79 methyl levels in Neuro-2A cells with variant *Dot1l* (D157N) overexpressed, wildtype (WT) *Dot1l*, or empty vector control. (**H**) Schematic of Cas9 targeting of dot1l and subsequent behavioral phenotyping paradigm performed on zebrafish. (**I**) Average distance traveled during dark flashes in *dot1l* crispant and control zebrafish (control: n = 113, *dot1l* crispant: n = 98; 3 independent experiments, unpaired two-tailed t-test). (**J**) Area under the curve of response to increasing stimulus intensity in *dot1l* crispant and control zebrafish (control: n = 113 control, *dot1l* crispant: n = 95; 3 independent experiments, Kruskal-Wallis Test). (**K**) Prepulse inhibition (PPI) to a medium intensity prepulse acoustic stimulus in *dot1l* crispant and control zebrafish (control: n = 113 control, *dot1l* crispant: n = 95; 3 independent experiments, Kruskal-Wallis Test). All box plot bounds indicate the 25th and 75th percentiles, the black line shows the median, and whiskers extend to the minimum and maximum value that are no further than 1.5X interquartile range. **p<0.01, ****p<0.0001.

Given that most of the variants lie within the catalytic domain, we assessed methyltransferase activity via endpoint histone methyltransferase assays using S-adenosyl homocysteine (SAH) production as a readout of methyltransferase activity. We assessed the p.D157N variant based on the identification of variants at residue 157 in two unrelated individuals. In addition, we selected two previously published variants^15^ (p.R292C and p.E123K) one of which was reported to have no effect (p.R292C) and the other of which was proposed to increase activity (p.E123K) (Supplementary Fig. 2A). Methyltransferase assays demonstrated that both p.D157N and p.R292C reduced methyltransferase activity (Fig. 2E, Supplementary Fig. 2B). In contrast, p.E123K increased activity, as previously reported (Supplementary Fig. 2C).^15^ Further, human fibroblasts harboring the p.D157N had a decrease in all three H3K79me states compared to age- and sex-matched control fibroblasts further supportive of loss of catalytic activity in DOT1L (Fig. 2F, Supplementary Fig. 2D). Lastly, to determine the effect of p.D157N in an orthogonal system without the confound of different genetic backgrounds from primary human fibroblasts, we overexpressed full-length wildtype *Dot1l* and variant *Dot1l* (p.D157N) in mouse Neuro-2A cells. Wildtype *Dot1l* increased H3K79me2 and, to a lesser extent H3K79me1/3 (Fig. 2G). However, variant *Dot1l* (p.D157N) had no detectable impact on H3K79me levels. These data demonstrate that the p.D157N variant has reduced catalytic activity in both a cell-free system and in multiple cell types.

Two variants, p.Q598* and p. p.L1067Dfs*66, cause early stop codons that are predicted to be in regions that will result in nonsense-mediated decay (NMDEscPredictor^40^). Further, these variants truncate DOT1L upstream of 1 (p.L1067Dfs66*) or 2 (p.Q598*) nuclear localization sequences (Fig. 1A), potentially preventing DOT1L from performing established nuclear functions if translated into protein. Together, this suggests that gain-of-function missense variants, loss-of-function missense variants, as well as monoallelic loss of the full protein, are associated with NDDs. Given the discovery of two individuals with nonsense variants and a lack of modeling of monoallelic *Dot1l* loss in the cortex, we chose to examine the effects of partial loss of DOT1L and to better understand the role of DOT1L in the brain.

### *Dot1l* controls behavioral responses to sensory stimuli in zebrafish

Given the *de novo* nature of the majority of individual variants, we first aimed to characterize early behavioral disruptions utilizing a system that allows for allele disruptions in the offspring of zebrafish. Importantly, zebrafish provide a vertebrate model with high genetic similarity to humans,^41^ including DOT1L (Catalytic domain: 85% identity, whole gene: 49% identity, 57% similarity, 21% gaps)^42^ (Fig. 2B). Zebrafish also develop robust stereotypical motor movements in response to sensory stimuli (visual or acoustic) detectable within the first six days of development. Prior work demonstrated that these behaviors are sensitive to mutations in genes associated with NDDs,^43–45^ suggesting their relevance to NDD pathophysiology.

To assess behavioral roles for DOT1L in early development, we first specifically disrupted the zebrafish *dot1l* gene using a CRISPR-Cas9-approach that generates biallelic null alleles^46^ in over 90% of animals. Notably, maternally deposited mRNA contribute to the zebrafish oocyte, and RNA sequencing evidence demonstrates that *dot1l* is expressed in the ovary and early embryo prior to zygotic gene activation (GRCz11).^47^ Thus, while full loss of *dot1l* is lethal in other model organisms, this system is expected to result in a partial loss of *dot1l* and typically allows organisms to survive early development even in cases where full knockout is expected to be lethal.^44,48^ We injected three guide RNAs that target non-overlapping sites along the *dot1l* gene into every fertilized embryo together with Cas9 protein, generating *dot1l* ‘crispants’ (Supplementary Fig. 2E). Control embryos were injected in parallel with three non-targeting gRNAs and Cas9. Using TIDE^49^, a computational approach that asses genome editing by Sanger sequence trace decomposition, we confirmed that each gRNAs targeted *dot1l* and efficiently introduced indels (Supplementary Fig. 2F and G). *Dot1l* crispants, were viable to 6 days post-fertilization (dpf) and did not display obvious gross morphological defects (control injected n=164, *dot1l* crispant n=144, four independent experiments). Behavior of *dot1l* crispants was then assessed at 6 dpf using a previously described pipeline that allows assessment of multiple sensorimotor behaviors including the visual motor response, responsiveness to flashes of light or darkness, and the acoustic startle response^43^ (Fig. 2H, Supplementary Fig. 2H). Compared to controls, *dot1l* crispants displayed exaggerated motor responses to multiple sensory inputs. Specifically, *dot1l* crispants displayed increased movement in response to changes in illumination, as illustrated by increased distance travelled in the visual motor response assay^50^ (Supplementary Fig. 2I) and increased movement in response to flashes of darkness^51^ (Fig. 2I). In addition, *dot1l* crispants are hypersensitive to acoustic stimuli, displaying startle responses^52^ following stimuli that do not elicit similar responses in controls (Fig. 2J). Further, *dot1l* crispants also show an increase in acoustic startle prepulse inhibition^53^ compared to controls (Fig. 2K). Together, these results demonstrate that zebrafish *dot1l* controls responses to visual stimuli and is required for establishing the acoustic startle threshold and acoustic startle sensorimotor gating in zebrafish. Further, they demonstrate that DOT1L loss affects early developmental behaviors.

### DOT1L regulates glutamatergic synaptic gene expression

Based on the broad neurodevelopmental phenotypes observed in individuals with *DOT1L* variants, and the robust effects of DOT1L loss in zebrafish behavior, we next tested the role of DOT1L in mouse models based on highly conserved DOT1L (Catalytic domain: 96% identity, whole gene: 84% identity, 88% similarity, 1% gaps)^42^ (Fig. 2B). We began by defining the regulation of *Dot1l* and its target histone modification H3K79me in developing mouse neurons. We leveraged mouse primary cultured neurons derived from E16.5 cortices to generate a pure neuronal population and found that both *Dot1l* mRNA expression and H3K79me increase throughout neuronal maturation (Supplementary Fig. 3A and B). To model partial loss of *Dot1l*, we infected primary cortical neurons with short hairpin RNAs (shRNA) targeting *Dot1l* or a non-targeting control (n.t.). We confirmed *Dot1l* knockdown and H3K79me2/3 depletion upon infection of *Dot1l* shRNAs (Supplementary Fig. 3C and D) demonstrating that continued DOT1L expression is required for H3K79me deposition in developing neurons.

Given the association between H3K79me and active gene expression, we next sought to determine the effect of *Dot1l* knockdown on transcription in primary neurons. Following *Dot1l* depletion, we performed RNA-sequencing and found widespread changes in gene expression with 677 genes significantly up-regulated and 1050 genes significantly down-regulated (Fig. 3A). The majority of gene expression changes had fold changes less than 1 (838 down and 526 up) suggesting that *Dot1l* depletion broadly effects many genes but to a modest degree. Gene ontology (GO) analysis indicated an enrichment of genes involved in synaptic transmission (such as GO:0099177 and GO:0050804) in down-regulated differentially expressed genes (DEGs) and no significant enrichment of GO terms in up-regulated DEGs (Fig. 3B). Given the dysregulation of synaptic-related genes, we further interrogated differentially expressed genes using SynGO^54^ which demonstrated enrichment for pre- and post-synaptic compartment proteins, and synaptic cleft proteins suggesting widespread disruption of expression of synaptic genes (Supplementary Fig. 3E). We next asked whether the observed changes in genes related to synaptic transmission were global or specific to a class of chemical synaptic transmission. To this end, we used gene set enrichment analysis to test for enrichment of genes related to glutamatergic, GABAergic, dopaminergic, and cholinergic synaptic transmission. Interestingly, glutamatergic transmission is enriched in down-regulated genes, while there is no significant enrichment of other classes of synaptic transmission (Fig. 3C, Supplementary Fig. 3F-H). In fact, down-regulated DEGs had significant overlap with the glutamatergic synaptic transmission gene set including genes such as *Gria2* and *Grin1*, two glutamate receptor subunits that are critical for appropriate levels of glutamatergic transmission throughout the brain (Fig. 3D, Supplementary Fig. 3I-K). Together these findings demonstrate that *Dot1l* depletion disrupts expression of critical synaptic genes.

**Figure 3.**
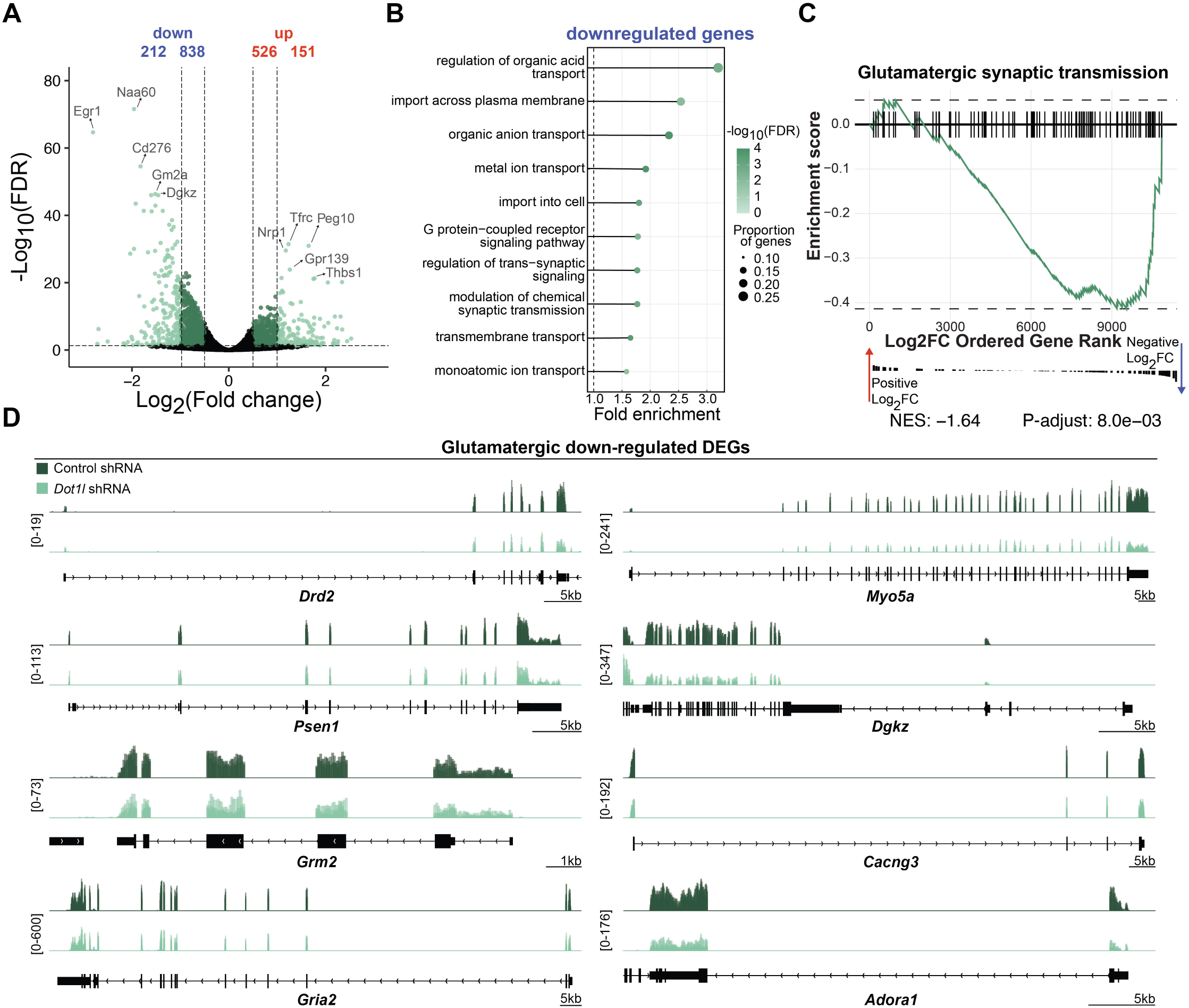
*Dot1l* promotes glutamatergic synaptic gene expression in neurons. (**A**) Volcano plot showing differentially expressed genes (DEGs) from primary cortical neurons infected with *Dot1l* shRNA or non-targeting control shRNA (n = 3 biological replicates/condition). Light green dots represent genes with log2(Fold change) >= 1 and FDR <= 0.05 and dark green dots represent genes with log2(Fold change) >= 0.5 and FDR <= 0.05. (**B**) Biological process gene ontology analysis of down-regulated genes. (**C**) Gene set enrichment analysis of genes involved in glutamatergic synaptic transmission. NES = normalized enrichment score. (**D**) RNA-seq genome browser tracks for down-regulated glutamatergic genes including: *Drd2, Myo5a, Psen1, Dgkz, Grm2, Cacng3, Gria2,* and *Adora1*.

### *Dot1l* promotes neuronal arborization, GluA2 levels, and neuronal activity

Given the disruption to expression of synaptic genes that we detected, we next tested the effect of *Dot1l* knockdown on neuronal morphology, synapses, and synaptic proteins. Neuronal branching and spine formation are critical components of neuronal maturation that allow for neuronal communication and downstream behaviors. To assess how *Dot1l* loss impacts neuronal architecture, we performed Sholl analysis on primary cortical neurons transfected with a *Dot1l* shRNA or control shRNA (Fig. 4A). *Dot1l* depleted neurons had a reduced number of intersections in comparison to controls indicative of reduced neuronal arborization (Fig. 4B and C). In addition to neuronal branching, spine formation is critical for synapse development and essential for neuronal communication and memory consolidation. Interestingly, the spine density of *Dot1l* depleted neurons was increased (Supplementary Fig. 4A). This could suggest aberrant spine development as seen in other developmental disorders^55^ or be a compensatory mechanism to offset the loss of neuronal branching or decreased glutamatergic synapse function. Given the downregulation of genes involved in glutamatergic transmission, we also assessed whether the glutamatergic receptor subunit GluA2 protein is regulated by DOT1L in primary neurons. Using immunocytochemistry, we found that GluA2 is depleted upon *Dot1l* loss demonstrating that transcriptional disruptions functionally affect protein levels of critical synaptic genes (Fig. 4D and E). Conversely, overexpression of *Dot1l* in neurons did not result in significant changes to neuronal arborization, spine density, or GluA2, suggesting that increased expression of *Dot1l* alone is not sufficient to enhance expression of GluA2 or alter neuronal architecture (Supplementary Fig. 4B-G). To examine whether these structural alterations have functional consequences, we recorded neuronal activity from *Dot1l* depleted neurons using a multielectrode array (Fig. 4F and G, Supplementary Fig. 4H). *Dot1l* depleted neurons did not have significantly altered firing rates (number of spikes per second) (Supplementary Fig. 4I) but had markedly reduced burst frequency, including network burst frequency (Fig. 4H and I). These findings held regardless of whether burst frequency was normalized by electrode activity (Supplementary Fig. 4J and K). Further, *Dot1l* depletion increased the interquartile range of network burst duration indicating greater variability in network burst duration (Fig. 4J). These findings support a functional role of changes identified through bulk-RNA-sequencing on gene targets with known roles in neuronal activity such as calcium influx (*Gria1, Grin1, Cacng3, Cacng4, Cang5*). Together, DOT1L loss affects neuronal morphology and synaptic protein content, ultimately resulting in functional differences in firing patterns in mature neurons.

**Figure 4.**
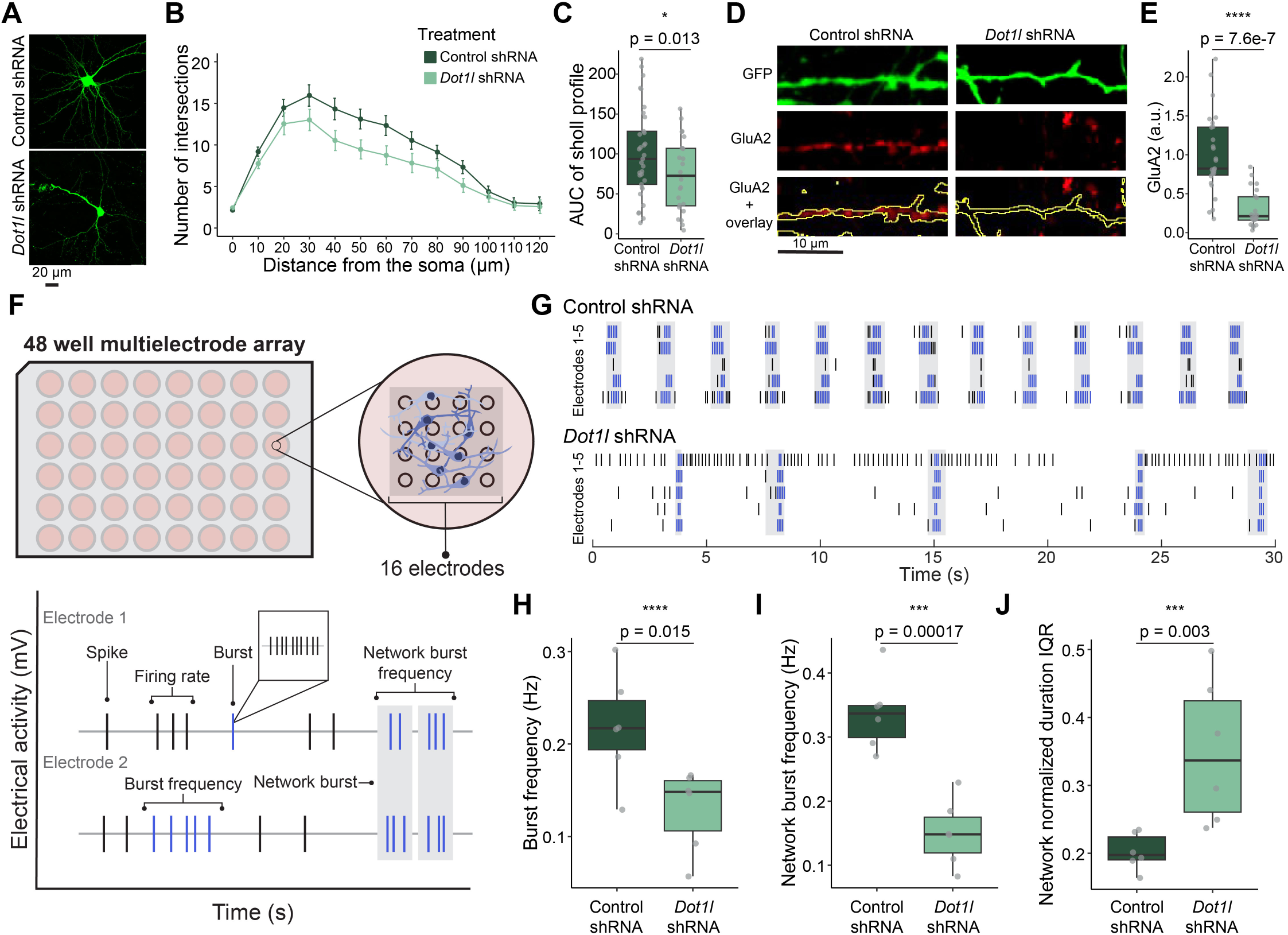
*Dot1l* promotes neuronal arborization, GluA2 levels, and neuronal activity. (**A**) Representative images of primary cortical neurons transfected with *Dot1l* shRNA or non-targeting control shRNA. Scale bar = 20 μm. (**B**) Number of branch intersections per radius of shRNA transfected neurons (control: n = 33 neurons from 5 biological replicates, *Dot1l* shRNA: n = 24 neurons from 5 biological replicates). (**C**) Area under the curve (AUC) quantification of (**B**) (control: n = 33 neurons from 5 biological replicates, *Dot1l* shRNA: n = 24 neurons from 5 biological replicates, mixed effect model). (**D**) Representative images of GluA2 staining in shRNA-transfected neurons. (**E**) Quantification of GluA2 staining in (**D**) (control: n = 27 neurons from 3 biological replicates, *Dot1l* shRNA: n = 22 neurons from 3 biological replicates, Kruskal-Wallis Test). (**F**) Schematic of multielectrode array apparatus and neuronal activity metrics. (**G**) Representative raster plot showing spike activity on individual electrodes in *Dot1l* shRNA infected neurons and controls (black line = spike, blue line = single electrode burst, gray = network burst, minimum of 5 spikes with inter-spike interval <100ms). (**H-J**) Activity metrics in shRNA infected neurons at 20 days *in vitro* that include (**H**) burst frequency (unpaired two-tailed t-test), (**I**) network burst frequency (unpaired two-tailed t-test), (**J**) and network normalized interquartile range (IQR) (control: n = 6 wells from 3 biological replicates, *Dot1l* shRNA: n= 6 wells from 3 biological replicates, Kruskal-Wallis Test). All box plot bounds indicate the 25th and 75th percentiles, the black line shows the median, and whiskers extend to the minimum and maximum value. *p<0.05, ***p<0.001, ****p<0.0001.

### DOT1L regulates cortical gene expression in a sex-specific manner

Given the transcriptional disruptions in our *in vitro* primary cultured neurons and behavioral disruptions in our zebrafish model, we next sought to analyze the transcriptional effects of monoallelic loss of *Dot1l* in mice to model the monoallelic loss in individuals with *DOT1L* variants. We first examined *Dot1l* mRNA expression and H3K79me deposition in mice during cortical development from E14 through postnatal day 28. We found that both *Dot1l* and H3K79me increase during this period, suggesting that DOT1L may play a role during this critical period of brain development (Supplementary Fig. 5A-D). Prior work thoroughly defined the effects of *Dot1l* loss on the transcriptome of stem cell populations and the effect of biallelic *Dot1l* loss on neurons early in development.^25–32^ However, to the best of our knowledge the transcriptional effect of monoallelic loss has only been tested in the midbrain^33^ with a focus on aging-related phenotypes and has not been tested beyond early development in brain regions relevant to the emerging disorder described here.

Given the notable increase of *Dot1l* mRNA expression and H3K79me from P0-P28 and the lack of characterization of *Dot1l* after brain development, we assessed the transcriptional impact of monoallelic loss of *Dot1l* in 10-week-old cortical mouse tissue. We used a floxed *Dot1l* mouse model containing loxP sites flanking exon 2 of *Dot1l* that was crossed to a ubiquitously expressing Cre line under the human cytomegalovirus (CMV) minimal promoter that expresses during early embryogenesis^56^ to generate *Dot1l*^floxed/+^;CMV-Cre^+/-^ (referred to as *Dot1l* HET) with littermate controls (*Dot1l*^+/+^;CMV-Cre^+/-^). Notably, parental lines that generated experimental cohorts included *Dot1l*^floxed/+^ crossed to CMV-Cre^+/+^. This ensures that parents of experimental mice have wildtype DOT1L expression to avoid effects of parental loss of *Dot1l* which may affect the health of offspring and to better mimic the affected individuals in which most variants are *de novo*.

We first confirmed depletion of *Dot1l* in *Dot1l* HET cortical tissue as expected (Supplementary Fig. 5E). Given that prior work established effects of complete *Dot1l* loss on neurogenesis and cortical layer development and our data demonstrating that *Dot1l* knockdown robustly affects gene expression within neurons, we harnessed single nucleus RNA-sequencing to capture both changes in cell type identity and changes in gene expression in cortical tissue (Fig. 5A). Using three male and three female animals for both control and *Dot1l* HET, we identified 25 clusters that include 10 excitatory neurons clusters (Slc17a7+), 7 inhibitory neuron clusters (Gad2+), 2 microglia clusters (Ctss+ and Ptprc+), an astrocyte cluster (Gja1 + and Gnb4+), and an oligodendrocyte cluster (Mog+, Enpp6+, and Opalin +) (Supplementary Fig. 5F). Interestingly, we did not find altered proportions of neuronal cell types and only a modest increase in microglia cell types in *Dot1l* HET mice in comparison to control suggesting that monoallelic *Dot1l* loss is not sufficient to alter cortical neuron identity as occurs following complete *Dot1l* deletion^25^ (Fig. 5B). However, we found widespread disruption of gene expression across most excitatory and inhibitory neuron clusters and modest changes in non-neuronal clusters (Fig. 5C and Supplementary Fig. 5G, 5H). We detected the greatest effects in excitatory neuron clusters (Fig. 5C) and thus examined the effect of *Dot1l* loss on excitatory clusters as a whole. We found 880 significantly down-regulated genes and 310 genes significantly up-regulated upon *Dot1l* loss *in vivo* fitting with culture data demonstrating more genes are decreased in gene expression following partial *Dot1l* loss (Fig. 5D). Gene ontology analysis indicated an enrichment of genes involved in synaptic function (GO:0140236, GO:0140241) and metabolic processes (GO:0006091, GO:0009141, GO:0009144, GO:0009199) in down-regulated DEGs and no significant enrichment of GO terms in up-regulated DEGs (Fig. 5E). Examining the excitatory cluster with the most DEGs (Ex_L2/3_1), showed similar effects with 602 significantly down-regulated genes and 221 genes significantly up-regulated (Fig. 5F). Gene ontology analysis of downregulated genes again indicated disruption of genes involved in synaptic function (GO:0016188, GO:0048167, GO:0099003, GO:0099504), a feature that was echoed in inhibitory neuron clusters as well (Fig. 5G and Supplementary Fig. 5I and J).

**Figure 5.**
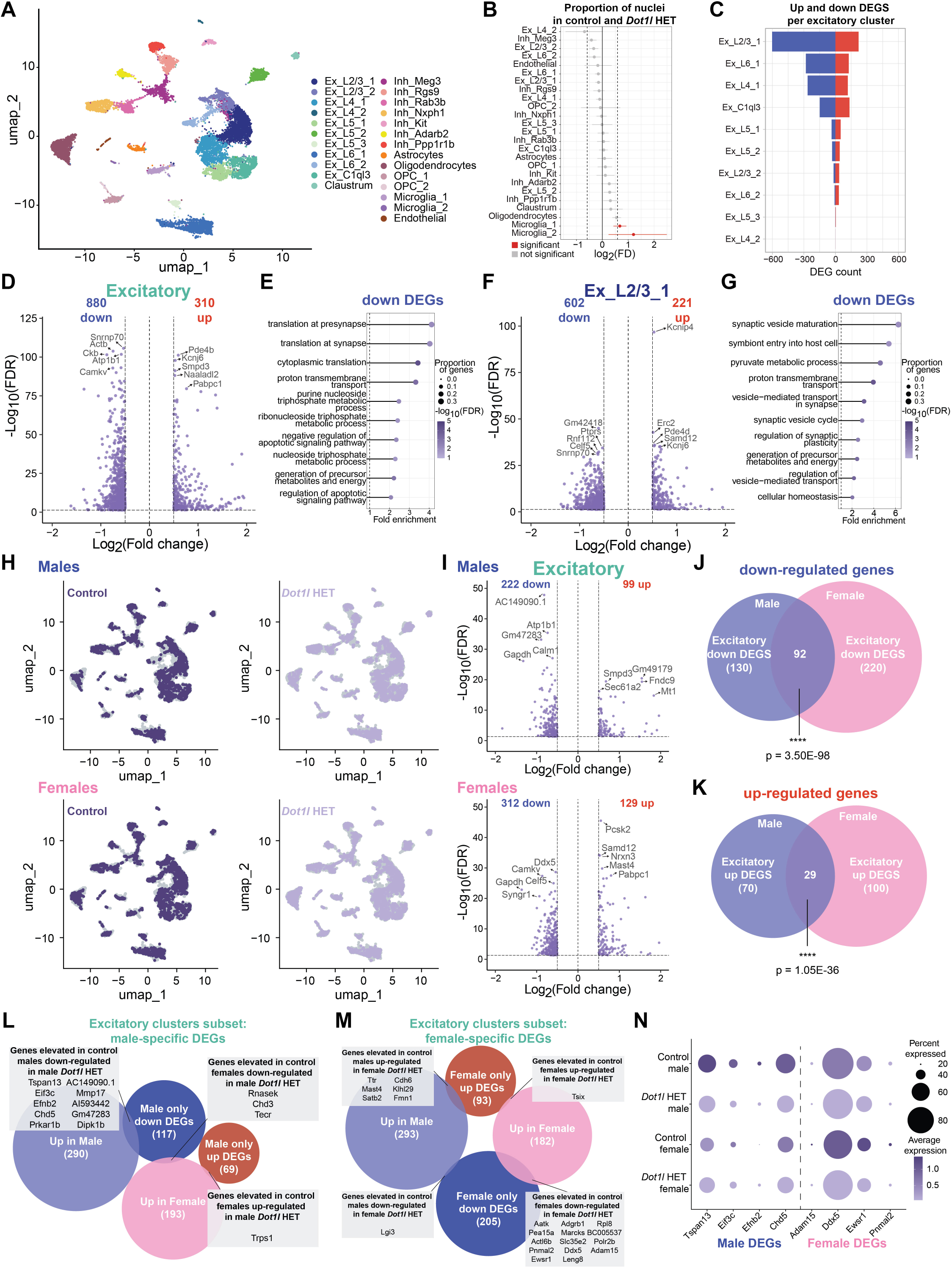
*Dot1l* regulates cortical gene expression in a sex-specific manner. (**A**) UMAP (Uniform Manifold Approximation and Projection for Dimension Reduction) of single nucleus RNA sequencing from 10-week-old Dot1l HET and control mouse cortex (n = 6 cortices per group [3 males, 3 females). (**B**) Proportion of nuclei in *Dot1l* HET and control in each cluster. Red indicates fold difference (FD) > 1.8 and FDR < 0.05. (C) Number of up- and down-regulated differentially expressed genes (DEGs) in excitatory clusters. (**D**) Volcano plot showing DEGs from *Dot1l* HET and control combined excitatory clusters. (**E**) Biological process gene ontology analysis of down-regulated DEGs from combined excitatory clusters. (**F**) Volcano plot showing DEGs from *Dot1l* HET and control from Ex_L2/3_1. (**G**) Biological process gene ontology analysis of down-regulated DEGs from Ex_L2/3_1. (**H**) UMAP clustering with dots representing nuclei from *Dot1l* HET or control separated by sex. (**I**) Volcano plots showing DEGs from male or female *Dot1l* HET and control from combined excitatory clusters. (**J-K**) Overlap of male and female (**J**) down and (**K**) up DEGs from combined excitatory clusters (hypergeometric test). (**L-M**) Overlap of (**L**) male and (**M**) female DEGs with genes up-regulated in male or female controls. (**N**) Dot plot showing genes elevated in male or female control mice that are sex-specifically down-regulated in *Dot1l* HETs. ****p < 0.00001.

Given that the majority of individuals within the cohort were female, we interrogated whether there may be sex-specific transcriptional alterations upon monoallelic *Dot1l* loss. To parse sex-specific effects, we separated male and female nuclei and found the sexes were equally represented in each cluster (Fig. 5H). We again detected widespread gene expression changes in both male and female excitatory neuron clusters upon monoallelic *Dot1l* loss (Fig. 5I). However, we detected slightly more down-regulated genes in female neurons with 220 uniquely down-regulated in female, 130 genes uniquely down-regulated in males, and 92 down-regulated genes shared between males and females (Fig. 5J). This effect was similar in up-regulated genes with 100 uniquely up-regulated in females and 70 uniquely up-regulated in males (Fig. 5K). Gene ontology analysis of these three down-regulated gene sets (males only, females only, and shared) indicated disruption of biological processes related to synaptic function in the males-specific DEGs, neuronal outgrowth in shared DEGs, and no significantly enriched biological processes in the female-specific DEGs (Supplementary Fig. 5K and L). Interestingly, motif analysis indicated that the enriched transcription factor binding sites differ between male and female-specific DEGs (Supplementary Fig. 5M). These findings suggest that male and female-specific DEGs may contribute to distinct behavioral responses to DOT1L loss and may be influenced by different transcription factor usage.

To assess whether there are baseline sex differences that may influence sensitivity to monoallelic *Dot1l* loss, we sought to compare DEGs with genes that are differentially expressed between sexes under control conditions. First, we examined baseline gene expression differences in male and female cortices. Although modest in fold change, there were 300 genes elevated in control males and 197 genes elevated in control females, including expected X and Y linked genes (Supplementary Fig. 5N). In *Dot1l* HETs, the sex differences were more modest with 50 elevated in males and 43 elevated in females (Supplementary Fig. 5O). While X and Y linked gene expression differences were still detected, many somatic chromosome sex-specific gene expression differences are not shared between controls and *Dot1l* HETs suggesting that baseline sex transcriptional differences are disrupted upon monoallelic *Dot1l* loss (Supplementary Fig. 5P and Q). In addition, a subset of male versus female DEGs are differentially expressed upon monoallelic *Dot1l* loss in a sex-specific manner (Fig. 5L-N and Supplementary Fig. 5R). This suggests baseline gene expression differences in male and female brains partially influences sensitivity to monoallelic *Dot1l* loss. Together, these findings demonstrate that there are both common and sex-specific transcriptional programs down-regulated upon monoallelic *Dot1l* loss. Further, these findings suggest that transcription factor usage contributes to some sex-specific effects while others may be due to underlying differences in transcriptional states for a small but potentially important subset of genes.

Finally, we compared *in vivo* and *in vitro* RNA-sequencing gene sets. Genes unique to each system were identified as expected due to the difference in methods (whole cell analysis in the *in vitro* system versus nuclei-specific analysis in the *in vivo* system) and due to the differences in length and method of DOT1L depletion (5-day knockdown verses long-term genetic depletion). However, despite these differences, we identified 69 down-regulated genes shared between our *in vitro Dot1l* shRNA dataset and the *in vivo Dot1l* HET dataset suggesting shared transcriptional disruptions between even highly extrinsic and systemic models of partial *Dot1l* loss (Supplementary Fig. 5S).

### Monoallelic *Dot1l* loss causes a global reduction of H3K79me2

Given that *DOT1L* variants cluster in the catalytic domain and at least two variants (p.D157N and p.R292C) disrupt H3K79 methyltransferase activity, we next sought to determine the relationship between H3K79me and gene expression in the brain. We performed ChIP-sequencing for H3K79me2 on cortical tissue from 10-week-old male and female *Dot1l* HET mice and controls. Aligning with prior work, H3K79me2 deposition peaks after the transcription start site and decreases throughout the gene body and (Fig. 6A). Given the broad distribution of H3K79me2, we examined read counts normalized by gene length rather than peak calling for analysis. We found that the amount of H3K79me2 deposition correlates with gene expression levels (Fig. 6B). Further, *Dot1l* HETs have a global reduction of H3K79me2 compared to controls regardless of the base level of gene expression in that gene (Fig. 6C and D). We next compared down, up, and non-DEGs identified in excitatory neuron clusters and found that only down-regulated and non-DEGs lost H3K79me2 (Fig. 6E). The reduction of H3K79me2 in down-regulated DEGs was greater than non-differentially expressed genes suggesting the degree of H3K79me2 loss directly relates to the effect on gene expression (Fig. 6F). Given the sex-specific transcriptional disruption, we examined whether this could be due to sex-specific deposition of H3K79me2. We found that H3K79me2 was similarly reduced in both males and females and, unlike transcriptional changes, there were no baseline differences in male and female control mice in amount of H3K79me2 deposition globally or in up, down, and non-DEGs (Fig. 6G, Supplementary 6A). Similarly, both male and female *Dot1l* HET mice showed the greatest H3K79me2 loss in down DEGs with a more subtle loss seen in non-DEGs (Supplementary 6B-D). Further we found that regardless of whether a gene was down-regulated only in male or female mice, *Dot1l* monoallelic loss caused a reduction of H3K79me2 (Fig. 6H-J). This suggests that sex-specific transcriptional changes are not due to greater H3K79me2 loss. Rather baseline gene expression differences or other differences between male and female neurons likely cause particular genes to be more sensitive to the loss of H3K79me2.

**Figure 6.**
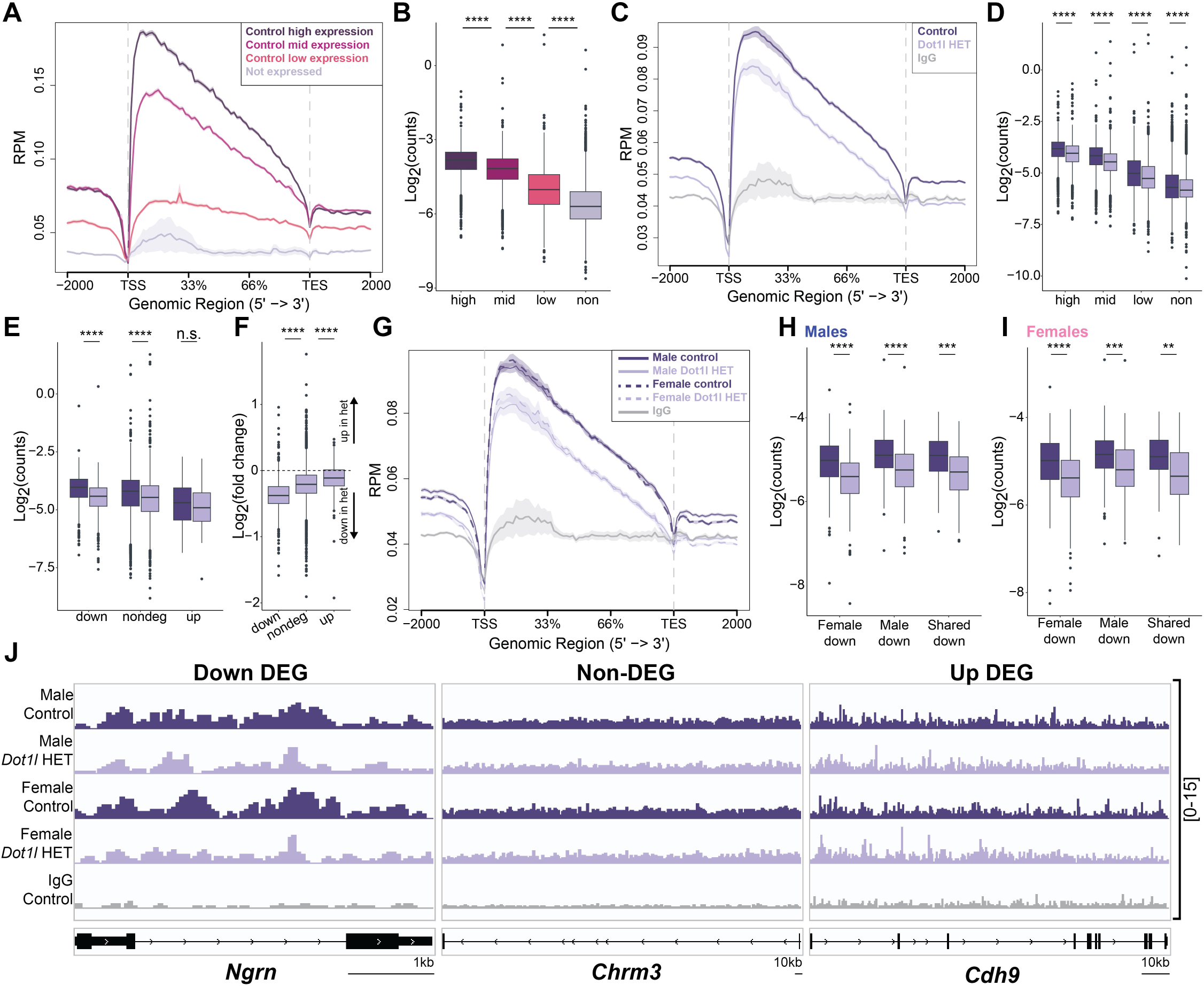
Monoallelic *Dot1l* loss causes a global reduction of H3K79me2. (**A**) Metaplot of H3K79me2 ChIP in control cortical nuclei binned by expression level. Plot shows read counts per million mapped reads (RPM) between the transcription start site (TSS) and transcription end site (TES) ± 2 kb (n = 4 biological replicates, 2 females, 2 males/genotype). Not expressed was defined as genes with an average expression < 0.01 by single nucleus RNA sequencing (n = 7,824 genes). Remaining genes were binned into three equally sized groups by average expression (n = 5,055 genes per group). (**B**) H3K79me2 log2(counts) binned by gene expression level (one-way ANOVA with post hoc pairwise t-tests with Bonferroni correction). (**C**) Metaplot of H3K79me2 ChIP in control and *Dot1l* HET cortical nuclei in all genes. (**D**) H3K79me2 log2(counts) binned by gene expression level in *Dot1l* HET and control (two-way ANOVA with post hoc Tukey HSD). (**E**) H3K79me2 log2(counts) binned by down DEGs, non-DEGS, and up DEGs (two-way ANOVA with post hoc Tukey HSD). (**F**) H3K79me2 log2(fold change) of *Dot1l* HET in comparison to controls (one-way ANOVA with post hoc pairwise t-tests with Bonferroni correction). (**G**) Metaplot of H3K79me2 ChIP in male and female control and *Dot1l* HET cortical nuclei. (**H**) H3K79me2 log2(counts) in male *Dot1l* HET and control binned by female down DEGs, male down DEGs, and shared DEGs (two-way ANOVA with post hoc Tukey HSD). (**I**) H3K79me2 log2(counts) in female *Dot1l* HET and control binned by female down DEGs, male down DEGs, and shared DEGs (two-way ANOVA with post hoc Tukey HSD). (**J**) H3K79me2 ChIP-seq genome browser tracks for *Ngrn, Chrm3,* and *Cdh9*. *p<0.05, **p < 0.01, ***p<0.001, ****p<0.0001.

### Monoallelic *Dot1l* loss alters early vocalization development and sociability

Previous studies using homozygous *Dot1l* mouse knockouts indicate that it is essential for hematopoiesis,^57^ cardio myocyte function,^58^ and neural progenitor proliferation and differentiation in the cortex, cerebellum, and spinal cord.^25–28^ However, defining the developmental and behavioral responses to monoallelic loss of *Dot1l* is critical to understand the implications of monoallelic variants in affected individuals. As previously reported,^16^ *Dot1l* HETs are viable and generated in approximately expected Mendelian ratios (Supplementary Fig. 7A and B). To assess impacts of monoallelic *Dot1l* loss on early development, we tracked developmental milestones including physical landmarks, and sensorimotor development in *Dot1l* HET and controls. Male *Dot1l* HET pups had no differences in weight but had delayed development of the visual placing response, a measurement of sensory development (Fig. 7A, Supplementary Fig. 7C). Female *Dot1l* HET pups weighed more than controls but had no delayed development (Fig. 7A, Supplementary Fig. 7C). Given the language delay seen in 13/16 individuals with *DOT1L* variants, and previous studies demonstrating ultrasonic vocalizations (USVs) changes in various NDD mouse models,^59^ we measured USVs in pups during 5 minutes of maternal separation at P6. Male *Dot1l* HET pups had significantly higher decibel calls, and a greater percentage of chevron type calls (Fig. 7B-E). Female *Dot1l* HET mice had decreased total USV calls with no differences in call characteristics (Fig. 7B-E). Finally, we found both male and female *Dot1l* HET mice were slower to complete a negative geotaxis assay where mice are placed face down on an angled platform to assess early motor and vestibular development (Fig. 7F).

**Figure 7.**
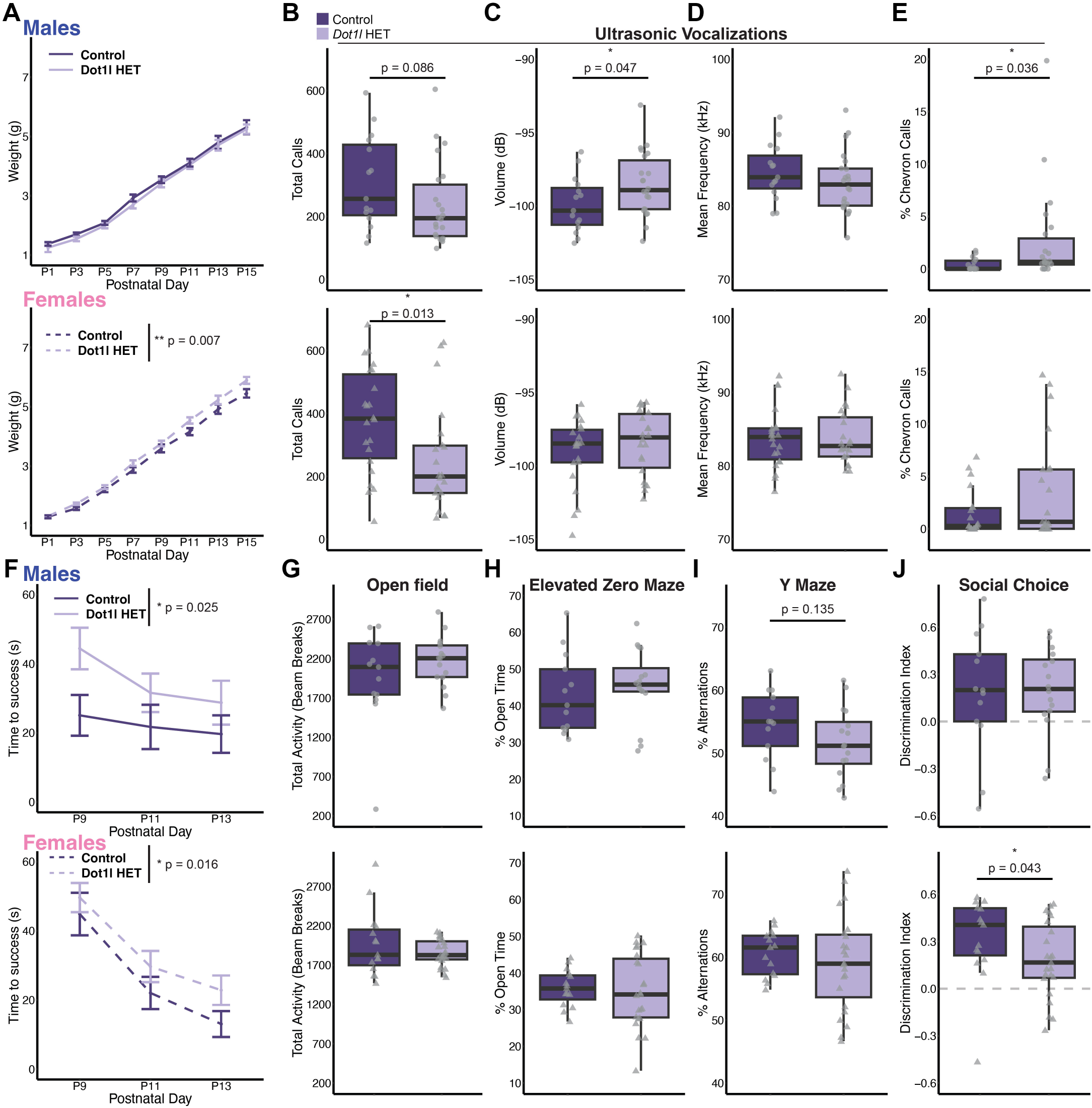
Monoallelic *Dot1l* loss alters early vocalization development and sociability. (**A**) Weight during first two weeks post-birth in male and female mice (male [control: n = 13, *Dot1l* HET: n = 16]; female [control: n = 15, *Dot1l* HET: n = 23], repeated measures ANOVA). (**B**) Total number of ultrasonic vocalizations (USVs) in P6 male and female pups (male [control: n = 15, *Dot1l* HET: n = 22]; female [control: n = 22, *Dot1l* HET: n = 22], Kruskal-Wallis Test). (**C**) Average volume in decibels (dB) of USVs in male and female pups (unpaired two-tailed t-test). (**D**) Average frequency in kilohertz (kHz) of calls in male and female pups. (**E**) Percent chevron calls out of total calls in male and female pups (Kruskal-Wallis Test). (**F**) Time to right self in negative geotaxis assay during development in male and female pups (male [control: n = 13, *Dot1l* HET: n = 16]; female [control: n = 15, *Dot1l* HET: n = 23], repeated measures ANOVA). (**G**) Total activity measured from beam breaks during 10-minute open field assay in 4-week-old male and female mice (male [control: n = 13, *Dot1l* HET: n = 16]; female [control: n = 15, Dot1l HET: n = 23], Kruskal-Wallis Test). (**H**) Percent of time spent in open arms of the elevated zero maze in male and female mice (unpaired two-tailed t-test). (**I**) Percent spontaneous alternations out of the total number of triads possible in male and female mice. (unpaired two-tailed t-test). (**J**) Discrimination index between interaction time with the mouse or rock cylinder during the three-chamber social choice assay in male and female mice (Kruskal-Wallis Test). All box plot bounds indicate the 25th and 75th percentiles, the black line shows the median, and whiskers extend to the minimum and maximum value that are no further than 1.5X interquartile range. *p<0.05, **p<0.01.

Next, we performed a battery of behavioral assays to assess motor and cognitive function in juvenile *Dot1l* HETs and controls. We found no impairments in gross motor function in *Dot1l* HET mice in an open field assay (Fig. 7G, Supplementary Fig. 7D). Further, there was no evidence of anxiety-related behaviors measured using percent of time spent in open arms of the elevated zero maze and percent of time spent in the center of the open field assay (Fig. 7H, Supplementary Fig. 7E and F). We also detected no changes in working memory in *Dot1l* HETs in comparison to controls measured using percent of spontaneous alternations completed in a Y maze (Fig. 7I, Supplementary Fig. 7G). To assess sociability, we performed the social choice assay where mice explore the three-chamber arena with one chamber holding a rock, one chamber holding a mouse, and a neutral center chamber. Female *Dot1l* HETs had a reduced time spent with the mouse measured using a discrimination index (time spent with mouse – time spent with rock / total interaction time) indicating sex-specific social behavior changes (Fig. 7J). Together, these data demonstrate that *Dot1l* HET mice have sex-dependent deficits in sensorimotor function, vocalization development, and sociability.

### Neuronal *Dot1l* loss alters early vocalization development and sociability

Given the behavioral alterations seen in *Dot1l* HETs, we next used a heterozygous forebrain neuron specific *Dot1l* conditional knockout (cKO) mouse to assess whether behavioral alterations in the *Dot1l* HET mice can be attributed to monoallelic loss of *Dot1l* specifically in forebrain neurons. To generate heterozygous *Dot1l* cKO mice, we crossed the floxed *Dot1l* mouse model to mice containing a NEX-cre driver which expresses beginning at approximately embryonic day 11.5 and is specifically expressed in forebrain neurons^60^. We confirmed depletion of *Dot1l* and H3K79me in *Dot1l* cKO cortical tissue as expected (Supplementary Fig. 8A and B). We again recorded USVs in P6 pups and found both male and female *Dot1l* cKO mice had altered frequency of calls in comparison to controls and male *Dot1l* cKO mice had reduced down calls (Fig. 8A-D). This suggests that neuronal expression of *Dot1l* may contribute to early vocal behavior but causes more distinct and more modest changes than those observed in ubiquitous *Dot1l* HET mice. We did not find any developmental delays or weight alterations suggesting effects observed in *Dot1l* HETs are independent of DOT1L function in neurons (Supplementary Fig. 8C-E). Similarly to *Dot1l* HET mice, *Dot1l* cKO had no motor activity or anxiety-related impairments (Fig. 8E, Supplementary Fig. 8F-I). In contrast, female *Dot1l* cKO had increased spontaneous alternations with no change observed in males (Fig. 8F, Supplementary Fig. 8J). Notably, we again found a sex-specific sociability deficit in the three-chamber social test in female *Dot1l* cKO similar to *Dot1l* HETs (Fig. 8G), indicating that DOT1L loss in neurons contributes to this effect. We also found long-term memory deficits in male *Dot1l* cKO mice in contextual fear conditioning (Fig. 8H). While no significant change was observed in females in contextual fear conditioning, freezing rates were low in control female mice, so it is possible that we lacked the dynamic range to detect differences. These data indicate that forebrain neuron-specific monoallelic depletion of *Dot1l* recapitulated sociability deficits in female *Dot1l* HETs and caused changes in vocalization behavior suggesting that expression of *Dot1l* in neurons contributes to specific behavioral alterations.

**Figure 8.**
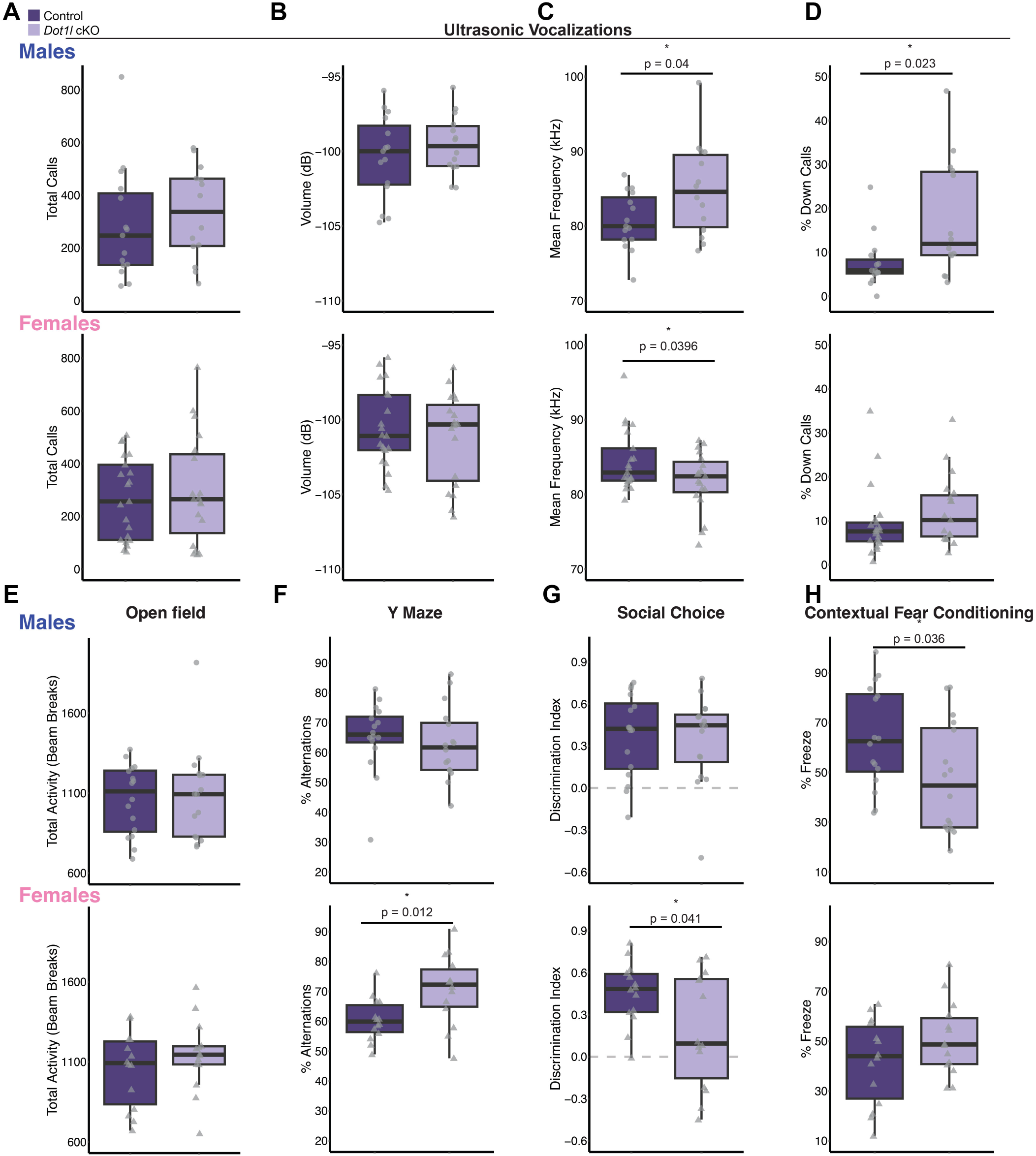
Monoallelic loss of *Dot1l* in forebrain neurons alters language development and sociability. (**A**) Total number of ultrasonic vocalizations (USVs) in P6 male and female pups (male [control: n = 15, *Dot1l* cKO: n = 14]; female [control: n = 21, *Dot1l* cKO: n = 19], Kruskal-Wallis Test). (**B**) Average volume in decibels (dB) of USVs in male and female pups (unpaired two-tailed t-test). (**C**) Average frequency in kilohertz (kHz) of calls in male and female pups (unpaired two-tailed t-test). (**D**) Percent of down calls out of total calls in male pups (Kruskal-Wallis Test). (**E**) Total activity measured from beam breaks during 10-minute open field assay in 4-week-old male and female mice (male [control: n = 16, *Dot1l* cKO: n = 16]; female [control: n = 14, *Dot1l* cKO: n = 14], Kruskal-Wallis Test). (**F**) Percent of spontaneous alternations out of the total number of triads possible in male and female mice (unpaired two-tailed t-test). (**G**) Discrimination index between interaction time with the mouse or rock cylinder during the three-chamber social choice assay in male and female mice (unpaired two-tailed t-test). (**H**) Freezing activity after 2 weeks post-contextual fear conditioning in male and female mice (unpaired two-tailed t-test). All box plot bounds indicate the 25th and 75th percentiles, the black line shows the median, and whiskers extend to the minimum and maximum value that are no further than 1.5X interquartile range. *p<0.05.

## Discussion

Here, we identified 16 individuals with *DOT1L* variants and NDDs, including developmental delays, ASD, and intellectual disability. We confirmed two missense variants disrupt methyltransferase activity of DOT1L which, along with two other variants that result in early stop codons, implicates monoallelic loss-of-function of DOT1L in this disorder. We found widespread transcriptional disruptions upon partial *Dot1l* loss in excitatory neurons both *in vitro* and *in vivo*. Monoallelic loss of *Dot1l* in the brain causes a global reduction of H3K79me2 with the greatest depletion in down-regulated DEGs. Further, *Dot1l* knockdown alters neuron arborization, spine density, expression of synaptic genes, and neuronal activity. In addition, we found that *dot1l* depletion in zebrafish increases activity in response to multiple sensory inputs. Finally, we show that both ubiquitous and neuron-specific monoallelic loss of *Dot1l* cause sex-specific vocalization disruptions and sociability deficits, with additional motor development deficits observed in full-body heterozygous mice. Together, this work demonstrates that partial *Dot1l* loss can lead to an emerging neurodevelopmental disorder and disrupt transcription, neuron morphology, and behavior.

We provide the first functional testing of the p.D157N loss-of-function variant that indicate loss of catalytic activity in *DOT1L* causes neurological dysfunction. Recent work proposed that gain-of-function variants in *DOT1L* contribute to the neurodevelopmental disorders based on modeling in flies and human HEK293T cells.^15^ However, the fly ortholog of *DOT1L*, *grappa*, is highly divergent from human DOT1L (Catalytic domain: 65% identity, Whole gene: 24% identity, 35% similarity, 28% gaps)^42^ and expression of wildtype human DOT1L did not rescue developmental defects caused by *grappa* loss. Our work confirmed the previously described variant p.E123K does indeed function as a gain-of-function. However, we also found that the variant p.R292C, previously proposed to have no effect, decreases DOT1L methyltransferase activity.^15^ This suggests that modeling in flies may not be sufficient to detect loss-of-function variants. Thus, we sought to characterize an additional *DOT1L* variant in a more homologous system. We found that p.D157N missense variant disrupts methyltransferase activity through multiple approaches. Further work characterizing the effects of all missense variants on catalytic activity is necessary to parse whether other detected variants similarly impact catalytic activity. Together with prior work, our findings suggest that both loss- and gain-of-function variants are likely disruptive to neuronal function placing DOT1L in a growing group of epigenetic regulators for which increases or decreases in function can lead to neurodevelopment disorders.^61,62^

These findings point toward the importance of having two functional copies of DOT1L and suggest transcriptional changes in specific neuronal cell types may be responsible for the resulting phenotypes. Given the ample work illustrating the importance of DOT1L in early corticogenesis,^25,28^ it is noteworthy that we did not detect major changes in cortical neuron identity following loss of just a single copy of *Dot1l*. Rather, we found robust changes in transcriptional programs, particularly in excitatory neurons, resulting in downregulation of genes related to synaptic function. In addition, we performed in-depth behavioral characterization of mice with monoallelic *Dot1l* loss in all tissue and in forebrain neurons. The behavioral overlap of sociability deficits suggests that DOT1L loss in neurons is at least partly responsible for behavioral deficits. The altered early vocalization changes were variable between the ubiquitous *Dot1l* HETs and the forebrain specific *Dot1l* cKO however, given the decrease in total calls in ubiquitous *Dot1l* HETs, this may limit the ability to detect more subtle vocal changes such as frequency. Given that we detected sex-specific effects on both gene expression and behavior, our findings indicate that monoallelic *Dot1l* loss has divergent effects based on sex. We identified a subset of genes that have sex-specific expression differences in control mice and are also differentially expressed upon monoallelic *Dot1l* loss in a sex-specific fashion. These baseline sex differences have modest fold changes but may make genes more sensitive to monoallelic *Dot1l* loss. For example, *Adgrb1* has elevated expression in control females and is only downregulated in female *Dot1l* HETs. Interestingly, *Adgrb1* loss in mice has been reported to lead to social deficits^63^. This is particularly intriguing given that our cohort includes more females than males, although whether this trend will be sustained as additional individuals are identified remains to be determined. Together, this work builds upon the previous modeling of *Dot1l* loss in the brain by indicating that partial loss of *Dot1l* is sufficient to cause changes in transcription, neuron maturation, and behavior and identifies transcriptional pathways and cell types that likely contribute to these deficits.

Our work indicates that H3K79me2 decreases upon monoallelic loss of *Dot1l* in the brain. Although we observed a global decrease in H3K79me2, down DEGs identified in single nucleus RNA-seq showed the greatest loss of H3K79me2 in comparison to non-DEGs and up-DEGs. This suggests that the amount of H3K79me2 loss directly linked to transcriptional disruptions. We did not detect differences in H3K79me2 loss in *Dot1l* HETs between sexes suggesting that the transcriptional differences in male and female mice are not mediated by H3K79me2 deposition. Several notable questions remain that will be critical to understanding the role of DOT1L and H3K79me2 in the brain and in neurodevelopmental disorders. It is unclear whether histone variant replacement or demethylase activity is responsible for H3K79me removal in the brain. There is also conflicting evidence on whether methylation of H3K79 is required for DOT1L to fulfill its role in neuronal differentiation.^30–32^ Given that the majority of variants lie in the catalytic domain and that at least two individual variants result in decreased catalytic activity, our work suggests that H3K79me is important in neuron function. However, our findings also allow for a critical function for H3K79me to emerge *after* neuronal differentiation. Notably, several of the phenotypes that we detected in mouse models were evident in neuron-specific monoallelic loss of *Dot1l* suggesting this cell type is particularly sensitive to *Dot1l* dosage. Although we characterized the impact of monoallelic *Dot1l* loss in the brain, whether transcriptional states of other cell types are also impacted in both mouse and human systems will be critical to understanding the role of DOT1L in contributing to developmental disorders. Lastly, predicted loss-of-function alleles for DOT1L are found within gnomAD suggesting highly variable expression, incomplete penetrance, and/or the potential for attenuation of the disorder described through other unknown factors.

In summary, this work examines the impact of partial loss of *Dot1l* spanning from the transcriptional level to the behavioral level. This research provides insights into the effect of variants on DOT1L and the neuronal changes that may contribute to phenotypes observed in DOT1L loss-of-function variant individuals. Further, our findings expand on our understanding of DOT1L by demonstrating that disruption of a single copy of *Dot1l* is sufficient to disrupt neuronal function and contributes to an emerging neurodevelopmental disorder.

## Data availability

RNA-sequencing, single nucleus RNA-sequencing, and ChIP-sequencing data generated in this study can be accessed under the following GEO accession number GSE279978. Variants p.I85M (SCV004169212), p.E134K (SCV004169195), and p.Gln598* (SCV003804054) are available on ClinVar. Raw patient derived sequencing data cannot be made available due to the given consent under which the individuals where recruited. All other data are available in the main text or the supplementary materials or can be made available upon request to corresponding author.

## Supporting information

Supplementary Figures 1-8, methods, raw westerns

## Data Availability

RNA-sequencing, single nucleus RNA-sequencing, and ChIP-sequencing data generated in this study can be accessed under the following GEO accession number GSE279978. Variants p.I85M (SCV004169212), p.E134K (SCV004169195), and p.Gln598+ (SCV003804054) are available on ClinVar. Raw patient derived sequencing data cannot be made available due to the given consent under which the individuals where recruited. All other data are available in the main text or the supplementary materials or can be made available upon request to corresponding author.

https://www.ncbi.nlm.nih.gov/geo/query/acc.cgi?acc=GSE279978

## Acknowledgements

We thank the patients and their families for sharing data and samples. The authors wish to acknowledge the resources of MSSNG (www.mss.ng), Autism Speaks and The Centre for Applied Genomics at The Hospital for Sick Children, Toronto, Canada. We also thank the participating families and clinicians for their time and contributions to this database, as well as the generosity of the donors who supported this program. We thank Zornitsa Pavlova, Tihomir Todorov, Veneta Bojinova for their contributions for the p.N421S variant. We thank Dr. Tanja Vogel for sharing DOT1L;Sun1-sfGFP mouse line and Dr. Andrea Stout for microscopy support. Behavioral procedures were performed at the Neurobehavior Testing Core at the University of Pennsylvania. Single nucleus RNA-sequencing library preparation was performed by the Single Cell Core at Children’s Hospital of Pennsylvania. We thank the Bhoj lab for providing fibroblasts.

## Funding

NIH NINDS grant 1F31NS129242 (MM) NIH NINDS grant NS134755A1 (EK, MM)

NIH/NICHD grant P50 HD105354, Intellectual and Developmental Disabilities Research Center (research support)

NIH NIMH grant 1DP2MH129985 (MB, EK)

NIH NIMH grant R00MH111836 (MB, EK)

NIH Shared Equipment Grant 1S10OD032363 (EK)

Klingenstein-Simons Fellowship from the Esther A. & Joseph Klingenstein Fund (MB, EK)

Simons Foundation (MB, EK)

Alfred P. Sloan Foundation Research Fellowship FG-2020-13529 (MB, EK)

Brain and Behavior Research Foundation NARSAD Young Investigator Award (MB, EK)

SickKids Research Institute (ARD, GC)

Azrieli Precision Child Health Platform (ARD)

NIHR Manchester Biomedical Research Centre NIHR203308 (SB)

MRC Epigenomics of Rare Diseases Node MR/Y008170/1 (SB)

Miguel Servet program from Instituto de Salud Carlos III, Spain CP22/00141 (DNB)

Netherlands Organisation for Scientific Research ZonMw Vidi, grant 09150172110002 (TSB)

EpilepsieNL (TSB) CURE Epilepsy (TSB)

NIH NINDS grant K08NS135125 (PDC)

University of Pennsylvania Autism Spectrum Program of Excellence (ASPE) (JM, PDC)

ANID-Chile Fondecyt grant #1211411 (GMR, VF)

“Joan Oró” of the Secretary of Universities and Research of the Department of Research and Universities of the Government of Catalonia with code 2024 FI-1 00075 (BEA)

European Union (BEA)

## Supplementary material

Supplementary material is available at *Brain* online.

