## Supplementary Figures 1-8, methods, raw westerns for "Loss of DOT1L disrupts neuronal transcription, behavior, and leads to a neurodevelopmental disorder"

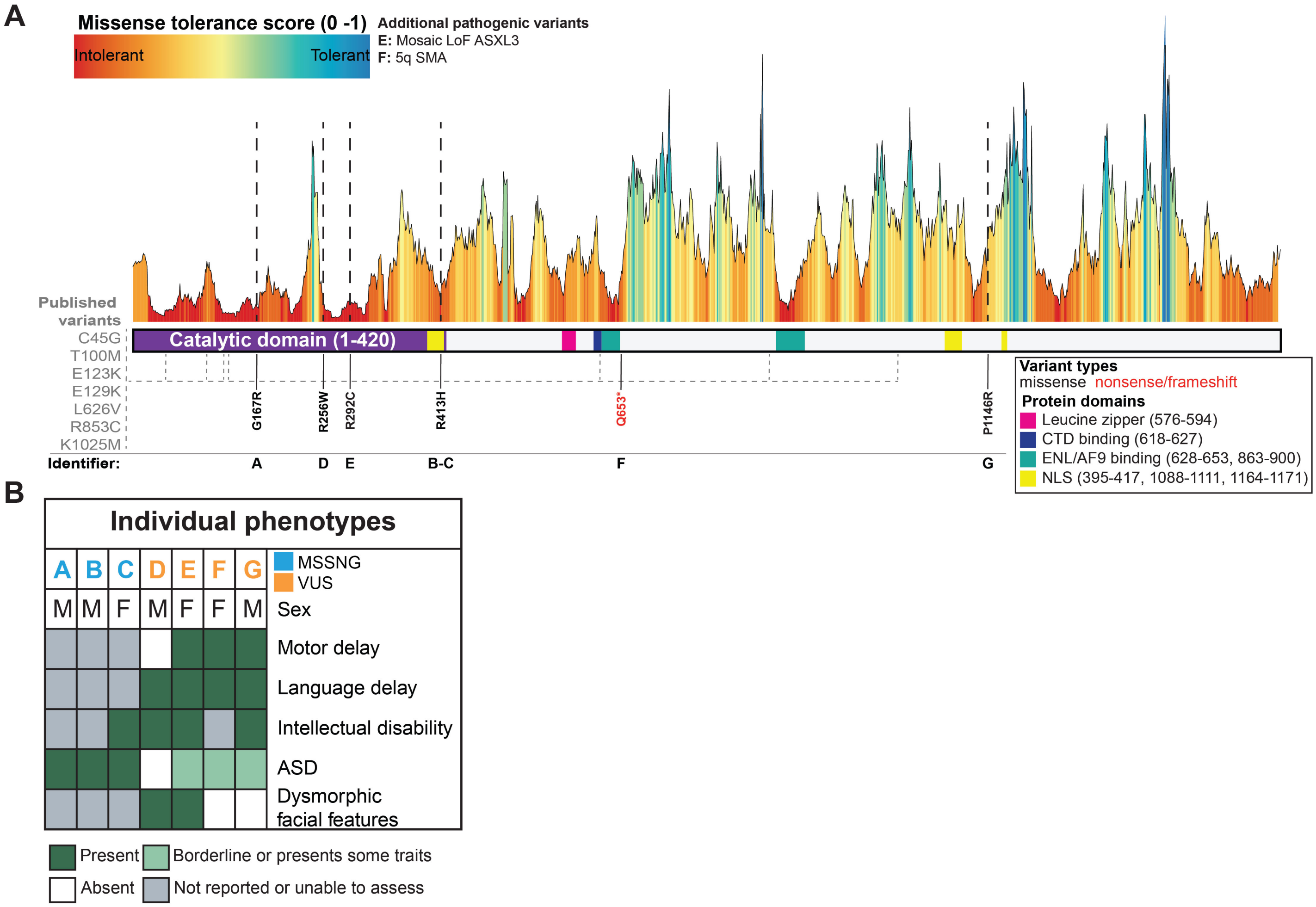


**Supplementary Figure 1.** (A) The landscape of missense tolerance of DOT1L from Metadome. Schematic of DOT1L protein domains (purple = catalytic, pink = leucine zipper, blue = CTD binding, teal = ENL/AF9 binding, and yellow = nuclear localization signal) and locations of DOT1L variants from individuals with ASD from MSSNG database or individuals with variants of uncertain signifcance (VUS) that are in the general population or individuals harboring additional pathogenic variants (black = missense, red = nonsense/frameshift). (B) *DOT1L* variant individuals' phenotypic spectrum and prevalences from MSSNG database (light blue) or individuals with VUS (orange) that are in the general population or individuals harboring additional pathogenic variants. ASD = autism spectrum disorder.

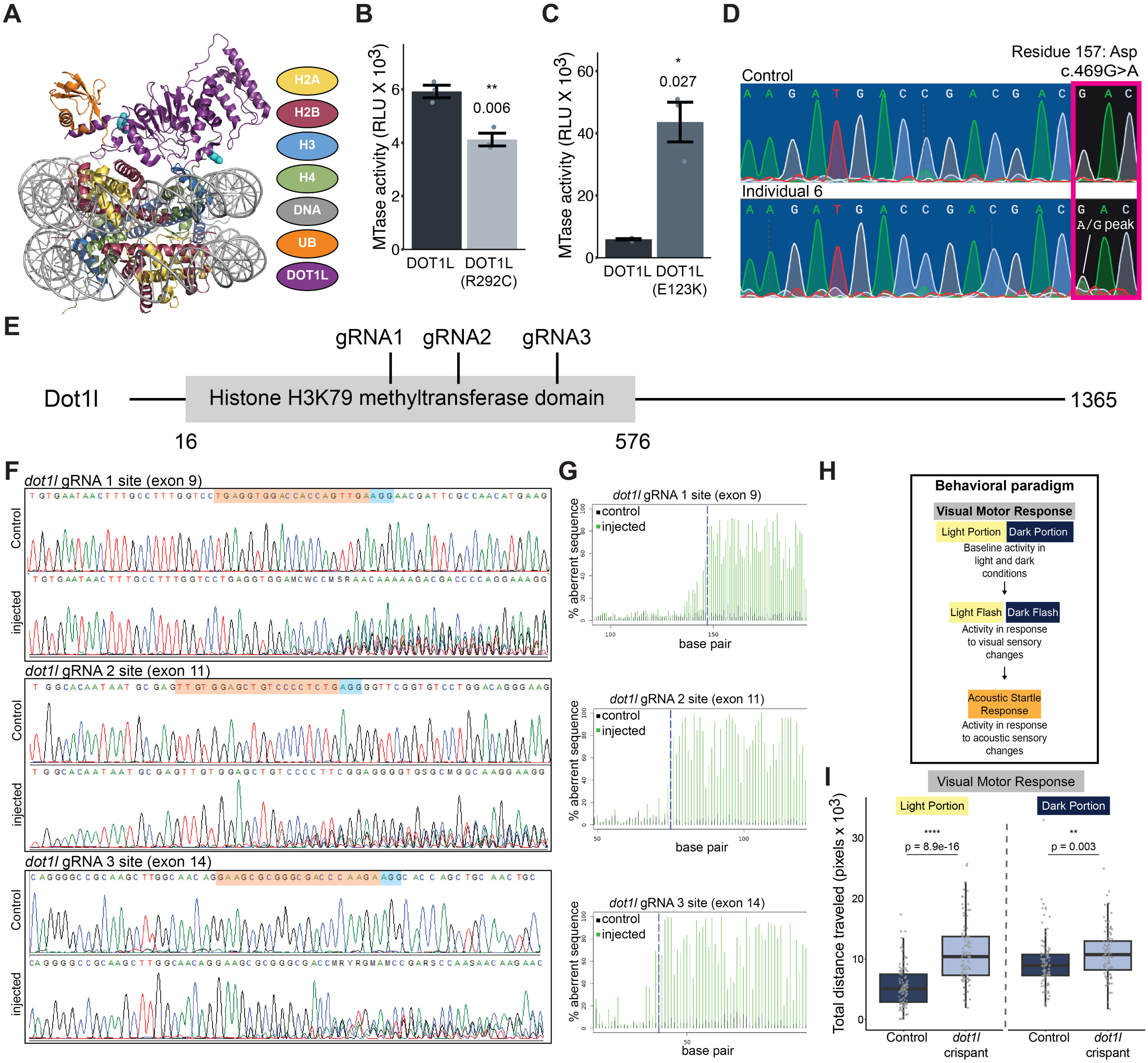


**Supplementary Figure 2.** (A) Structure of the DOT1L nucleosome complex (PDBID: 6NJ9) highlighting the position of previously published variants (variants = cyan spheres, DOT1L = purple, DNA = grey, H2A = yellow, H2B = red, H3 = blue, H4 = dark green, ubiquitin [UB] = orange, and S-adenosylmethionine [SAM] = green sticks). (B-C) Methyltransferase (MTase) activity of human DOT1L or variants of DOT1L, p.R292C (B) or p.E123K (C) on unmodified nucleosomes. Graph shows mean ± SE (n = 3/condition, unpaired two-tailed t-test). RLU = relative light units. (D) Sanger sequencing of human fibroblasts from individual 6 with variant DOT1L (D157N) and from age- and sex-matched control with wildtype DOT1L. (E) Protein structure of zebrafish Dot1l showing location of PANTHER-defined Histone H3K79 methyltransferase domain in relation to the three gRNAs. (F) Sanger sequence traces of PCR-amplified genomic region around each gRNA target site in *dot1l* gRNA-injected and control-injected embryos. *dot1l* gRNA-injected samples initially show correct sequence, which then becomes aberrant at the site of the gRNA target indicating successful mutagenesis. (G) Aberrancy plots for each gRNA. Plots show percentage of peaks for each base pair position that are different from the expected wild-type base pair. *dot1l* gRNA-injected samples initially show % aberrancy similar to control samples followed by a clear increase around and after the gRNA target site (blue dashed line). (H) Schematic behavioral paradigm performed on zebrafish. (I) Total distance traveled with lights on and off during the visual motor response assay in *dot1l* crispant and control zebrafish (control: n = 107, dot1l crispant: n = 89; 3 independent experiments, Kruskal-Wallis Test). All box plot bounds indicate the 25th and 75th percentiles, the black line shows the median, and whiskers extend to the minimum and maximum value that are no further than 1.5X interquartile range. *p<0.05, **p<0.01, ****p<0.0001.

* p = 0.0105 ** p = 0.0037 * p = 0.0348

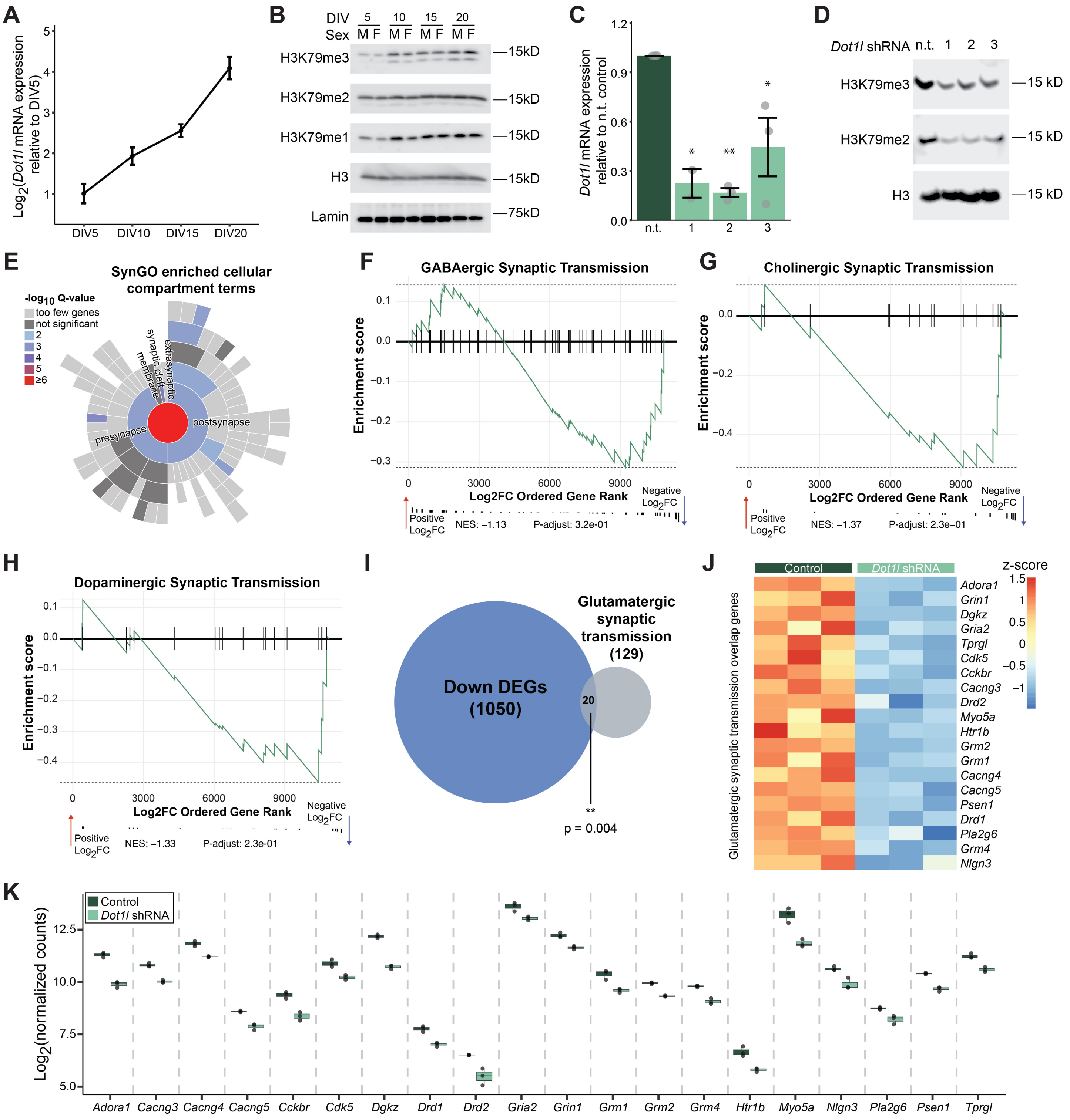


**Supplementary Figure 3.** (A) *Dot1l* mRNA expression in primary cortical neurons from 5-20 days in vitro (DIV) Graph shows mean ± SE. (n=6/DIV). (B) Representative western blot of H3K79me1/2/3 in primary cortical neuron from male (M) and female (F) pups from 5-20 DIV. (C) *Dot1l* mRNA expression in primary cortical neurons infected with 3 different *Dot1l* shRNAs or non-targeting control (n.t.). Graph shows mean ± SE. (n.t.: n = 3, *Dot1l* shRNA 1: n = 2, *Dot1l* shRNA 2: n = 3, *Dot1l* shRNA 3: n = 3, ANOVA with post-hoc pairwise t-test with Bonferroni correction). (D) Representative western blot of H3K79me2/3 in primary cortical neurons infected with *Dot1l* shRNA or n.t. control. (E) SynGO analysis of cellular compartment in down-regulated DEGs. (F-H) Gene set enrichment analysis of genes involved in (F) GABAergic, (G) cholinergic, (H) or dopaminergic synaptic transmission. NES = normalized enrichment score. (I) Overlap of down-regulated DEGs and glutamatergic synaptic transmission genes (hypergeometric test). (J) Heatmap of the 20 glutamatergic synaptic transmission genes that are down-regulated DEGs. (K) Log2(normalized counts) of the 20 glutamatergic synaptic transmission genes that are down-regulated DEGs. *p<0.05, **p<0.01.

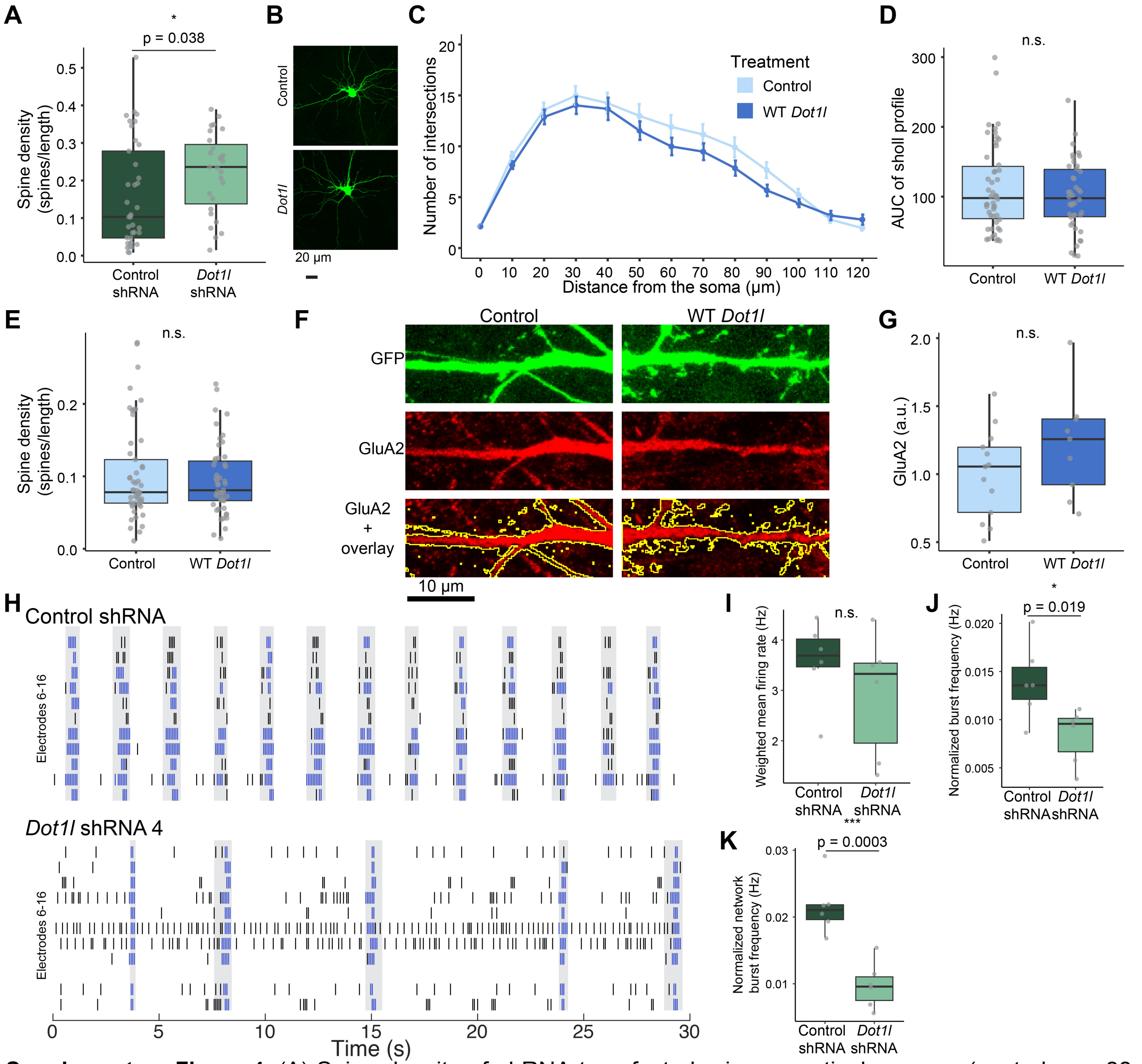

**Supplementary Figure 4.** (A) Spine density of shRNA-transfected primary cortical neurons (control: n = 33 neurons from 5 biological replicates, *Dot1l* shRNA: n = 23 neurons from 5 biological replicates, Kruskal-Wallis Test). (B) Representative images of primary cortical neurons transfected with wildtype (WT) *Dot1l* or empty vector control. Scale bar = 20 µm. (C) Number of branch intersections per radius of WT *Dot1l* or control transfected neurons (control: n = 50 neurons from 8 biological replicates, *Dot1l*: n = 42 neurons from 8 biological replicates). (D) Area under the curve (AUC) quantification of (C) (control: n = 50 neurons from 8 biological replicates, *Dot1l*: n = 42 neurons from 8 biological replicates, mixed effect model). (E) Spine density of WT *Dot1l* or control transfected neurons (control: n = 50 neurons from 8 biological replicates, *Dot1l*: n = 42 neurons from 8 biological replicates, Kruskal-Wallis Test). (F) Representative images of GluA2 staining in WT *Dot1l* or control transfected neurons. (G) Quantification of GluA2 staining in (F) (control: n = 13 neurons from 2 biological replicates, *Dot1l*: n = 9 neurons from 2 biological replicates, Kruskal-Wallis Test). (H) Representative raster plot showing spike activity in *Dot1l* shRNA and control infected neurons (black = spike, blue = burst, gray = network burst). (I-K) Activity metrics in *Dot1l* shRNA and control infected neurons at 20 days in vitro that include (I) weighted mean firing rate (unpaired two-tailed t-test), (J) normalized burst frequency (unpaired two-tailed t-test), and (K) normalized network burst frequency (control: n = 6 wells from 3 biological replicates, *Dot1l* shRNA: n= 6 wells from 3 biological replicates, unpaired two-tailed t-test). All box plot bounds indicate the 25th and 75th percentiles, the black line shows the median, and whiskers extend to the minimum and maximum value that are no further than 1.5X interquartile range. n.s. = not significant. *p<0.05, ***p<0.001.

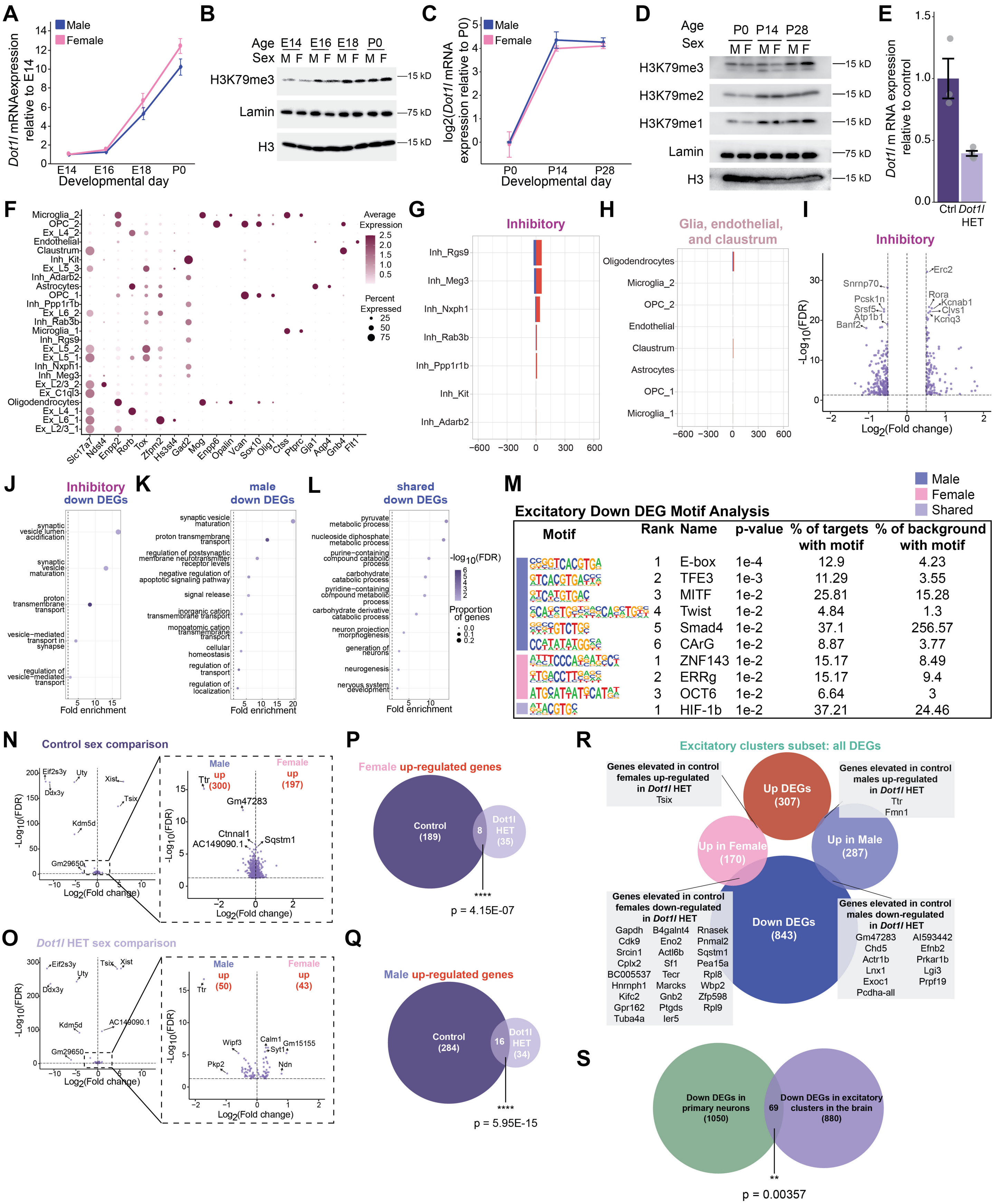
A
Male
Female
Dot1l mRNAexpression relative to E14
E14 E16 E18 P0
Developmental day
B
Age E14 E16 E18 P0
Sex M F M F M F M F
H3K79me3
Lamin
H3
—15 kD
—75 kD
—15 kD
C
Male
Female
log2(Dot1l mRNA expression relative to P0)
P0 P14 P28
Developmental day
D
Age P0 P14 P28
Sex M F M F M F
H3K79me3
H3K79me2
H3K79me1
Lamin
H3
—15 kD
—15 kD
—15 kD
—75 kD
—15 kD
E
Dot1l m RNA expression relative to control
Ctrl Dot1l HET
F
Microglia_2
OPC_2
Ex_L4_2
Endothelial
Claustrum
Inh_Kit
Ex_L5_3
Inh_Adarb2
Astrocytes
OPC_1
Inh_Ppp1r1b
Ex_L6_2
Inh_Rab3b
Microglia_1
Inh_Rgs9
Ex_L5_2
Ex_L5_1
Inh_Nxph1
Inh_Meg3
Ex_L2/3_2
Ex_C1ql3
Oligodendrocytes
Ex_L4_1
Ex_L6_1
Ex_L2/3_1
Slc17a7 Ndst4 Enpp2 Rorb Tox Zfpm2 Hs3st4 Gad2 Mog Enpp6 Opalin Vcan Sox10 Olig1 Ctss Ptprc Gja1 Aqp4 Gnb4 Flt1
Average Expression
2.5
2.0
1.5
1.0
0.5
Percent Expressed
25
50
75
G
Inhibitory
Inh_Rgs9
Inh_Meg3
Inh_Nxph1
Inh_Rab3b
Inh_Ppp1r1b
Inh_Kit
Inh_Adarb2
H
Glia, endothelial, and claustrum
Oligodendrocytes
Microglia_2
OPC_2
Endothelial
Claustrum
Astrocytes
OPC_1
Microglia_1
I
Inhibitory
Erc2
Snrnp70
Pcsk1n
Srsf5
Atp1b1
Banf2
Rora
Kcnab1
Clvs1
Kcnq3
-Log10(FDR)
Log2(Fold change)
J
Inhibitory down DEGs
synaptic vesicle lumen acidification
synaptic vesicle maturation
proton transmembrane transport
vesicle-mediated transport in synapse
regulation of vesicle-mediated transport
Fold enrichment
K
male down DEGs
synaptic vesicle maturation
proton transmembrane transport
regulation of postsynaptic membrane neurotransmitter receptor levels
negative regulation of apoptotic signaling pathway
signal release
inorganic cation transmembrane transport
monoatomic cation transmembrane transport
cellular homeostasis
regulation of transport
regulation of localization
Fold enrichment
L
shared down DEGs
pyruvate metabolic process
nucleoside diphosphate metabolic process
purine-containing compound catabolic process
carbohydrate catabolic process
pyridine-containing compound metabolic process
carbohydrate derivative catabolic process
neuron projection morphogenesis
generation of neurons
neurogenesis
nervous system development
Fold enrichment
-log10(FDR)
Proportion of genes
M
Male
Female
Shared
Excitatory Down DEG Motif Analysis
Motif | Rank | Name | p-value | % of targets with motif | % of background with motif
1 E-box 1e-4 12.9 4.23
2 TFE3 1e-3 11.29 3.55
3 MITF 1e-2 25.81 15.28
4 Twist 1e-2 4.84 1.3
5 Smad4 1e-2 37.1 256.57
6 CArG 1e-2 8.87 3.77
1 ZNF143 1e-2 15.17 8.49
2 ERRg 1e-2 15.17 9.4
3 OCT6 1e-2 6.64 3
1 HIF-1b 1e-2 37.21 24.46
N
Control sex comparison
Eif2s3y
Uty
Ddx3y
Xist
Tsix
Kdm5d
Gm29650
Male up (300)
Female up (197)
Ttr
Gm47283
Ctnnal1
AC149090.1
Sqstm1
-Log10(FDR)
Log2(Fold change)
O
Dot1l HET sex comparison
Eif2s3y
Uty
Ddx3y
Tsix
Xist
Kdm5d
AC149090.1
Gm29650
Male up (50)
Female up (43)
Ttr
Wipf3
Calm1
Gm15155
Syt1
Pkp2
Ndn
-Log10(FDR)
Log2(Fold change)
P
Female up-regulated genes
Control (189)
8
Dot1l HET (35)
****
p = 4.15E-07
Q
Male up-regulated genes
Control (284)
16
Dot1l HET (34)
****
p = 5.95E-15
R
Excitatory clusters subset: all DEGs
Genes elevated in control females up-regulated in Dot1l HET
Tsix
Up DEGs (307)
Genes elevated in control males up-regulated in Dot1l HET
Ttr
Fmn1
Up in Female (170)
Up in Male (287)
Genes elevated in control females down-regulated in Dot1l HET
Gapdh B4galnt4 Rnasek
Cdk9 Eno2 Pnmal2
Srcin1 Actl6b Sqstm1
Cplx2 Sf1 Pea15a
BC005537 Tecr Rpl8
Hnrnph1 Marcks Wbp2
Kifc2 Gnb2 Zfp598
Gpr162 Ptgds Rpl9
Tuba4a Ier5
Down DEGs (843)
Genes elevated in control males down-regulated in Dot1l HET
Gm47283 AI593442
Chd5 Efnb2
Actr1b Prkar1b
Lnx1 Lgi3
Exoc1 Prpf19
Pcdha-all
S
Down DEGs in primary neurons (1050)
69
Down DEGs in excitatory clusters in the brain (880)
**
p = 0.00357

**Supplementary Figure 5.** (A) *Dot1l* mRNA expression in whole cortex from embryonic day 14 (E14) to postnatal day 0 (P0) in male and female mice. Graph shows mean ± SE (E14[male: n = 5, female n= 3], E16[male: n = 4, female: n = 4], E18[male: n = 3, female: n = 6], P0[male: n = 4, female: n = 5]). (B) Representative western of H3K79me3 levels in whole cortex from E14-P0 in male and female mice. (C) *Dot1l* mRNA expression in cortical nuclei from P0-P28 in male and female mice (P0[female: n = 2], all remaining groups n = 3/timepoint/sex). (D) Representative western of H3K79me1/2/3 in cortical nuclei from P0-P28 in male and female mice. (E) *Dot1l* mRNA expression in control and *Dot1l* HET whole cortex. Graph shows mean ± SE (control: n = 3 [1 male, 2 females]; *Dot1l* HET: n = 4 [2 males, 2 females]). (F) Dot plot showing expression of cell type-specific marker genes for each cluster. (G-H) Number of up- and down-regulated differentially expressed genes (DEGs) within (G) inhibitory clusters and (H) glia, endothelial, and claustrum clusters. (I) Volcano plot showing DEGs from *Dot1l* HET and control combined inhibitory clusters. (J) Biological process gene ontology analysis of down-regulated DEGs from combined inhibitory clusters. (K-L) Biological process gene ontology analysis of down-regulated DEGs in (K) only males or (L) shared *Dot1l* HETs from excitatory clusters. (M) Motif enrichment analysis of male-specific, female-specific, or shared down-regulated DEGs. (N) Volcano plot showing DEGs from control male and control female combined excitatory clusters. (O) Volcano plot showing DEGs from *Dot1l* HET male and *Dot1l* HET female combined excitatory clusters. (P) Overlap of genes up-regulated in female controls with genes up-regulated in female *Dot1l* HETs (hypergeometric test). (Q) Overlap of genes up-regulated in male controls with genes up-regulated in male *Dot1l* HETs (hypergeometric test). (R) Overlap of DEGs in all *Dot1l* HET excitatory clusters with genes up-regulated in male or female controls. (S) Overlap of down-regulated DEGs from primary neurons infected with *Dot1l* shRNA and down-regulated DEGs from combined excitatory clusters from *Dot1l* HET cortical nuclei (hypergeometric test). ** $p < 0.01$, **** $p < 0.0001$.

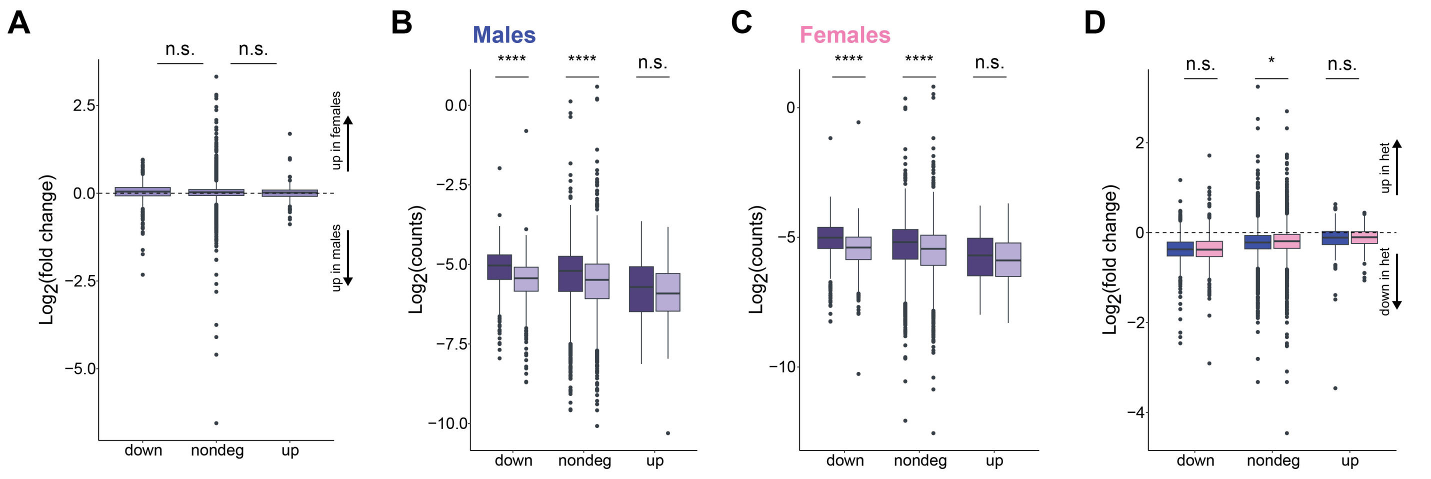


**Supplementary Figure 6.** (A) H3K79me2 log2(fold change) of control female in comparison to control males binned by down DEGs, non-DEGS, and up DEGs (one-way ANOVA with post hoc pairwise t tests with Bonferroni correction). (B) Male *Dot1l* HET and control H3K79me2 log2(counts) binned by down DEGs, non-DEGS, and up DEGs (two-way ANOVA with post hoc Tukey HSD). (C) Female *Dot1l* HET and control H3K79me2 log2(counts) binned by down DEGs, non-DEGS, and up DEGs (two-way ANOVA with post hoc Tukey HSD). (D) H3K79me2 log2(fold change) of male Dot1l HET (blue) or female *Dot1l* HET (pink) in comparison to within sex controls males binned by down DEGs, non-DEGS, and up DEGs (two-way ANOVA with post hoc Tukey HSD). *p<0.05, ****p<0.0001.

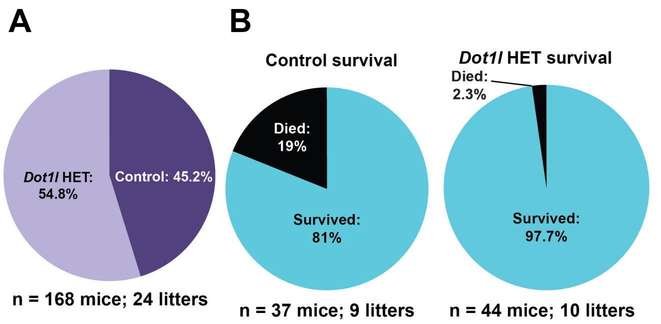


C

| | Males | | Females | |
|---|---|---|---|---|
| Milestone | Day Achieved | | Day Achieved | |
| | Ctrl | Dot1l | Ctrl | Dot1l |
| Eye Opening | 14.1 ± 0.3 | 14.2 ± 0.2 | 14.3 ± 0.2 | 14.2 ± 0.1 |
| Ear Opening | 15.1 ± 0.3 | 15.5 ± 0.4 | 14.8 ± 0.1 | 15 ± 0.2 |
| Fur | 13.3 ± 0.3 | 13.7 ± 0.2 | 13.5 ± 0.2 | 13.3 ± 0.2 |
| Incisor Eruption | 9.5 ± 0.2 | 10.5 ± 0.2 | 9.7 ± 0.3 | 9.5 ± 0.2 |
| Walking | 11.2 ± 0.2 | 11.3 ± 0.2 | 10.4 ± 0.3 | 10.9 ± 0.2 |
| Grasp Reflex | 6 ± 0.3 | 6.4 ± 0.3 | 6.5 ± 0.2 | 5.9 ± 0.2 |
| Cliff avoidance | 9.5 ± 0.5 | 10.5 ± 0.5 | 9.7 ± 0.6 | 9.5 ± 0.5 |
| Place response | 9.5 ± 0.3 * | 10.5 ± 0.3 | 9.7 ± 0.4 | 9.5 ± 0.3 |
| Tactile Startle | 11.2 ± 0.4 | 11.4 ± 0.6 | 11.4 ± 0.6 | 11.8 ± 0.4 |

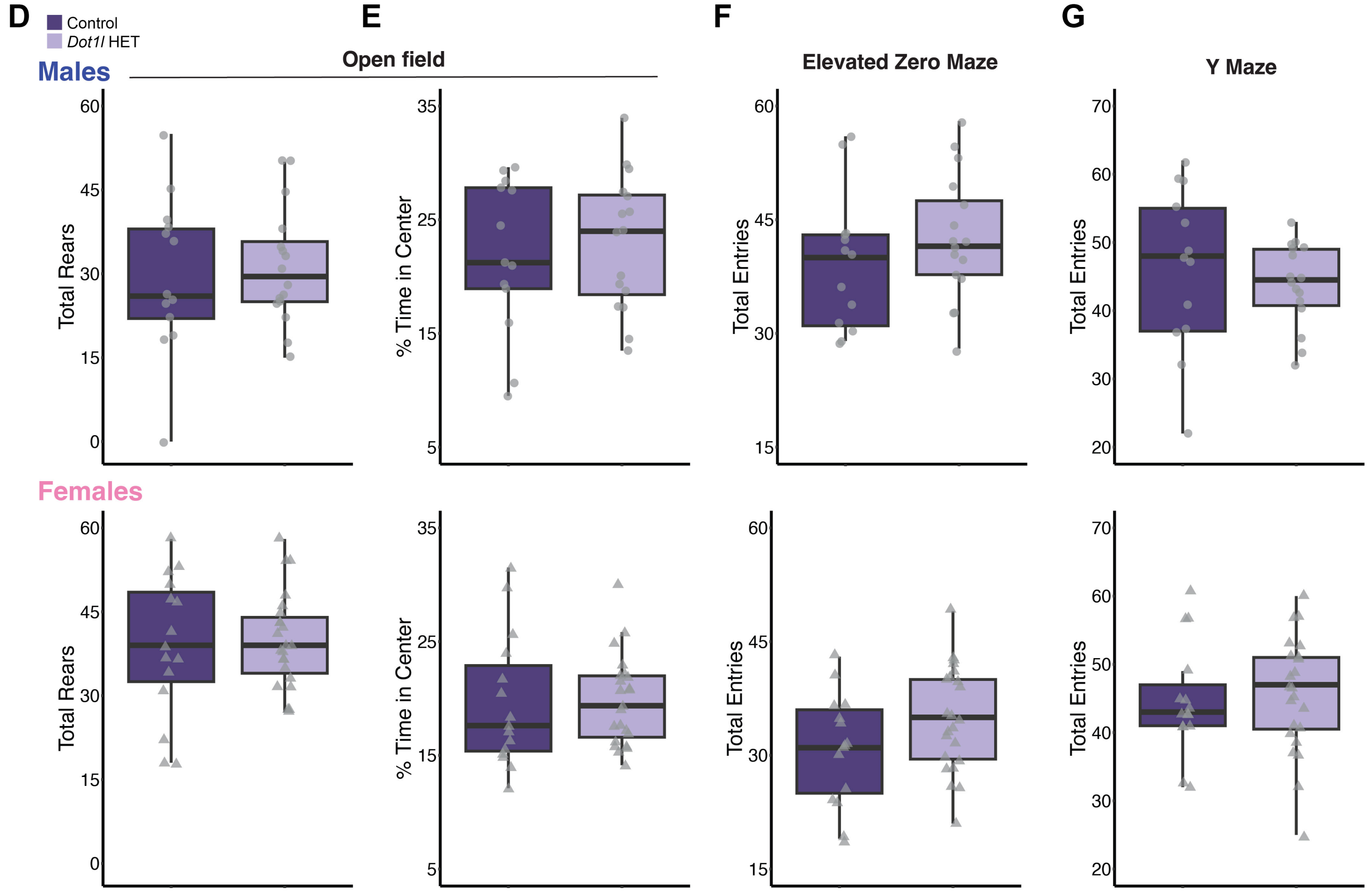


**Supplementary Figure 7.** (A) Pie chart of progeny from Dot1l$^{floxed/+}$ x CMV-Cre$^{+/+}$ crosses (n = 168 mice from 24 litters). (B) Survival rate at weaning age (approximately 4 weeks) in *Dot1l* HET and control mice (control: n = 37 mice from 9 litters, *Dot1l* HET: n = 44 mice from 10 litters). (C) Developmental milestone achievements in male and female mice (male[control: n = 13, *Dot1l* HET: n = 16]; female[control: n = 15, *Dot1l* HET: n = 23], unpaired two-tailed t-test). (D) Total number of rears during the open field assay in male and female mice. (males[control: n = 13, *Dot1l* HET: n = 16]; female[-control: n = 15, *Dot1l* HET: n = 23], unpaired two-tailed t-test). (E) Percent of time spent in the center of the open field arena in male and female mice (Kruskal-Wallis Test). (F) Total entries during the elevated zero maze in male and female mice (unpaired two-tailed t-test). (G) Total arm entries during the Y maze in male and female mice (unpaired two-tailed t-test). All box plot bounds indicate the 25th and 75th percentiles, the black line shows the median, and whiskers extend to the minimum and maximum value that are no further than 1.5X interquartile range.

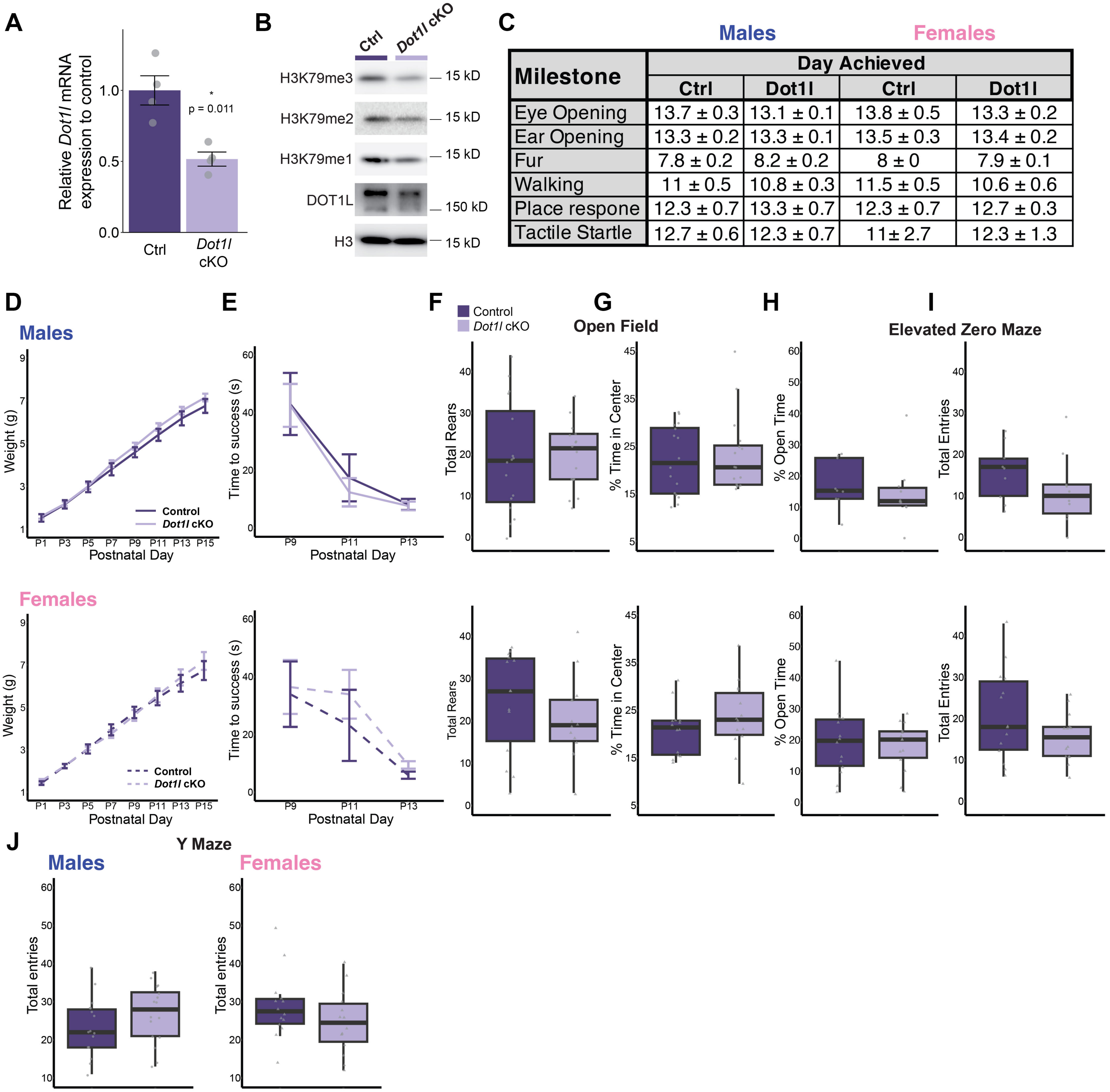


| Milestone | Day Achieved | | | |
|---|---|---|---|---|
| | Ctrl | Dot1l | Ctrl | Dot1l |
| Eye Opening | 13.7 ± 0.3 | 13.1 ± 0.1 | 13.8 ± 0.5 | 13.3 ± 0.2 |
| Ear Opening | 13.3 ± 0.2 | 13.3 ± 0.1 | 13.5 ± 0.3 | 13.4 ± 0.2 |
| Fur | 7.8 ± 0.2 | 8.2 ± 0.2 | 8 ± 0 | 7.9 ± 0.1 |
| Walking | 11 ± 0.5 | 10.8 ± 0.3 | 11.5 ± 0.5 | 10.6 ± 0.6 |
| Place respone | 12.3 ± 0.7 | 13.3 ± 0.7 | 12.3 ± 0.7 | 12.7 ± 0.3 |
| Tactile Startle | 12.7 ± 0.6 | 12.3 ± 0.7 | 11± 2.7 | 12.3 ± 1.3 |

**Supplementary Figure 8.** (A) *Dot1l* mRNA expression in whole cortex from embryonic day 14 (E14) to postnatal day 0 (P0) in male and female mice. Graph shows mean ± SE (E14[male: n = 5, female n= 3], E16[male: n = 4, female: n = 4], E18[male: n = 3, female: n = 6], P0[male: n = 4, female: n = 5]). (B) Representative western of H3K79me3 levels in whole cortex from E14-P0 in male and female mice. (C) Developmental milestone achievements in male and female mice (male[control: n = 6, *Dot1l* cKO: n = 11]; female[control: n = 4, *Dot1l* cKO: n = 9], unpaired two-tailed t-test). (D) Weight during first two weeks post-birth in male and female mice (repeated measures ANOVA). (E) Time to right self in negative geotaxis assay during development in male and female pups (repeated measures ANOVA). (F) Total number of rears during the open field assay in male and female mice (male[control: n = 16, *Dot1l* cKO: n = 16]; female[control: n = 14, *Dot1l* cKO: n = 14], unpaired two-tailed t-test). (G) Percent of time spent in the center of the open field arena in male and female mice (Kruskal-Wallis Test). (H) Percent of time spent in open arms of the elevated zero maze in male and female mice. (male[control: n = 9, *Dot1l* cKO: n = 10; female[control: n = 14, *Dot1l* cKO: n = 14], unpaired two-tailed t-test). (I) Total entries during the elevated zero maze in male and female mice (unpaired two-tailed t-test). (J) Total arm entries during the Y maze in male and female mice (unpaired two-tailed t-test). All box plot bounds indicate the 25th and 75th percentiles, the black line shows the median, and whiskers extend to the minimum and maximum value. * $p < 0.05$

### Materials and methods

#### Study Participants

##### Identification of DOT1L Variants

Variants in DOT1L were identified through connections made through collaborating clinicians, GeneMatcher,[34] Deciphering Developmental Disorders Research Study[6], GeneDx[35], and the MSSNG (www.mss.ng) database[36]. The first individual of interest (Individual E) was identified through a prior publication.[6,12] The remainder of participants were identified through GeneMatcher or GeneDx apart from the following: individual 2, individual 7, individual 11, and individual G (identified through pre-existing collaboration), individual 3, 4, 9 (GeneDx), individuals A-C (MSSNG database). The initial GeneMatcher entry was made on May 23, 2023, and all matches until February 2025 were considered in this study (Supplementary Table 1). Variants are reported according to Human Genome Variation Society[37] nomenclature in reference to the DOT1L transcript (NM_032482.3). Allele counts were gathered from gnomAD (v4.1.0), TOPMed Bravo, and RGC Million Exome Variant Browser (Supplementary Table 2, Supplementary Table 3). Pathogenicity of missense variants was predicted using an aggregation of the following databases: Metadome[38] and AlphaMissense[39] (Supplementary Table 2, Supplementary Table 3). Variants p.I85M (SCV004169212), p.E134K (SCV004169195), and p.Gln598* (SCV003804054) are available on ClinVar.

##### Ethical Statement

Voluntary, informed consent was obtained from human participants and/or family, consistent with the institutional principles of research ethics and the legal requirements of each referring author's jurisdiction. Ethical approvals were obtained for participation, phenotyping, sample collection and generation/derivation of affected individual and control fibroblasts (MEC-2012-387, Erasmus MC institutional review board; IRB#16-013278_AM118, Children's Hospital of Philadelphia). The authors also confirm that human research participants and/or family provided written informed consent for publication of the images in Fig. 1.

### Methyltransferase Activity

#### Expression and Purification of DOT1L and Mutants

DOT1L and mutants were expressed and purified as previously described.[40] Briefly, the proteins were expressed in BL21 One Shot (DE3) (ThermoFisher) E. coli cells. They were grown at 37°C until reaching an OD600 equal to 0.6-0.8 and then were induced using 0.5 mM IPTG for 3 hours at 37°C. The cells were harvested (Sorvall LYNX6000) and then lysed (AvestinEmulsiflexC3) (Lysis buffer: 500 mM NaCl, 50 mM Tris-HCl pH 8.0, 5% Glycerol, 5 mM Imidazole, 2 mM BME, 1x Protease Inhibitor). Lysate was incubated with Ni-NTA Beads (Qiagen). Protein was eluted (Elution buffer: 500 mM NaCl, 50 mM Tris-HCl pH 8.0, 5% Glycerol, 300 mM Imidazole, 2 mM BME) and cleaved by TEV protease overnight in dialysis (Dialysis buffer: 75 mM NaCl, 20 mM Tris pH 8.0, 5% Glycerol, 2 mM BME). Sample was then purified over a HiTrap SP HP column (Cytvia) (Buffer A: 75 mM NaCl, 25 mM HEPES pH 7.5, 5% Glycerol, 2 mM BME) and eluted with a linear salt gradient (75 mM to 1000 mM NaCl); and then further purified over HiLoad Superdex 200 16/600 size exclusion column (GE Healthcare) (150 mM NaCl, 10 mM HEPES pH 7.5, 2 mM DTT). Protein was concentrated, then flash frozen in liquid nitrogen and stored in a -80°C freezer.

#### Purification of Widom 601 DNA

Widom 601 DNA was transformed into DH5a competent E. Coli cells (NEB) from a plasmid containing 8 copies of 147 bp repeats flanked by EcoRV sites.[41] The cells were grown overnight at 37°C, then harvested and lysed. The DNA was then further purified using established protocols.[42]

#### Expression and Purification of Xenopus Histones

Xenopus laevis histones H2A, H2B, H3 and H4 were expressed and purified using previously published protocols.[42] Briefly, the histone constructs were cloned in a pET-3 vector and grown in pLysS (DE3) cells (NEB) to an OD600 of 0.6-0.8 and induced at 0.5 mM IPTG at 37°C for 3 hours. The protein was then extracted from inclusion bodies and purified over a size exclusion column Sephacryl S200 (Cytvia) followed by an SP anion exchange column (Tosoh). Proteins were then dialyzed in 1mM BME and then lyophilized using a Vertis Sentry lyophilizer.

##### Reconstitution of Nucleosomes

Unmodified nucleosome was assembled as previously described.[41,42] First equimolar ratios of unfolded histones H2A, H2B, H3, and H4 were mixed and dialyzed in refolding buffer. The assembled octamer was then purified over a size exclusion chromatography column Superdex 200 26/600(GE Healthcare) using refolding buffer. Nucleosomes were assembled by combining an equimolar quantity of octamer and Widom 601 DNA, followed by an overnight salt gradient dialysis using a peristaltic pump (Gilson).

##### Endpoint Methylation Assay

The endpoint methylation assay was performed as described.[40] Assays on DOT1L and mutants with unmodified nucleosome were done in three replicates. Briefly, in methyltransferase buffer, 250 nM and 125 nM of DOT1L or mutants were combined with 1 µM of nucleosome. In a volume of 20 ml, the reaction was incubated at 30°C for 30 minutes (min). The reaction was stopped using 5ml of 0.5% TFA. SAH production was determined using a MTase-Glo methyltransferase kit (Promega). The luminescence was measured using an EnSpire 2300 Multilabel plate reader (Perkin Elmer).

#### Western blotting

Protein lysates or histone samples were mixed with 5X Loading Buffer (5% SDS, 0.3M Tris pH 6.8, 1.1mM Bromophenol blue, 37.5% glycerol), boiled for 10 min, and cooled on ice. Protein was resolved by 4%–20% Tris-glycine or 16% Tris-glycine SDS-PAGE, followed by transfer to a 0.45-µm PVDF membrane for immunoblotting. Membranes were blocked for 1 hour at RT in 5% milk in 0.1% TBST and probed with primary antibody overnight at 4°C (Supplementary Table 4). Membranes were incubated with secondary antibody for 1 hour at RT.

### Cell culture

#### Human Fibroblasts

Individual 6 fibroblasts and age- and sex-matched control fibroblasts were donated from collaborating clinicians. Fibroblasts were cultured in DMEM (with 4.5 g $L^{-1}$ glucose, L-glutamine and sodium pyruvate) supplemented with 15% FBS (Sigma-Aldrich, F2442-500ML) and 1% penicillin-streptomycin (Gibco, 15140122).

#### Neuro-2A cells

Neuro-2A cells were obtained from the American Type Culture Collection (ATCC), cultured in DMEM (with 4.5 g $L^{-1}$ glucose, L-glutamine and sodium pyruvate) supplemented with 10% FBS (Sigma-Aldrich, F2442-500ML) and 1% penicillin-streptomycin (Gibco, 15140122) and maintained free of mycoplasma. N2A transfections were performed in DMEM using lipofectamine 2000 (Life Technologies, 11668027). Lipofectamine and DNA complexes were left on for overnight. Cells were harvested for analysis 2 days after transfection.

#### Primary neuronal culture

Cortices were dissected from E16.5 C57BL/6J embryos and cultured in neurobasal medium (Gibco 21103049) supplemented with B27 (Gibco 17504044), GlutaMAX (Gibco 35050061), penicillin-streptomycin (Gibco 15140122) in TC-treated twelve or six-well plates coated with 0.05 mg/mL Poly-D-lysine (Sigma-Aldrich A-003-E). At 3 DIV, neurons were treated with 0.5 µM AraC. Transfections were performed using lipofectamine 2000 (Life Technologies, 11668027). Neurons were put in a 1 mM kynurenic acid solution during transfection to prevent excitotoxicity. Lipofectamine and DNA complexes were left on neurons for 15 min. Transfections were performed at 8 to 12 DIV for constructs expressing DOT1L and cells were fixed two to three days later. shRNA transfections were performed at 9 to 12 DIV and fixed three to four days later. Neuronal infections were transduced overnight with lentivirus containing the constructs described below. Virus was removed the following day, and neurons were cultured for 5-7 days.

#### Constructs

The GFP control plasmid was obtained from Addgene, pLenO-CMV-MCS-GFP-SV-puro (Addgene plasmid# 73582). The pET28-MHL-DOT1L (1-420) was received from the Armache lab originally purchased from Addgene (Addgene plasmid# 40736). The pET28-MHL-DOT1L contains only the catalytic domains of human DOT1L and was utilized in Fig. 2E and Supplementary Fig. 2B-C. The pDSV-DOT1l-HA-Flag-mRFP-nls and empty pDSV-mRFP-nls plasmids were received from the Vogel lab. The pDSV-DOT1l-HA-Flag-mRFP-nls contains the full-length coding sequence of mouse *Dot1l* and was utilized in Fig. 2G and Supplementary Fig. 4B-G. The Sun1-GFP plasmid was a gift from Jeremy Nathans Lab, pCDNA3-CMV-Sun1-GFP-6xMyc. *Dot1l* shRNA and control Luciferase shRNAs were inserted into the pLKO.1 vector backbone (Addgene plasmid# 10878). *Dot1l* shRNA target sequences were as follows:

1. *Dot1l* shRNA 1:

   CCGGGTCCAGTTTGTACTGTCAATACTCGAGTATTGACAGTACAAACTGGACTTTTTG

2. *Dot1l* shRNA 2:

   CCGGCCTCGGTTTACACAGCTTCAACTCGAGTTGAAGCTGTGTAAACCGAGGTTTTTG

3. *Dot1l* shRNA 3:

   CCGGCGGCAGAATCGTATCCTCAAACTCGAGTTTGAGGATACGATTCTGCCGTTTTTG

DOT1L mutants were generated using sited directed mutagenesis using Pfu Turbo HotStart DNA polymerase (Agilent, 600322, for p.R292C and p.E123K) or NEB Q5 Polymerase (M0491S, for p.D157N), and primers were created using the DNA-based primer design feature of the online PrimerX tool or manually creating primers using ~15 bp overlap strategy. Plasmid sequences were verified through Sanger sequencing and/or Plasmidsaurus long read sequencing.

##### Lentiviral production

HEK293T cells were cultured in high-glucose DMEM growth medium (with 4.5 g $L^{-1}$ glucose, L-glutamine and sodium pyruvate), 10% FBS (Sigma-Aldrich F2442-500ML), and 1% penicillin-streptomycin (Gibco 15140122). Calcium phosphate transfection was performed with Pax2 and VSVG packaging plasmids. Viral media was removed 2 hours after transfection and collected at 48 and 72 hours later. Viral media was passed through a 0.45-μM filter and precipitated for 48 hours with PEG-it solution (40% PEG-8000 [Sigma-Aldrich P2139-1KG], 1.2 M NaCl [Fisher Chemical S271-1]). Viral particles were pelleted and resuspended in 200 μL PBS.

#### Zebrafish experiments

Experiments were conducted on 6 dpf larval zebrafish (Danio rerio, TLF strain) raised in E3 medium at 29 °C on a 14:10 h light cycle. At this developmental stage the sex of the organism is not yet determined. Breeding adult zebrafish were maintained at 28 °C on a 14:10 h light cycle. Crispants were generated as described[43] by Kroll et al. Three gRNAs targeting three different regions across the dot1l locus were designed using ChopChop v3 (https://chopchop.cbu.uib.no/). Custom Alt-R CRISPR-Cas9 crRNAs (IDT) were annealed with tracrRNA (IDT, #1072533) to form gRNAs which were subsequently complexed with Cas9 protein (IDT, #1081061) to make the final ribonucleoprotein (RNP) complex. Three non-targeting crRNAs (IDT, #1072544, 1072545, 1072546) were used to make the RNP for controls. Single-cell wildtype (TLF) zebrafish embryos were then microinjected within 15 min of fertilization with 1nl of RNP mix containing 357 pg (10.1 fmol) of each gRNA and 5029pg (30.5 fmol) of Cas9. Embryos displaying acute toxicity or damage from microinjection were removed from analysis. The remaining embryos were raised to 6dpf at which point they were arrayed on a 100-well plate and multiple sensorimotor behaviors including the visual motor response, responsiveness to flashes of light or darkness, and the acoustic startle response were assessed as described previously[44]. To confirm that each of the three gRNA-Cas9 RNP complexes was able to target the predicted dot1l locus and cause mutations, genomic DNA was also extracted from dot1l crispants at 6 dpf. The predicted target sites were amplified by PCR using primers that flank the region, and the PCR product was then sent for Sanger sequencing. Each of the three RNPs caused mutations at the predicted target site that were not present in control injected embryos. The targeting gRNA sequences are included in Supplementary Table 5. All experiments were conducted in accordance with and approval of the IACUC at the University of Pennsylvania.

### RNA-sequencing

#### Library preparation & sequencing

RNA was isolated using Zymo Quick-RNA Miniprep Plus Kit (R1057). Libraries were generated using the Illumina TruSeq stranded mRNA library prep kit (Illumina 20020595). Prior to sequencing libraries were quantified by qPCR using a KAPA Library Quantification Kit (Roche 07960140001). Libraries were sequenced on an Illumina NextSeq 500/550; reads (75-bp read length, single end). Data can be accessed under the following GEO accession number: GSE279978.

#### Data processing and analysis

Reads were mapped to Mus musculus genome build mm10 with Star (v2.7.9a). The R packages DESeq2[45] (v1.34.0) and limma (v3.50.3) via edgeR (v3.36.0) were used to perform differential gene expression analysis. We defined genes as differentially expressed where FDR < 0.05 and an absolute log2 fold change > 0.5. Volcano plots were generated using Enhanced Volcano. IGV tools[46] (v2.12.3) was used to generate genome browser views.

#### Gene ontology

PANTHER[47,48] (v18.0) was used to perform an overrepresentation test against the biological process complete ontology using default parameters. SynGO[49] was used for synaptic gene ontologies and overrepresentation tests of differentially expressed genes. All expressed genes (defined as any gene that did not have an NA p adjusted value in the DeSeq2 output and did not have an NA gene name) was used as a background gene list.

#### Revigo

Revigo[50] was used to remove redundant terms and gather a concise list based on a published protocol.[51] In brief, the Panther output of the Biological Process gene ontology terms and their associated FDR-corrected p-values were input into Revigo. Revigo input parameters used were: size of resulting list – small; remove obsolete GO terms – yes; species – Mus Musculus; semantic similarity measure – Resnik. Revigo output was then filtered using the following conditions: reference genes within a gene ontology term <= 3000, dispensability < 0.2, and fold enrichment > 1. The resulting top 10 gene ontology terms based on FDR-corrected p-values were displayed.

#### GSEA

The R package FGSEA[52] was used to perform pre-ranked gene set enrichment analysis (GSEA) based on log2 fold changes obtained from DESeq2 differential expression analysis. Genes without a defined adjusted p-value and genes with a base mean < 100 were removed prior to running GSEA. The GSEA[53,54] database (https://www.gsea-msigdb.org/gsea/index.jsp) was used for synaptic transmission based gene sets.

#### Immunocytochemistry

GluA2 antibody (Synaptic Systems: 182103) was added to the media of live cells and incubated for 45 min. Cells were fixed in 4% PFA for 10 min and washed with PBS. Cells were blocked in blocking solution (PBS with 3% BSA and 2% serum) for at least 1 hour. Cell coverslips were then incubated with secondary antibody for 1 hour at room temperature. For detection of GluA2, Goat anti-Rabbit Alexa Fluor™ 647 (Thermo Fisher, A-21244; 1:500) was added to the secondary antibody solution. Nuclei were stained with DAPI (1:1,000 in PBS) for 10 min with washing in PBS. Coverslips were mounted onto microscope slides using ProLong Gold antifade reagent (Thermo Fisher).

### Image acquisition

Cells were imaged on an upright Leica DM 6000, TCS SP8 laser scanning confocal microscope with 405-nm, 488-nm, 552-nm and 638-nm lasers. The microscope uses two HyD detectors and three PMT detectors. The objective used was a ×63 HC PL APO CS2 oil objective with an NA of 1.40. Type F immersion liquid (Leica) was used for oil objectives. Images were 175.91 × 171.91 $\mu M^2$, 1,024 × 1,024 pixels and 16 bits per pixel. Coverslips were imaged with a *z* stack through the neuron.

### Image analysis

#### Sholl, Spine, and GluA2 Analysis

Images were analyzed using ImageJ (v2.14.0/1.54f) software. A singular z stack image's maximum projection of the GFP channel was generated. The image was traced in Simple Neurite Tracer (SNT)[55] and the Sholl analysis feature[56] was used to generate a data table with number of intersections per step size (radius step size = 10 µm). An R script (https://zenodo.org/records/1158612) was used generate graphs and summary statistics using a mixed effect model[57]. Spine density was quantified from the maximum projection image. The three largest neurite branches were measured and projections from these branches were counted. Spines had to be > 0.4 µm and < 8 µm in order to be counted based off previous literature.[58,59] Spine density for each branch was calculated as number of spines/branch length and this value was averaged together for each neuron imaged. To quantify GluA2 levels, an in-house macro was created. In short, this macro creates an outline of the imaged neuron from each individual stack using the GFP channel and then measures the fluorescent intensity in that stack from the far-red channel used to stain GluA2. The fluorescent intensity of each image is normalized to the average intensity of the control transfected neurons.

### Multi-electrode array (MEA)

Neurons were plated on CytoView MEA 48-well plates (Axion BioSystems M768-tMEA-48W). Prior to plating, plates were coated with 50 ug/mL poly-D-lysine (Sigma-Aldrich A-003-E) in borate buffer pH 8.4, incubated overnight at 37°C, washed 4X with H2O, and air dried overnight. After air drying, wells were coated with 20 ug/mL laminin (Roche 11243217001) in ice-cold Opti-MEM (Gibco 51985091) and incubated 4 hours at 37°C. Immediately prior to seeding, laminin was removed from wells and 80K neurons were seeded on the CytoView plate. Recordings were performed using an Axion Maestro Pro™ multiwell microelectrode array with 5% CO2 at 37°C. Baseline recordings were taken after 20 days in vitro. Neurons acclimated to the apparatus for 30 min before recording 5 min of neuronal activity. AxIS software (Axion Biosystems) was used for the extraction of spikes and bursts. Burst activity was defined as a minimum of 5 spikes with a maximum inter-spike interval of 100 ms.

#### Single nuclei RNA-sequencing (snRNAseq)

##### Nuclei Isolation

For each biological replicate, one cortical hemisphere of a mouse was dissected, and flash frozen in liquid nitrogen and stored at −80 °C. The nuclei isolation procedure used was modified from.[60,61] Tissue was homogenized in douncers using a loose pestle (~10-15 strokes) in 1.2 mL of homogenization buffer supplemented with 1 mM DTT, 0.15 mM spermine, 0.5 mM spermidine, RNasin ® Plus Ribonuclease Inhibitor (Promega N2611), and EDTA-free protease inhibitor (Roche). A 5% IGEPAL-630 solution was added (107 µl), and the homogenate was further homogenized with the tight pestle (~10-15 strokes). The sample was then mixed with 1.3 mL of 50% iodixanol density medium (Sigma D1556) and added to a polypropylene thin wall tube (13.2 mL, Beckman and Coulter, 331372). The sample was then underlaid with a gradient of 30% and 40% iodixanol, and centrifuged at 10,000 x g for 18 min (no brake) in a swinging bucket centrifuge at 4 °C. Nuclei from control and *Dot1l* HET mice were individually counted (3 males, 3 females per genotype) and proportionally combined with all other biological replicates within each genotype. These samples were washed 3 times in DPBS and spun at 1000g for 5 min. Samples were resuspended in 1X Nuclei Buffer (10x Genomics PN-2000153 / 2000207) at a concentration of approximately 5,000 nuclei/µl for subsequent library preparation. All steps were performed on ice or at 4°C.

#### Library preparation & sequencing

For the generation of ATAC and Gene Expression libraries, the 10X Genomics Chromium Next GEM Single Cell Multiome ATAC + Gene Expression (CG000338 Rev F) protocol was followed. Briefly, 16,100 nuclei from each sample underwent a transposition reaction before being loaded on the 10X genomics Chromium controller to target 10,000 recovered nuclei per sample. The resulting barcoded transposed DNA and barcoded cDNA were then used to generate ATAC and gene expression libraries, respectively, following the manufacturer's guidelines. Quality control was performed during library preparation using an Agilent Bioanalyzer and a Thermo Fisher Qubit. Prior to sequencing libraries were quantified by qPCR using a KAPA Library Quantification Kit (Roche 07960140001). Libraries were sequenced on an Illumina NextSeq 1000, using 28 cycles for Read 1, 10 cycles for the i7 index, 10 cycles for the i5 index, and 90 cycles for Read 2. Data can be accessed under the following GEO accession number: GSE279978.

#### Preprocessing of snRNAseq data

Paired end sequencing reads were processed using 10X Genomics Cellranger v5.0.1. Reads were aligned to the mm10 genome optimized for single cell sequencing through a hybrid intronic read recovery approach.[62] In short, reads with valid barcodes were trimmed by TSO sequence, and aligned using STAR v2.7.1 with MAPQ adjustment. Intronic reads were removed, and high-confidence mapped reads were filtered for multimapping and UMI correction. Empty GEMs were also removed as part of the pipeline. Initial dimensionality reduction and clustering was performed prior to processing to enable batch correction and removal of cell free mRNA using SoupX.[63] Raw expression matrices with counted, individual nuclei UMI and genes were used for subsequent steps and filtering by QC metrics.

#### Clustering and merging by genotype and comparison

Raw matrices for each individual genotype were converted to Seurat objects using Seurat 5.0.1 and filtered to remove UMIs with thresholds of > 200 minimum features, < 5% mitochondrial reads, and < 5% ribosomal reads. Each genotype (control and *Dot1l* HET, each containing 6 biological replicates) were merged to generate an object for the subsequent steps. Each dataset was normalized (NormalizeData) using the default scale factor of 10000, variable selection (FindVariableFeatures) was performed using 2000 features, then scaled and centered (ScaleData) using all features without regressing any variables. Dimensionality reduction with PCA (RunPCA) used the first 30 principal components and the nearest-neighbor graph construction (FindNeighbors) used the first 10 dimensions. Clustering (FindClusters) was next performed using a resolution of 0.5 before layers corresponding to each genotype were integrated (IntegrateLayers) using CCA Integration with a k weight of 60 and then rejoined (JoinLayers). The dataset per condition was then dimensionally reduced using the integrated CCA at with 30 dimensions (RunUMAP) and the same resolution of 0.5.

#### Marker gene identification

To identify marker genes for each cluster, differential expression analysis was performed using the Seurat function FindAllMarkers. Differentially expressed genes that were expressed at least in 25% cells within the cluster and with a fold change more than 0.5 (log scale) were considered marker genes. Cell identity was determined using well-established marker genes for major cortical cell types. Marker gene analysis led to the identification of 17 cortical neuron clusters (10 excitatory, 7 inhibitory), 1 subcortical neuron cluster, and 7 non-neuronal clusters. Neuronal clusters were annotated according to the cortical layer they occupy, or—if unidentifiable by cortical layer—according to the gene most differentially expressed in that cluster relative to all other excitatory or inhibitory neuronal clusters.

#### Differential gene expression analysis and parsing sex of nuclei

Differential gene expression analysis between control and *Dot1l* HET groups was performed using the Seurat function FindMarkers (min.pct = 0.001, logfc.threshold = 0.5) with a MAST test. Genes with an adjusted p-value $< 0.05$ and an absolute log2 fold change $> 0.5$ were considered differentially expressed between control and *Dot1l* HET. The sex of the nuclei was determined based on the following parameters: females were categorized as nuclei with Xist expression at or above the 70$^{th}$ percentile in comparison to total nuclei and with no expression of Eif2s3y or Ddx3y;

males were categorized as nuclei with Xist expression below 70$^{th}$ percentile in comparison to total nuclei and non-zero expression of Eif2s3y or Ddx3y.

##### Gene ontology

PANTHER[47,48] (v19.0) was used to perform an overrepresentation test against the biological process complete ontology using default parameters. All expressed genes (defined as any gene within the current Seurat object subset with a min.pct = 0.001) was used as a background gene list.

##### Revigo

Revigo[50] was used to remove redundant terms and gather a concise list based on a published protocol[51]. In brief, the Panther output of the Biological Process gene ontology terms and their associated FDR-corrected p-values were input into Revigo. Revigo input parameters used were: size of resulting list – small; remove obsolete GO terms – yes; species – Mus Musculus; semantic similarity measure – Resnik. Revigo output was then filtered using the following conditions: reference genes within a gene ontology term <= 2000 and >= 25, dispensability < 0.5, and fold enrichment > 2. The resulting top 10 gene ontology terms based on FDR-corrected p-values were displayed.

#### ChIP-sequencing

##### Chromatin immunoprecipitation

Cortical nuclei were isolated from 10 week old Dot1l HET and control mice as described in the nuclei isolation section (2 males and 2 females/genotype). Approximately 6.9 million cortical nuclei per biological replicate were used as input. After isolation, cells were fixed with 1% PFA in PBS for 15 min at RT and the reaction was quenched with 2.5M glycine. Nuclei were washed twice with ice cold PBS, and then pelleted at 1,500 g for 5 min at 4°C. Nuclei were then resuspended in lysis buffer 2 (10 mM Tris-HCl pH 8, 200 mM NaCl, 1 mM EDTA and 0.5 mM EGTA) to lyse nuclei. Samples were rotated for 10 min at room temperature and were spun again at 1,500 g for 5 min at 4°C. The supernatant was discarded, and the pellet was resuspended in lysis buffer 3 (10 mM Tris-HCl pH 8, 100 mM NaCl, 1 mM EDTA, 0.5 mM EGTA, 0.1% sodium deoxycholate and 0.5% N-lauroylsarcosine). Lysates were sonicated using a Covaris S220 Focused-ultrasonicator for 1 hour (peak power: 140; duty factor: 5.0; 200 cycles per burst; avg power: 7.0). Triton X-100 was added to reach a final concentration of 1%, and lysates were spun at 18,000 g for 10 min at 4°C. 50 µl of lysate was saved as input shearing control.

Antibody-conjugated Protein A Dynabeads (15µg of antibody conjugated to 75 µl of Protein A Dynabeads, resuspended in 50 µl per immunoprecipitation) were added to the lysates overnight with rotation at 4°C. Beads were then washed 8X with RIPA wash buffer (50 mM HEPES-KOH pH 7.5, 500 mM LiCl, 1 mM EDTA, 1% NP-40 and 0.7% sodium deoxycholate) and 1X with TE + 50 mM NaCl. Chromatin was eluted from beads for 30 min with shaking at 65°C in elution buffer (50 mM Tris-HCl pH 8.0, 10 mM EDTA and 1% SDS). Samples were removed from beads and cross-linking was reversed in both input and IP samples by incubating overnight at 65 °C. RNA was digested with 5ug/mL RNase for 1 hr at 37 °C, and protein was digested with 0.2ug/mL proteinase K for 1 hr at 55 °C. DNA was then purified with the Zymo DNA Clean & Concentrator Kit with a final elution in 50µl molecular biology grade H2O.

#### Library preparation & sequencing

Sequencing libraries were prepared using the TruSeq ChIP Library Preparation Kit (Illumina 15023092). Prior to sequencing, library size distribution was confirmed by capillary electrophoresis using an Agilent 4200 TapeStation with high sensitivity D1000 reagents (5067-5585), and libraries were quantified by qPCR using a KAPA Library Quantification Kit (Roche 07960140001). Libraries were sequenced on an Illumina NextSeq1000 instrument (61-bp read length, paired end).

#### Data processing and analysis

Reads were mapped to Mus musculus genome build mm10 with Bowtie 2 (v2.4.5). Eighteen million reads from each biological replicate were subset and each condition was then merged across biological replicates (SAMtools56 v1.15). Metaplots were generated using ngs.plot58 (v2.63) against the mouse genome. Read counts were generated using deepTools.[64]

#### Mice

A floxed *Dot1l* mouse line crossed with the Sun1-sfGFP line (JAX Strain #:030952) was received from Tanja Vogel. In brief, the floxed *Dot1l* mouse line is floxed at exon 2 causing a frameshift that results in an early stop codon and nonfunctional gene product (C57BL6/J background). The *Dot1l* mouse line was originally obtained from the Knockout Mouse Project (KOMP). Heterozygous floxed *Dot1l* mice (*Dot1l*$^{floxed/+}$;Sun1-sfGFP$^{+/+}$) were crossed to the NEX-Cre line[65] for neuron-specific behavioral testing. *Dot1l* cKO were *Dot1l*$^{floxed/+}$;Sun1-sfGFP$^{+/-}$;NEX-Cre$^{+/-}$ and controls were *Dot1l*$^{+/+}$;Sun1-sfGFP$^{+/-}$;NEX-Cre$^{+/-}$. Heterozygous *Dot1l* mice that did not also harbor alleles from the Sun1-sfGFP line (*Dot1l*$^{floxed/+}$; Sun1-sfGFP$^{-/-}$) were crossed to the CMV-Cre line (JAX Strain #:006054) for ubiquitous monoallelic *Dot1l* loss behavioral testing. *Dot1l* HET were *Dot1l*$^{floxed/+}$; CMV-Cre$^{+/-}$ and controls were *Dot1l*$^{+/+}$;CMV-Cre$^{+/-}$. All mice were housed in a 12-hour light-dark cycle and fed a standard diet. All experiments were conducted in accordance with and approval of the IACUC at the University of Pennsylvania.

#### Behavioral assays

##### Behavioral cohorts

Male and female controls (*Dot1l*$^{+/+}$; Sun1-sfGFP$^{+/-}$;NEX-Cre$^{+/-}$ or *Dot1l*$^{+/+}$;CMV-Cre$^{+/-}$), *Dot1l* HET (*Dot1l*$^{floxed/+}$;CMV-Cre$^{+/-}$), and *Dot1l* cKO (*Dot1l*$^{floxed/+}$;Sun1-sfGFP$^{+/-}$;NEX-Cre$^{+/}$) mice were tested in the behavioral tests described below:

For control and *Dot1l* HET mice, two cohorts were generated a month apart from each other and used for developmental milestone testing from P1 – P18. Additionally, these mice were used at 4 weeks old at the onset of behavioral testing which included: elevated zero maze, open field, Y maze, social choice, and fear conditioning, in that order. The breakdown of these cohorts was as follows: Cohort 1[control: male = 4, female = 10, *Dot1l* HET: male = 2, female = 10], Cohort 2 control: male = 9, female = 5, *Dot1l* HET: male = 14, female = 13]. A third cohort of mice were used for ultrasonic vocalizations at P6 – P7. The breakdown was as follows: *Dot1l* HET cohort[ litter = 13, control: male = 15, female = 22, *Dot1l* HET: male = 22, female = 22].

For control and *Dot1l* cKO mice, a cohort was generated for developmental milestone testing from P1 – P18 [control: male = 6, female = 4, *Dot1l* cKO: male = 11, female = 9]. A separate cohort [control: male = 16, female = 14, *Dot1l* cKO: male = 16, female = 14] of mice was used at 4 weeks old at the onset of behavioral testing which included: open field, Y maze, social choice, and fear conditioning, in that order. A third cohort of mice was used at four weeks old at the onset of behavioral testing for elevated zero maze [control: male = 9, female = 14, *Dot1l* cKO: male = 10, female = 14]. A fourth cohort of mice used for ultrasonic vocalizations at P6 – P7 [litter = 10, control: male = 15, female = 21, *Dot1l cKO* male = 14, female = 19]. For all behavioral testing, the experimenter was blinded to genotype of the mice.

#### Ultrasonic vocalizations

Multiple litters were used for both the *Dot1l* HET and *Dot1l* cKO cohorts. Pups at approximately P6 – P7 were individually placed into a soundproof chamber with fresh bedding. A Condenser ultrasound microphone (Avisoft-Bioacoustics CM16/CMPA, part #40011) microphone and UltraSoundGate 116H (Avisoft Bioacoustics, part # 41163, 41164) recording device was used with Avisoft-RECORDER USGH software. The recording sessions were 5 min in length and recorded with the following parameters: sampling rate = 375000 Hz, range = 15 – 180 kHz, and min whistle duration = 5 ms. USVs were analyzed using a MATLAB based software, VocalMat.[66] Mice with fewer than 50 calls were excluded.

#### Elevated zero maze

The elevated zero apparatus consists of a circular shaped platform raised approximately 16 inches above the floor. Two opposing quadrants have raised walls (wall height = 4 inches, circle width = 2 inches) without a ceiling leaving these closed quadrants open to overhead light. The two remaining opposing quadrants were open (wall height = 0.25 inches). Mice were placed into a closed quadrant and allowed to freely explore for 5 min. The entire testing session was recorded, and videos were analyzed using ANY-maze software.

#### Open field

Mice were placed into an empty arena (15 inches x 15 inches) and allowed to freely explore for 10 min. Activity was measured using beam breaks recorded using Photobeam Activity System Open Field software (San Diego Instruments) and percent of center activity was quantified as number of beam breaks in the center / total beam breaks *100.

#### 3-chamber social choice assay

The social choice test was carried out in a three-chambered apparatus, consisting of a center chamber and two outer chambers. Before the start of the test and in a counter-balanced manner, one end chamber was designated the social chamber, into which a stimulus mouse would be introduced, and the other end chamber was designed the nonsocial chamber. Two identical, clear Plexiglas cylinders with multiple holes to allow for air exchange were placed in each end chamber. In the habituation phase of the test, the experimental mouse freely explores the three chambers with empty cue cylinders in place for 10 min. Immediately following habituation, an age- and sex-matched stimulus mouse was placed in the cylinder in the social chamber while a rock was simultaneously placed into the other cylinder in the nonsocial chamber. The experimental mouse was tracked during the 10 min habituation and 10 min social choice phases. All testing was recorded, and videos were analyzed manually.

#### Y maze

The Y maze test was performed on a Y shaped apparatus composed of 3 enclosed arms equidistant apart (3 in wide x 5 in wall x 15 in long). Mice were handled for 2 min each on 3 consecutive days immediately prior to the onset of testing. For Y maze testing mice were placed at the distal end of the arm closest to the experimenter of the Y maze apparatus. Mice were allowed to freely explore for 8 min. Entries into each arm were defined as all four paws of the mouse entering. A spontaneous alternation was defined as a consecutive entry into each of the 3 arms without returning to the arm that the mouse had been in immediately prior. Spontaneous alternation triads over the total number of possible triads based on the total entries were calculated as spontaneous alternations/ (total entries - 2). All testing was recorded, and videos were analyzed manually.

#### Contextual and cued fear conditioning

Mice were handled for 2 min each the day immediately prior to the onset of testing. On training day, mice were placed in individual chambers for 2 min followed by a loud tone lasting 30 second that co-terminated with a 2 second, 1.25-mA foot shock. One minute later mice received another tone-shock pairing and were then left undisturbed for an additional 1 minute in the chamber before being returned to their home cage. Freezing behavior, defined as no movement except for respiration, was determined before and after the tone-shock pairings and scored by MedAssociates VideoFreeze software. To test for context-dependent learning, we placed mice back into the same testing boxes 24 hours later for a total of 5 min without any tone or shock, and again measured the total time spent freezing. Following an additional 24 hours, we tested for cue-dependent fear memory by placing the mice into a novel chamber consisting of altered flooring, wall-panel inserts, and vanilla scent. After 2 min in the chamber, the cue tone was played for a total of 3 min, and the total time spent freezing during the presentation of this cue tone was recorded. Long-term contextual and cued fear memory were again tested with the same protocol at 14 days (contextual) or 15 days (cued) post-training.

### Statistical analysis

All statistical analyses were performed using readily available code in R. Number of replicates and details of statistical tests are reported in figure legends. Shapiro-Wilk's method was used to test for normality of a given dataset. Detailed information on statistical tests as well as all relevant test statistics can be found in Supplementary Table 6.

Figure 2F blot

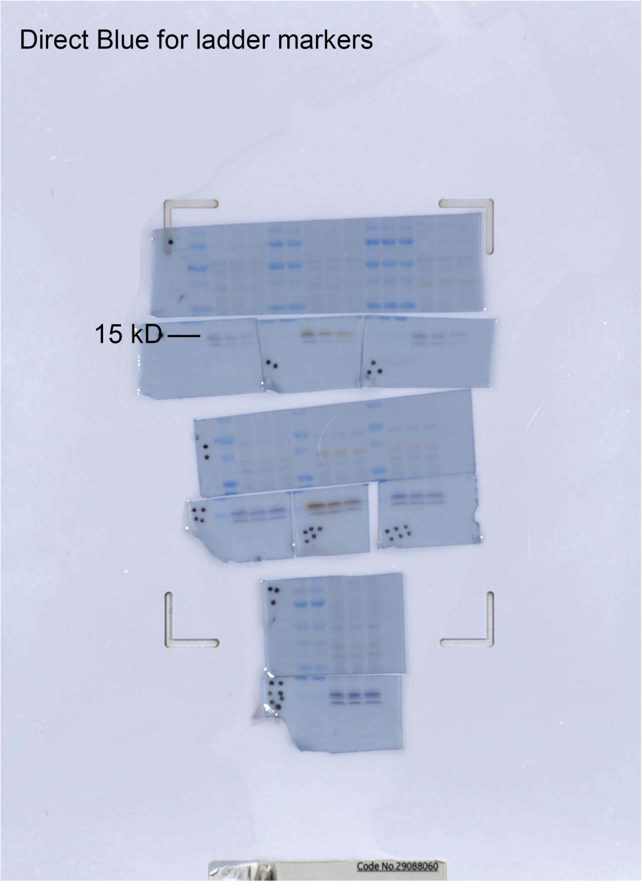


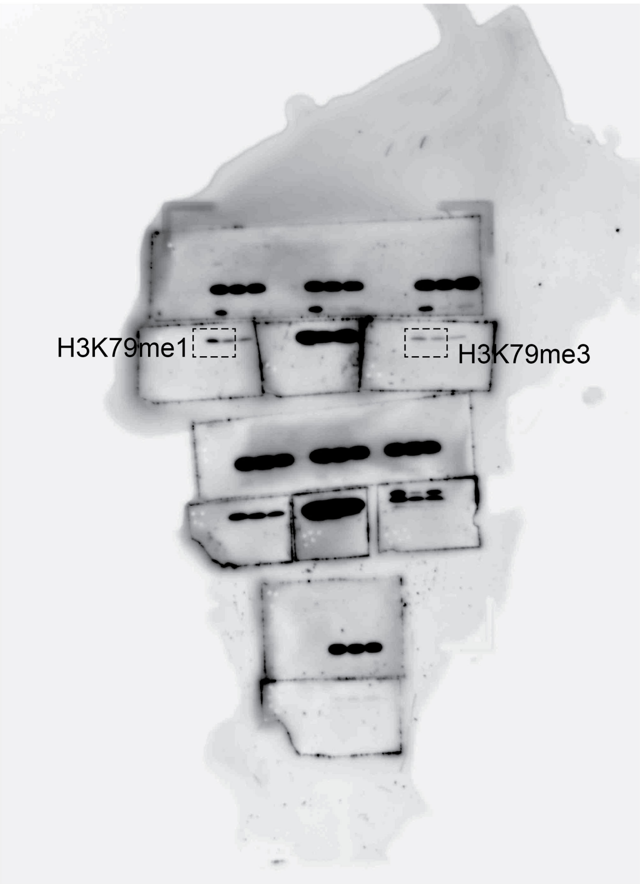


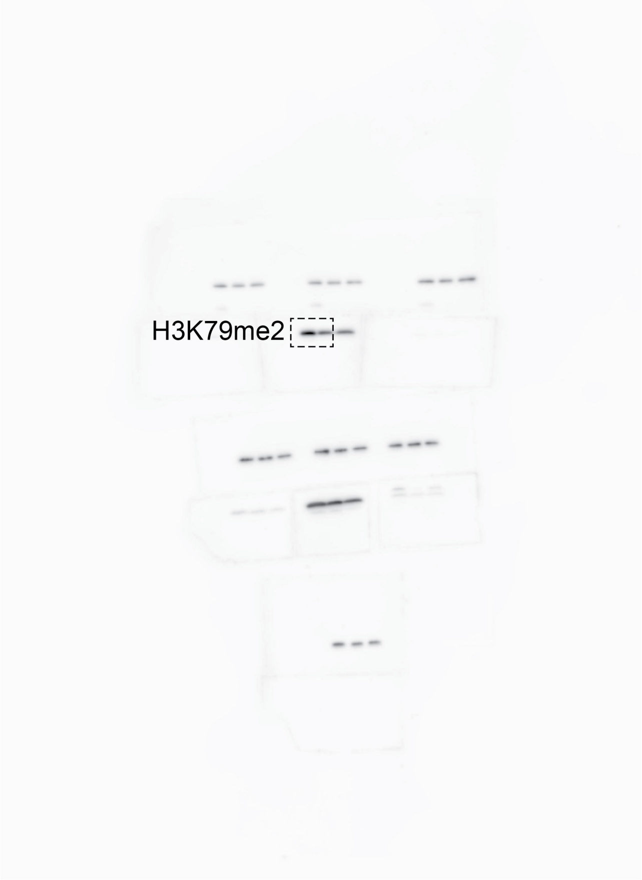

Figure 2F blot

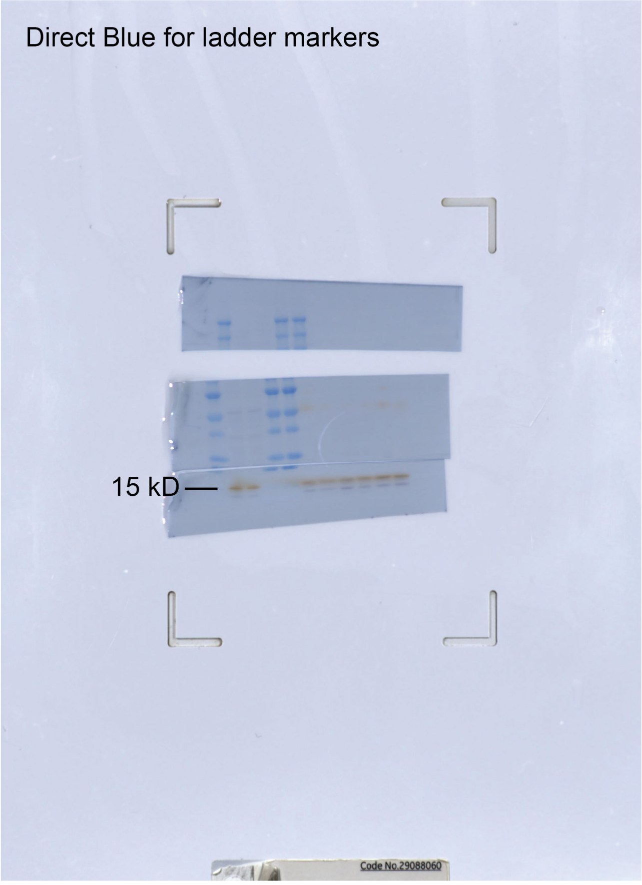


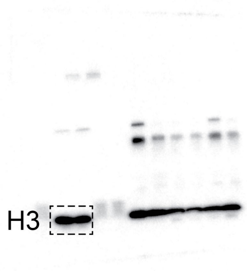

Figure 2G blot

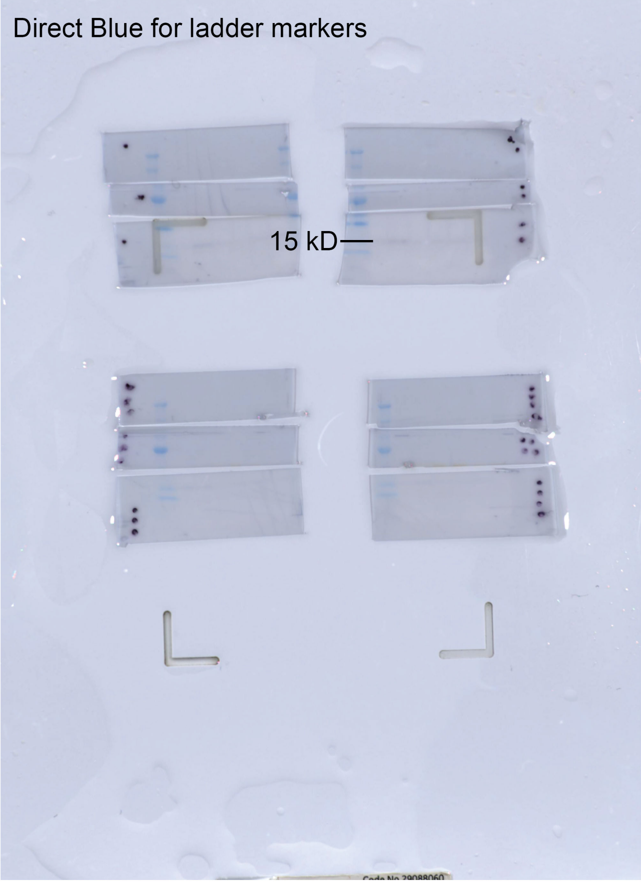


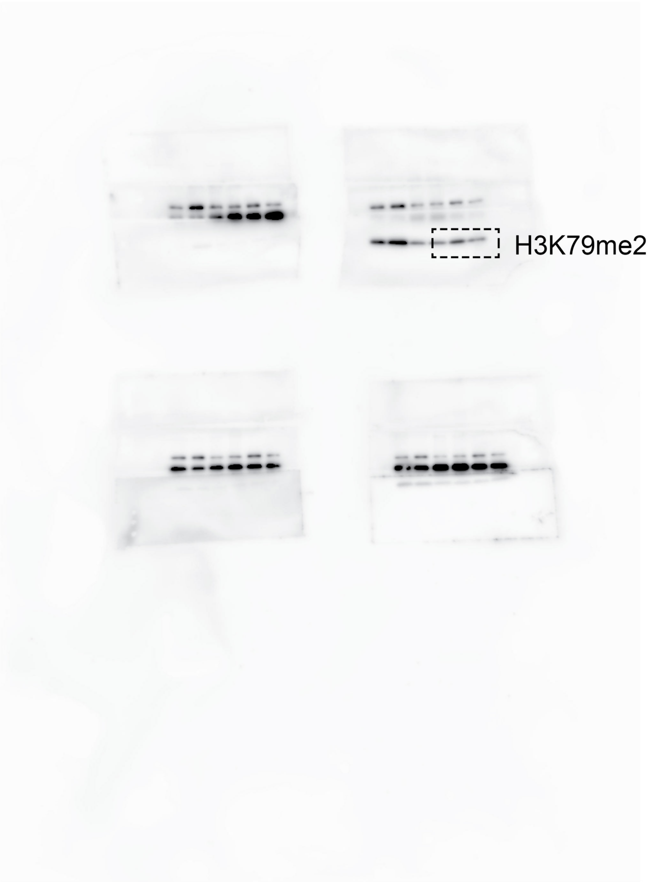


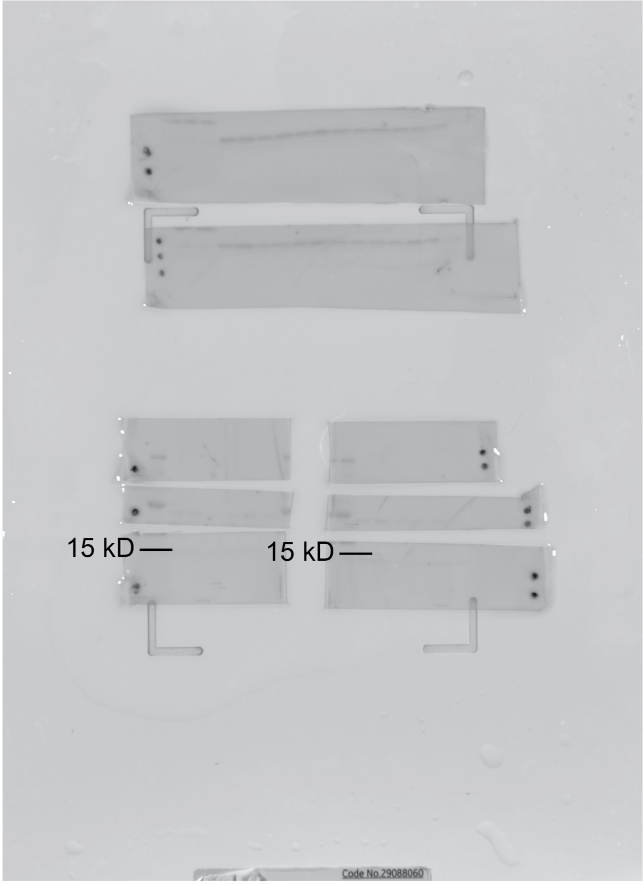


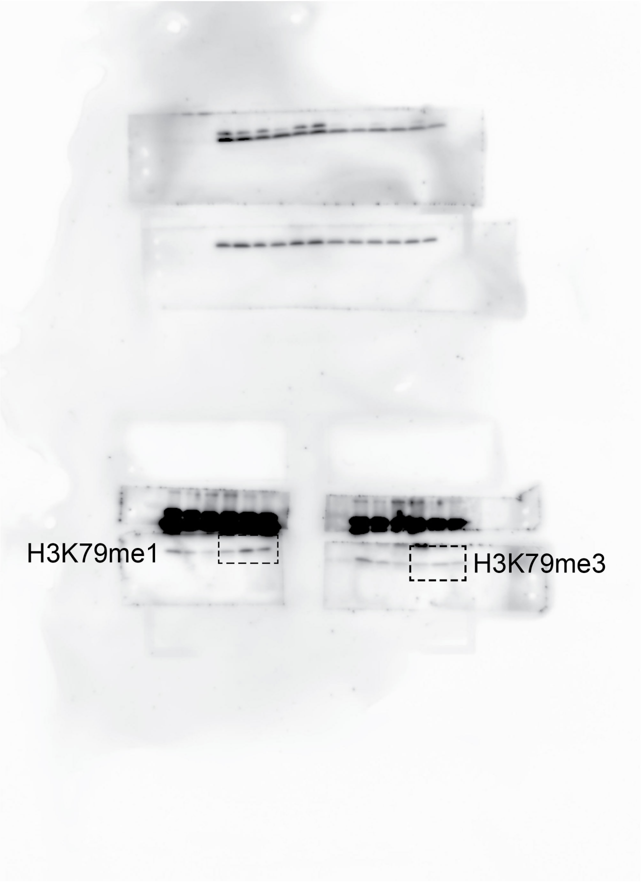

### Figure 2G blot

Direct Blue for ladder markers

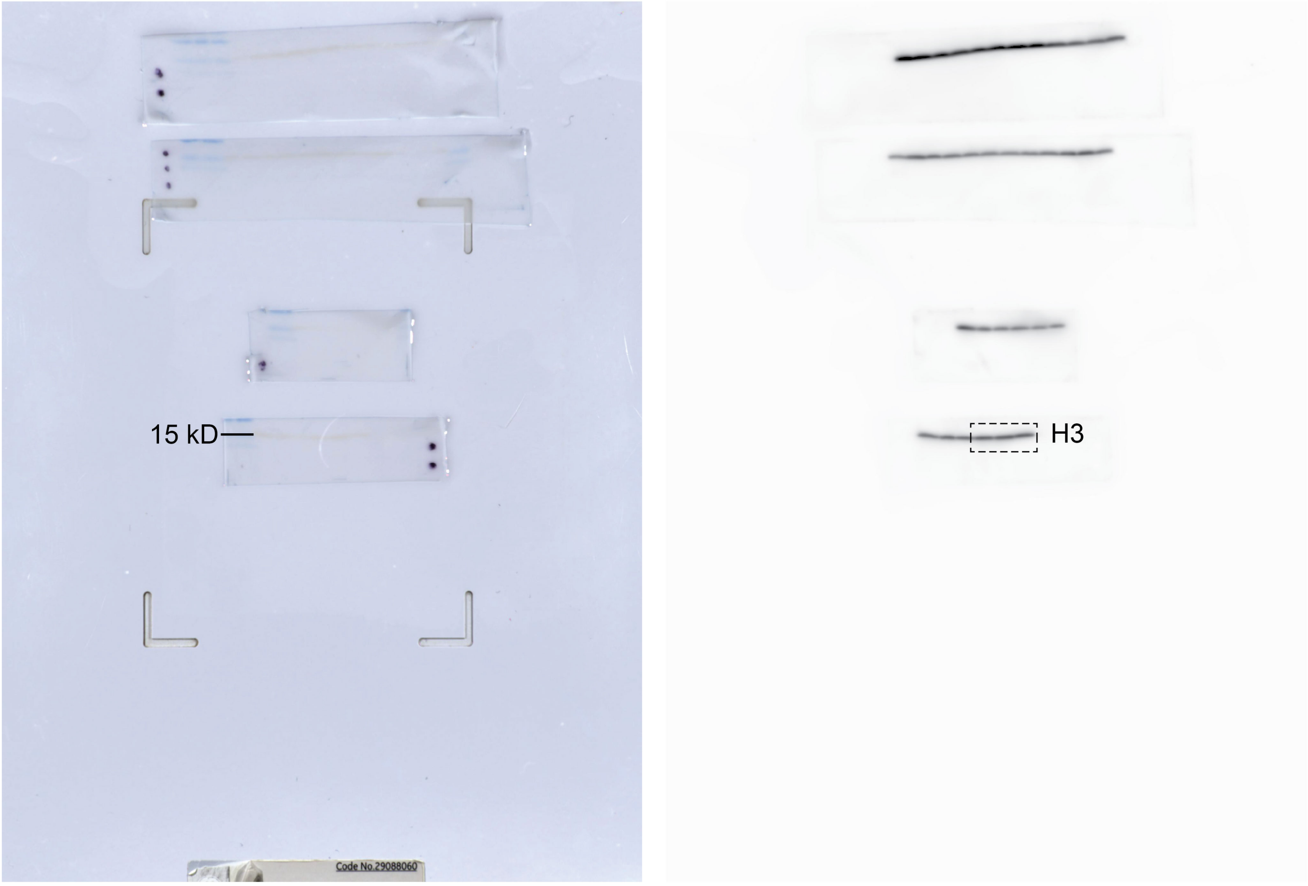

Figure S3B blot

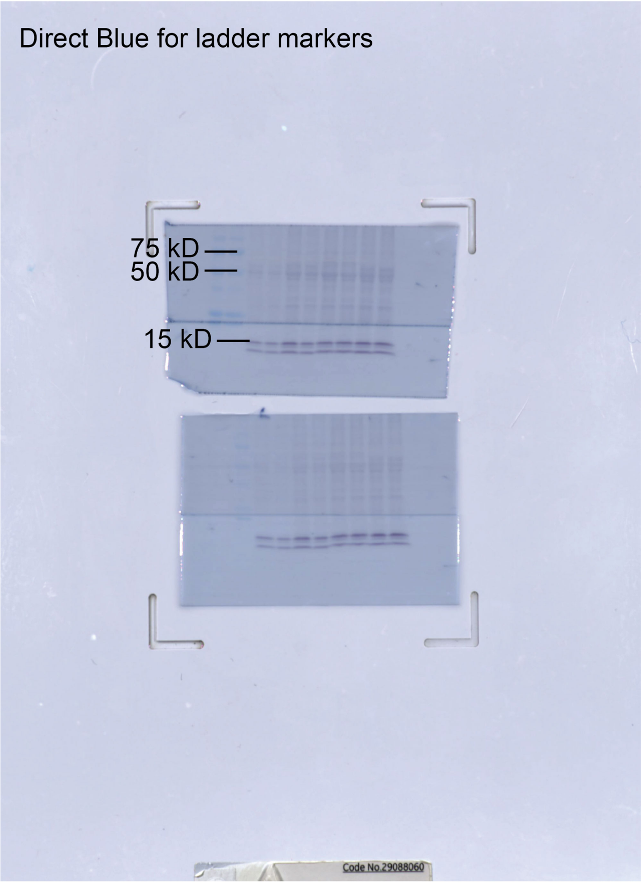


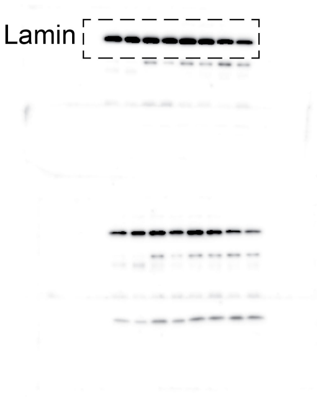


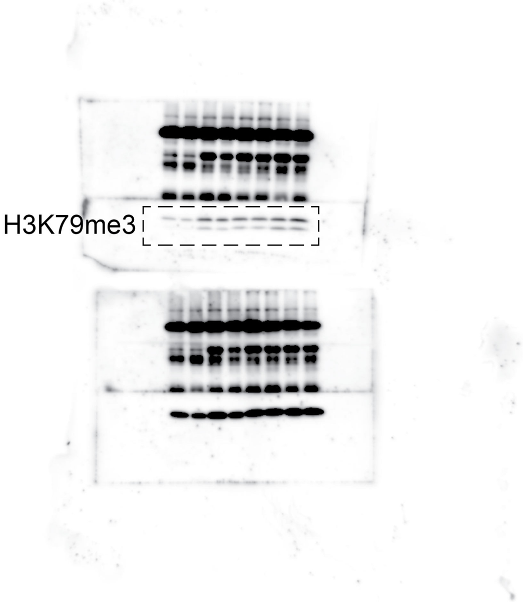


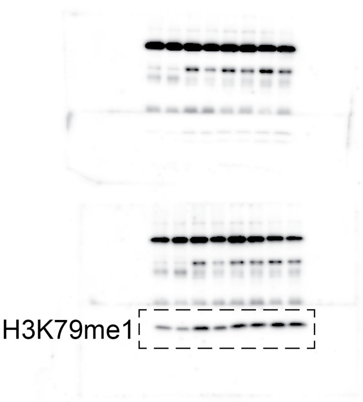

Figure S3B blot

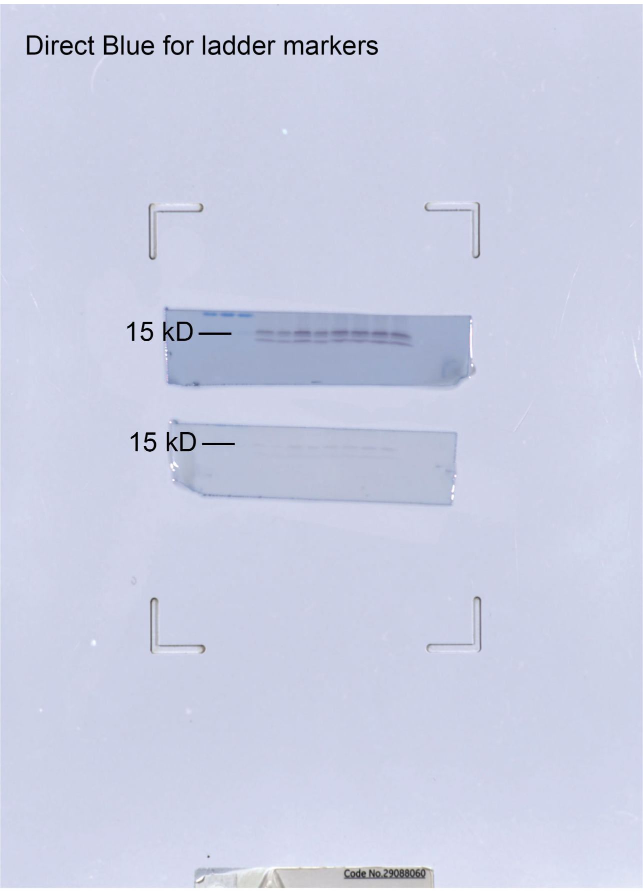


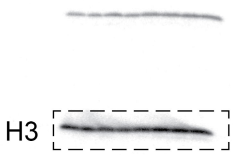


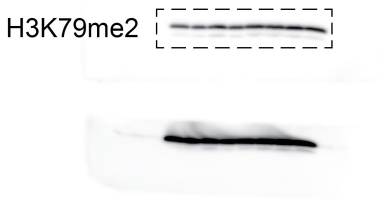

Figure S3D blot

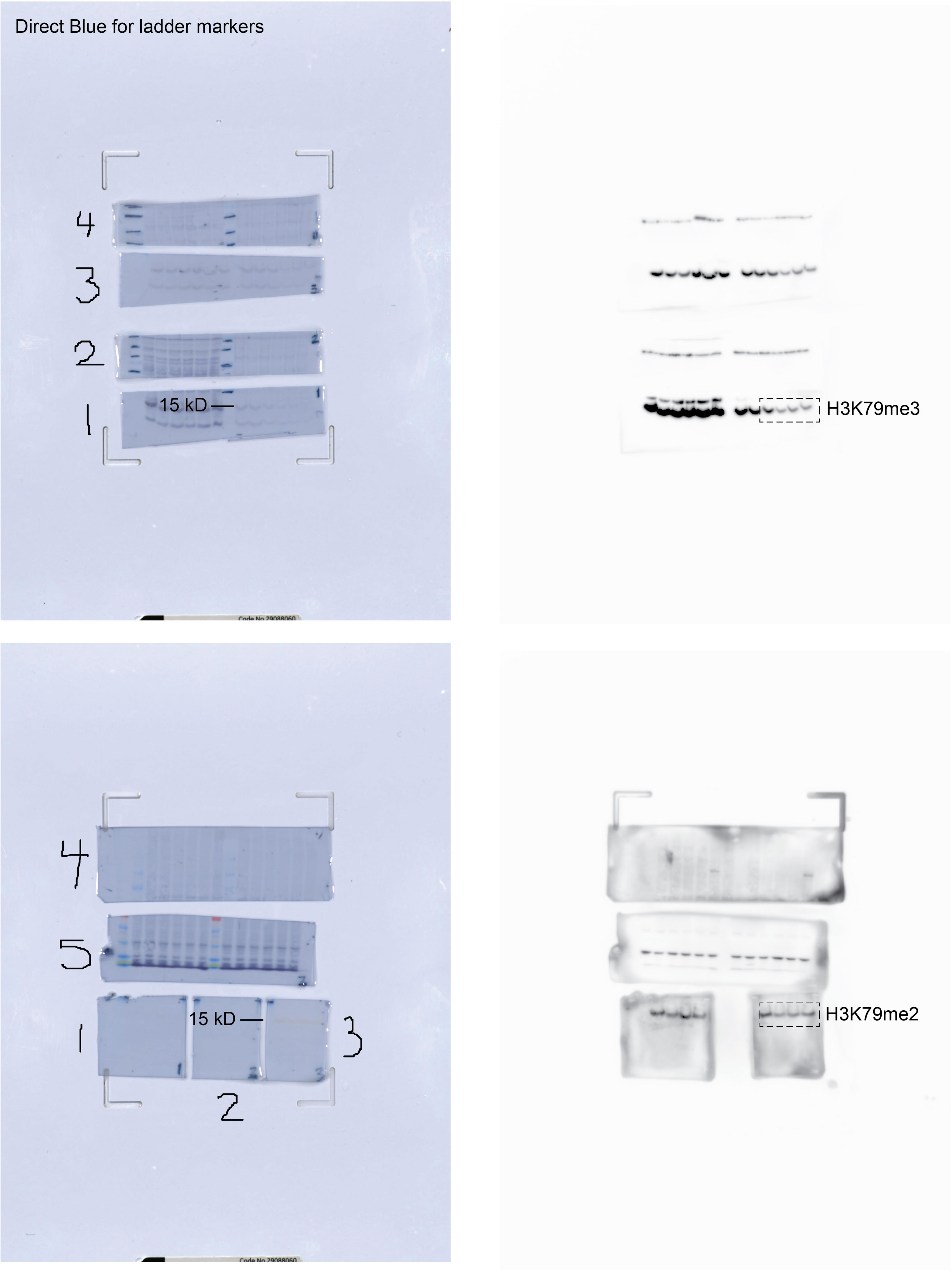

Figure S3D blot

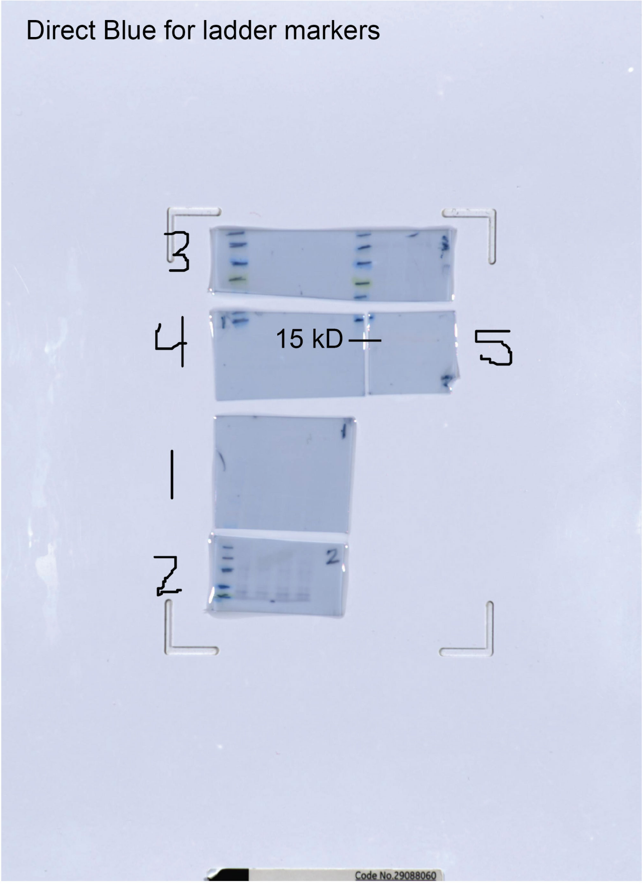


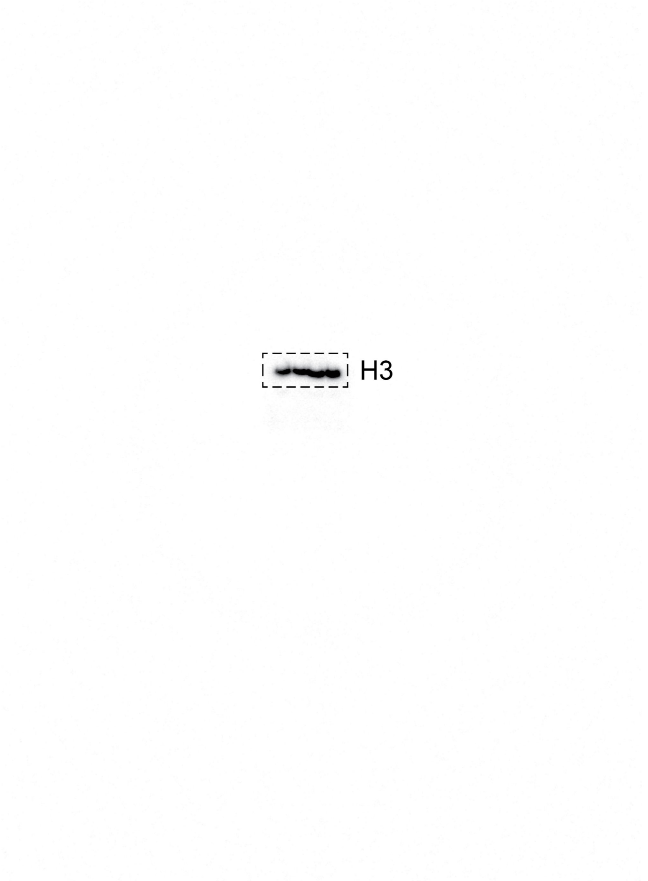

Figure S5B blot

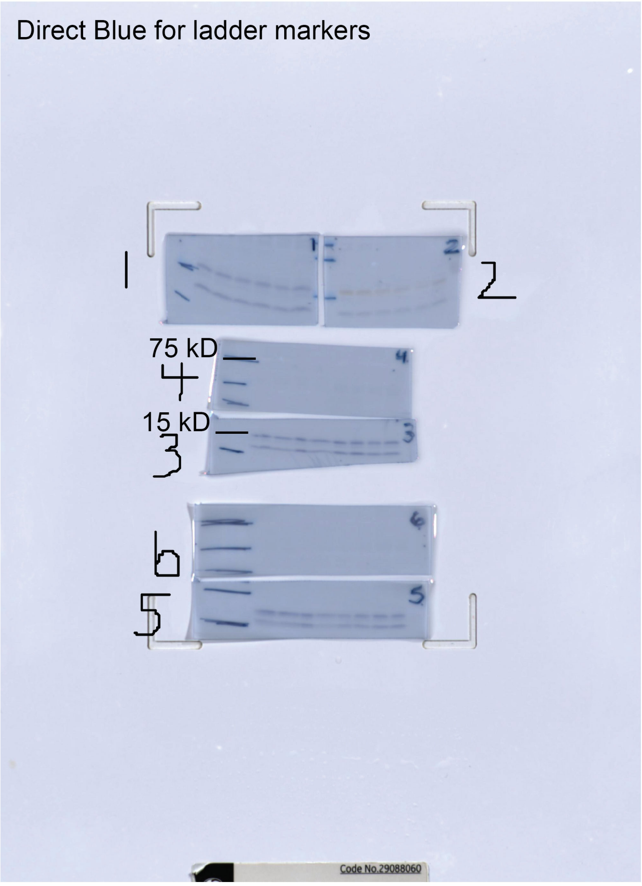


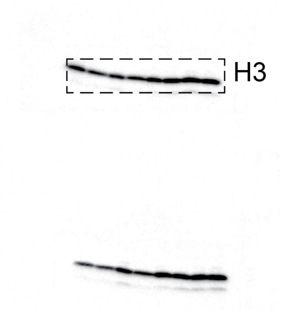


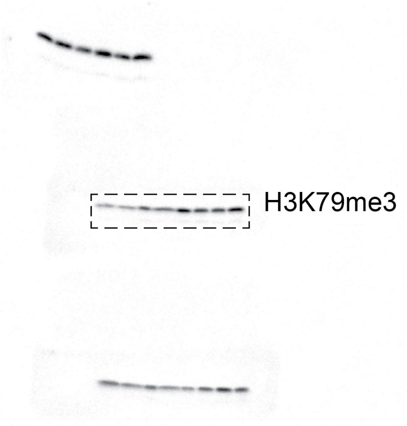


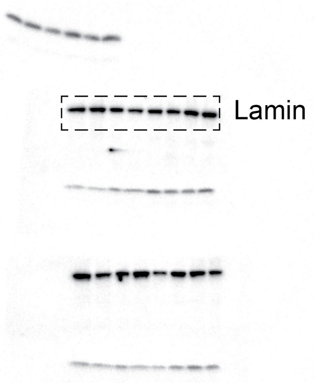

Figure S5D blot

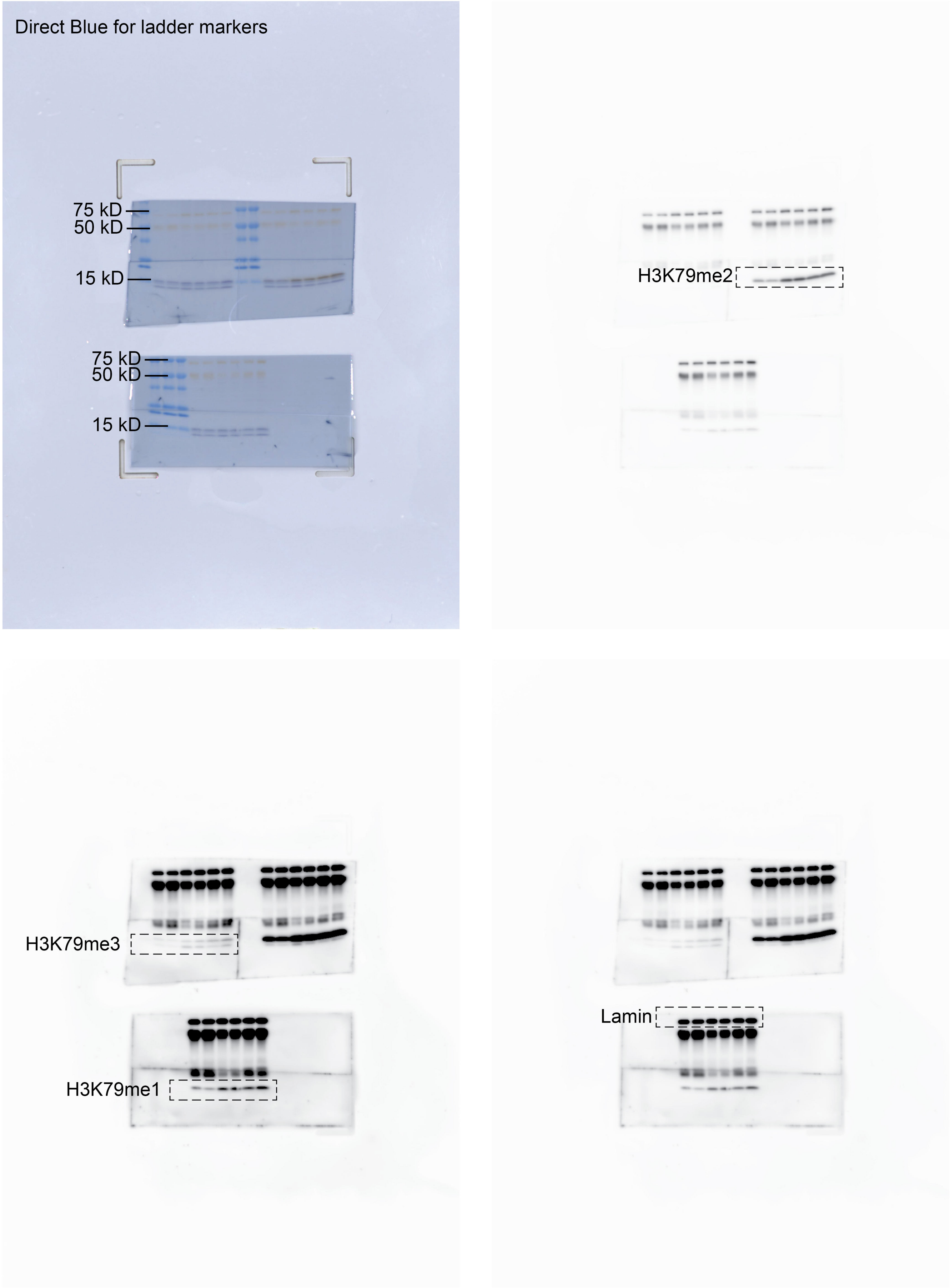

Figure S5D blot

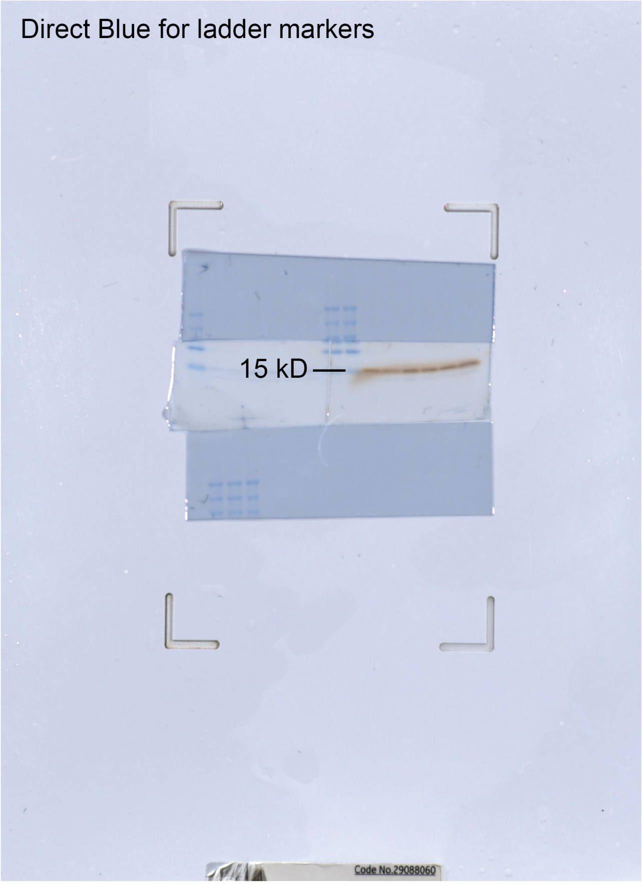


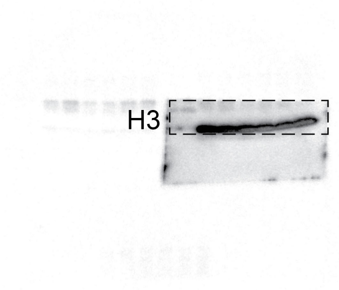

Figure S8B blot

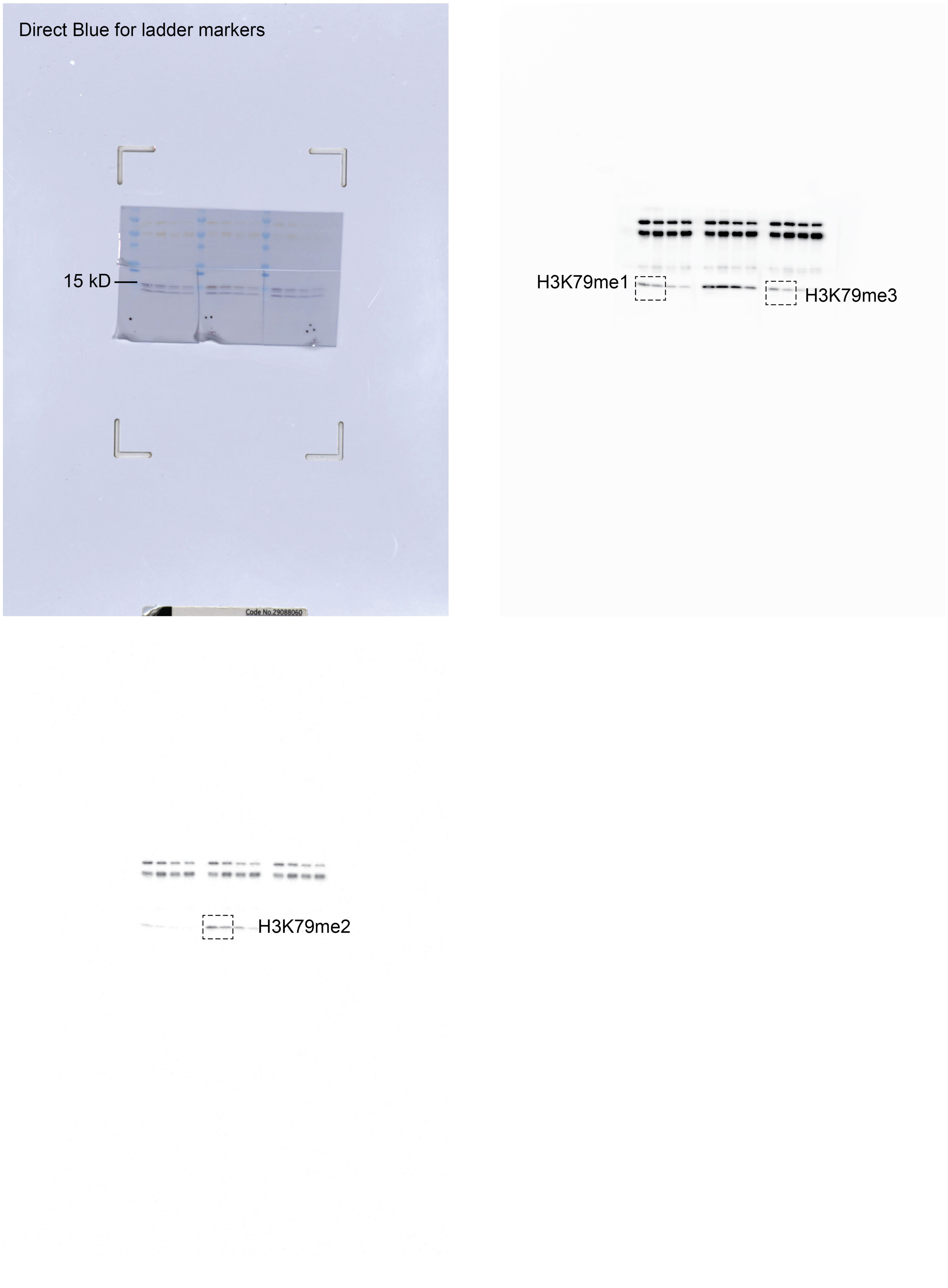

Figure S8B blot

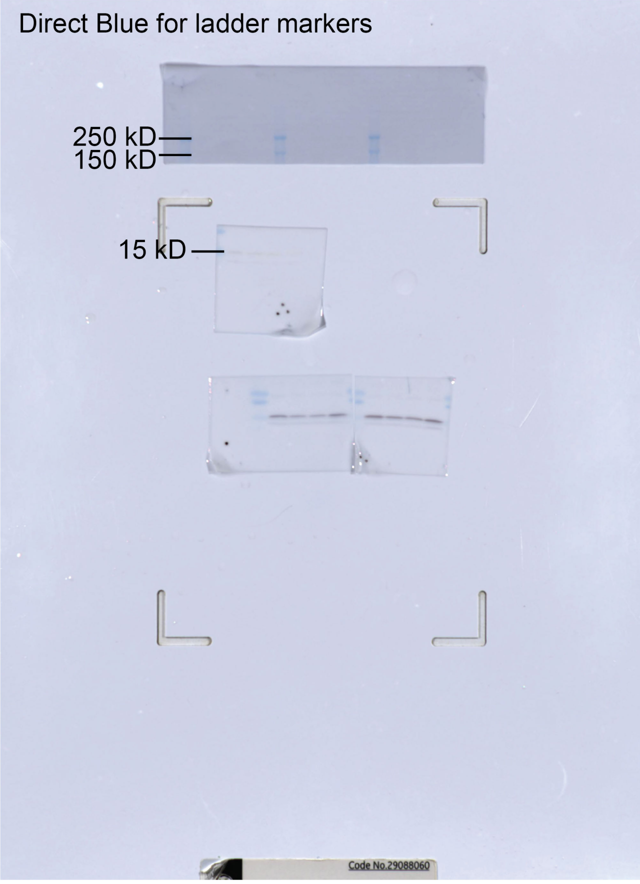


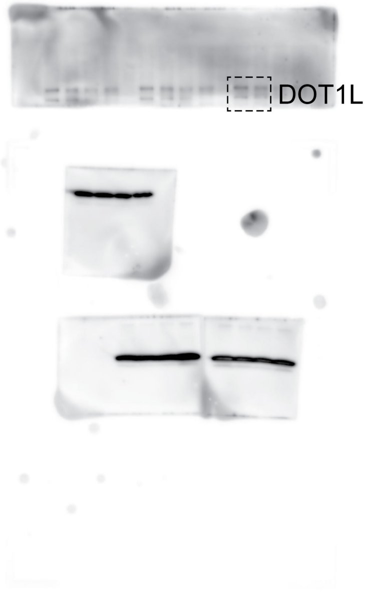


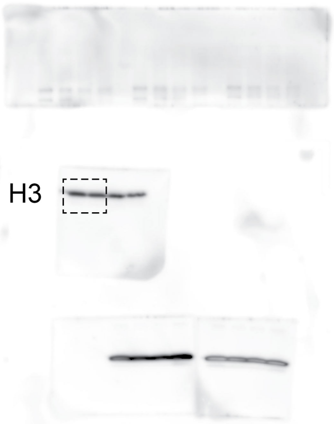
